# Predictive Modelling of Depression Treatment Response using Individual Symptoms and Latent Factors

**DOI:** 10.1101/2025.09.17.25335980

**Authors:** Sarah K Buehler, Jakob Heinzle, Mahmoud Eladawi, Chi Tak Lee, Anna K Hanlon, Veronica O’Keane, Siobhan Harty, Kevin Lynch, Klaas Enno Stephan, Claire M Gillan

## Abstract

Machine learning models have increasingly been used to identify predictors of treatment response in depression, and it is hoped that they may eventually help with clinical decision making. However, the performance of these models has generally been poor. One possible reason is that they are typically trained to predict aggregate scores of several depression symptoms; by contrast, individual symptoms may behave differently, be more predictable and/or more responsive to treatment. We tested this possibility by comparing the performance of machine learning models for predicting early response to psychotherapy based on 21 different outcome measures: (i) 16 individual depression symptoms, (ii) 4 latent symptom factors for sleep, appetite, motivation, and negative affect related symptoms, and (iii) total scores based on the widely used Quick Inventory of Depressive Symptomatology (QIDS). We used a large real-world dataset of 85 baseline features spanning sociodemographic, cognitive, clinical, lifestyle and physical health assessments in patients (N=776) initiating internet-delivered cognitive behavioural therapy (iCBT). For all 21 outcome measures, we developed elastic net models (N=543) and validated their performance in an unseen hold-out sample (N=233). In the hold-out dataset the model predicting total depression scores achieved an R^2^ of 40% variance explained, while there was substantial variability in model performance for individual symptoms (R^2^:2.1%-44%) and latent symptom factors (R^2^:26%-44%). Model comparisons revealed that most individual symptom and latent factor models with all 85 predictors were not superior to simpler benchmark models comprising only age, sex and baseline levels of the respective depression outcome measure. The benchmark was outperformed by models predicting total scores (*ΔR*^*2*^*=0*.*054, p=0*.*034)*, sad mood (*ΔR*^*2*^*=0*.*106, p=0*.*001)*, loss of interest (*ΔR*^*2*^*=0*.*079, p=0*.*021)* and a latent factor representing negative affect and thought (*ΔR*^*2*^*=0*.*054, p=0*.*038)*. Specifically, these models benefitted from additional predictors, such as treatment expectation, suicidal ideation, social support, or functional impairment. Our predictive modelling approach suggests new avenues towards a more patient-centred precision psychiatry, by providing clinicians with individual-level prognoses and predictors for interventions at the symptom level.

## Introduction

Major Depressive Disorder is amongst the most common and most burdensome diseases globally (Ferrari et al., 2024) and can cause severe life-long impairment if not treated effectively (Ferrari et al., 2013). Despite the widespread availability of psychotherapeutic and pharmacological treatments for depression, the most common of which are cognitive behavioural therapy (CBT) and antidepressant medications such as selective serotonin reuptake inhibitors (SSRIs), around one third to half of patients do not respond and even fewer experience remission (Cuijpers et al., 2014, 2021; Rush et al., 2006). It is possible the overall efficacy of these treatments could be improved through smart prescribing, i.e., matching patients to the treatment they are likely to respond to, based on their personal characteristics. Machine learning models have been developed for this purpose, using large and multimodal datasets to predict clinical outcomes (Chekroud et al., 2021; Rost et al., 2023). For instance, using demographic, clinical or genetic features, machine learning models have been shown to predict above chance-level the clinical response to internet-delivered CBT (iCBT) or antidepressant medication (Chekroud et al., 2016; Hornstein et al., 2021; Iniesta et al., 2018; Lee et al., 2025). However, the performance of these models is viewed by many as disappointing, explaining a relatively small amount of variance.

Typically, treatment response is determined using total scores from clinical questionnaires or scales, pooling information across items that indicate the severity or frequency of individual depression symptoms. However, it is possible is that individual symptoms might be more precisely predicted than these aggregates. Across individual patients, symptoms can vary so substantially that, depending on the diagnostic criteria, hundreds of different symptom profiles can qualify for the same depression diagnosis (Fried & Nesse, 2015a). For instance, an analysis of a large clinical trial for depression revealed 1,030 unique symptom profiles using the Quick Inventory of Depressive Symptomatology (QIDS), of which 48.6% were unique (Chekroud et al., 2017; Rost et al., 2023). Consequently, summary scores can lead to poor consistency across questionnaires that seek to diagnose the same psychiatric condition and obscure important symptom heterogeneity between individuals (Feczko et al., 2019; Newson et al., 2020). Extending this to treatment prediction, studies have found that some symptoms of depression are more responsive to treatments than others (Chekroud et al., 2017; Hieronymus et al., 2016; Madhoo & Levine, 2016; Uher et al., 2009). Specifically, antidepressants are particularly effective in reducing depressed mood, or feelings of guilt and suicidality, while other symptoms like sleep disturbances or fatigue may be less affected (Chekroud et al., 2017; Faries et al., 2000; Hieronymus et al., 2016; Madhoo & Levine, 2016). Other studies have directly compared treatments and in some cases found that specific symptoms are more responsive to medication vs CBT (Boschloo et al., 2019; Dunlop et al., 2018), while in other cases there are no differences in efficacy (Kappelmann et al., 2020). Previous studies have also examined the predictability of individual symptoms, but often at longer post-intervention timeframes, such as 3 months or more, with only very few predictors, often based on less than 5 different questionnaires, and most importantly without validation on unseen samples (Buckman et al., 2023; O’Driscoll et al., 2021). Consequently, there remains a gap in our understanding of how precision medicine tools like machine learning can be leveraged to use large multimodal feature sets to predict early response to treatment in unseen data.

To address this, the current project utilized multimodal real-world data from a longitudinal Precision in Psychiatry (PIP) study (Lee et al., 2023). This dataset allowed us to develop machine learning models (elastic net regression) to predict markers of early treatment response to psychotherapy based on the Quick Inventory of Depressive Symptomatology (QIDS) (Rush et al., 2003; Trivedi et al., 2004). Early clinical outcomes were assessed at 4-weeks, which is highly indicative of later treatment response at 8-14 weeks (Beard & Delgadillo, 2019; Nierenberg et al., 2000; Richards et al., 2020; Schibbye et al., 2014). In addition to comparing model performance for QIDS total scores versus symptom-level models, we also trained models to predict latent factors that might better balance precision and reliability while capturing meaningful symptom domains. We chose elastic net regression models due to the interpretability of their predictor importance rankings relative to other machine learning approaches. All models were trained using a large sample of patients who initiated psychotherapy (N=543), using a clinically effective and scalable internet-based form of CBT (Andersson et al., 2019; Karyotaki et al., 2021). This allowed us to determine which socio-demographic, psychosocial, physical health and lifestyle, clinical and cognitive measures meaningfully predict early changes in depression phenotypes in real-world treatment seeking patients. To validate our model and assess generalisability, we used an entirely separate hold-out dataset (N=233). Furthermore, to ascertain the treatment-specificity of our models we tested their generalisability in a smaller patient sample (N=107) initiating antidepressant medication in real-world service settings. The current study goes beyond previous approaches by comparing the suitability of individual symptoms with total scores as well as latent symptom-derived factors for predicting early treatment response and rigorously validating these models in two separate unseen hold-out samples to determine model generalisability and treatment-specificity. Alongside this, we provide novel systematic comparisons with quantitative significance testing to determine for which models a full feature set outperforms a more parsimonious benchmark model.

## Methods

### Study Design and Participants

The PIP study involves a longitudinal 4-week assessment of a range of multimodal measures, including clinical, demographic and behavioural data collected before and after initiating psychotherapy or pharmacotherapy. The detailed study design, procedure and sample description has already been published (Lee et al., 2023), as have the results of predictive modelling of total scores (Lee et al., 2025). The psychotherapy treatment group consisted of patients initiating a low-intensity evidence-based therapeutic intervention based on cognitive behavioural therapy (CBT) delivered online (Andersson et al., 2019; Karyotaki et al., 2021). A secondary treatment group consisted of patients initiating antidepressant medication. All patients were included within two days of starting treatment and a score of ≥10 on the Work and Social Adjustment Scale, a transdiagnostic measure of mental health related functional impairment. See supplementary material for more details. Following a previously established protocol for imputing missing scores in this study (Lee et al., 2025), the final retained sample size for all analyses was N=883 (psychotherapy group n=776, pharmacotherapy group n=107).

### Outcome Measures

#### Individual Depression Symptoms

Individual symptom scores were derived from the 16 items measured by the Quick Inventory of Depressive Symptomatology (QIDS), a highly reliable (Cronbach’s alpha =0.86) assessment tool that is widely used in clinical and research settings to determine the severity of major depressive disorder symptoms (Rush et al., 2003). The 16 individual item-level symptoms are based on the DSM-IV symptom criterion domains. These include 1) Insomnia (sleep onset), 2) Insomnia (mid-nocturnal), 3) Insomnia (early morning), 4) Hypersomnia, 5) Sad Mood, 6) Appetite decreased, 7) Appetite increased, 8) Weight decreased, 9) Weight increased, 10) Concentration or Decision-Making problems, 11) Self-critical thoughts, 12) Suicidal ideation, 13) Loss of interest, 14) Loss of energy, 15) Psychomotor slowing, 16) Psychomotor agitation. All symptom level scores are rated on a scale from 0-3 and were assessed before and after 4 weeks post-intervention to capture early response to treatment.

#### Latent Factors

We derived latent factors of depression symptomatology using an exploratory factor analysis (EFA) of the 16 symptom-level items measured by the QIDS at baseline, which allowed us to identify interpretable latent factors that account for the shared variance among symptoms (see Supplementary Material for details). Factor loadings were extracted to quantify the correlation between each symptom and the four latent factors (see figure 1). Based on these loadings, we assigned the following descriptive names to the four latent factors: appetite and weight, sleep disturbances, motivation and cognition, negative affect and thought. Each participant’s score on these latent factors was then computed via matrix multiplication of their symptom-level item scores with the factor weights (i.e., coefficients). These factor scores indicate the degree of difference from the sample mean and were then used as outcome measures for the predictive modelling.

**Figure 1.**
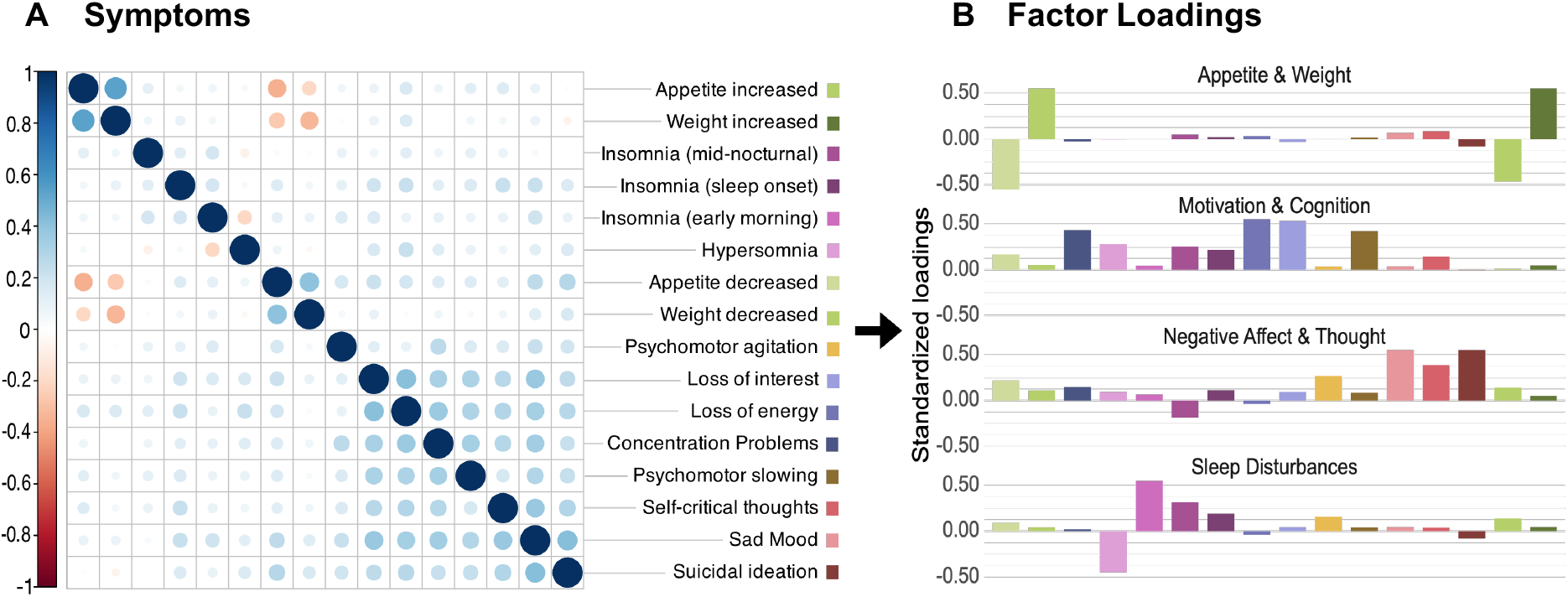
Exploratory Factor Analysis (EFA) pipeline to derive latent factors. First, a) the number of latent factors was determined by computing eigenvalues based on a spearman correlation matrix of all 16 QIDS symptoms at baseline. Then b) factor loadings were extracted to quantify the correlation between each symptom and the four latent factors, which we assigned the following descriptive names: appetite and weight, sleep disturbances, motivation and cognition, negative affect and thought. Finally, each participant’s score on these latent factors was computed via matrix multiplication of their symptom-level item scores with the baseline factor weights (i.e., coefficients).

#### QIDS Total Score

We compared the performance of models predicting individual QIDS symptoms and latent factors to QIDS total scores 4 weeks post-intervention. The QIDS total score ranges from 0 to 27 and is derived by summing the highest scores across nine symptom subdomains.

### Predictors of Interest

At baseline, a battery of self-report assessments and cognitive tasks was completed by participants. From this multimodal data we derived 85 predictors of interest across six categories: socio-demographics, psychosocial, physical health and lifestyle, clinical, treatment, and cognitive data. Total scores were computed for most clinical questionnaires, with the exception of the QIDS, for which we included the total scores as well as all 16 item-level scores as features (see supplementary materials for details). These predictors became the feature set for our full prediction model (hereafter referred to as full model). See Supplementary Material 1 for data processing and 2 for the full variable directory.

### Model Development

We used elastic net models, which combine lasso (L1) and ridge (L2) regularization to perform a linear regression with variable selection on the input feature set while maintaining interpretability of the coefficients. We carried out a 70:30 random split of the psychotherapy sample (N=776) into a training (N=543) and a hold-out test dataset for model validation (N=233), balancing baseline severity in total QIDS scores, gender, and sampling site. The pharmacotherapy sample (N=107) was reserved as a second hold-out dataset for testing model generalisability across treatment domains. The full modelling pipeline is illustrated in figure 2. We trained a model for each of the 16 QIDS symptoms, 4 latent factors and 1 total score on the pharmacotherapy training sample to predict post-intervention scores at the 4-week final assessment. To train the models and internally check their performance, we utilized 10-fold nested repeated cross-validation (nrCV). To evaluate model performance we used R squared (R^2^), which captures the proportion of variance in the outcome measure explained by the model. See supplementary table 2 for a complete set of performance metrics including root mean squared error (RMSE), mean absolute error (MAE) and correlation of actual to predicted scores.

**Figure 2.**
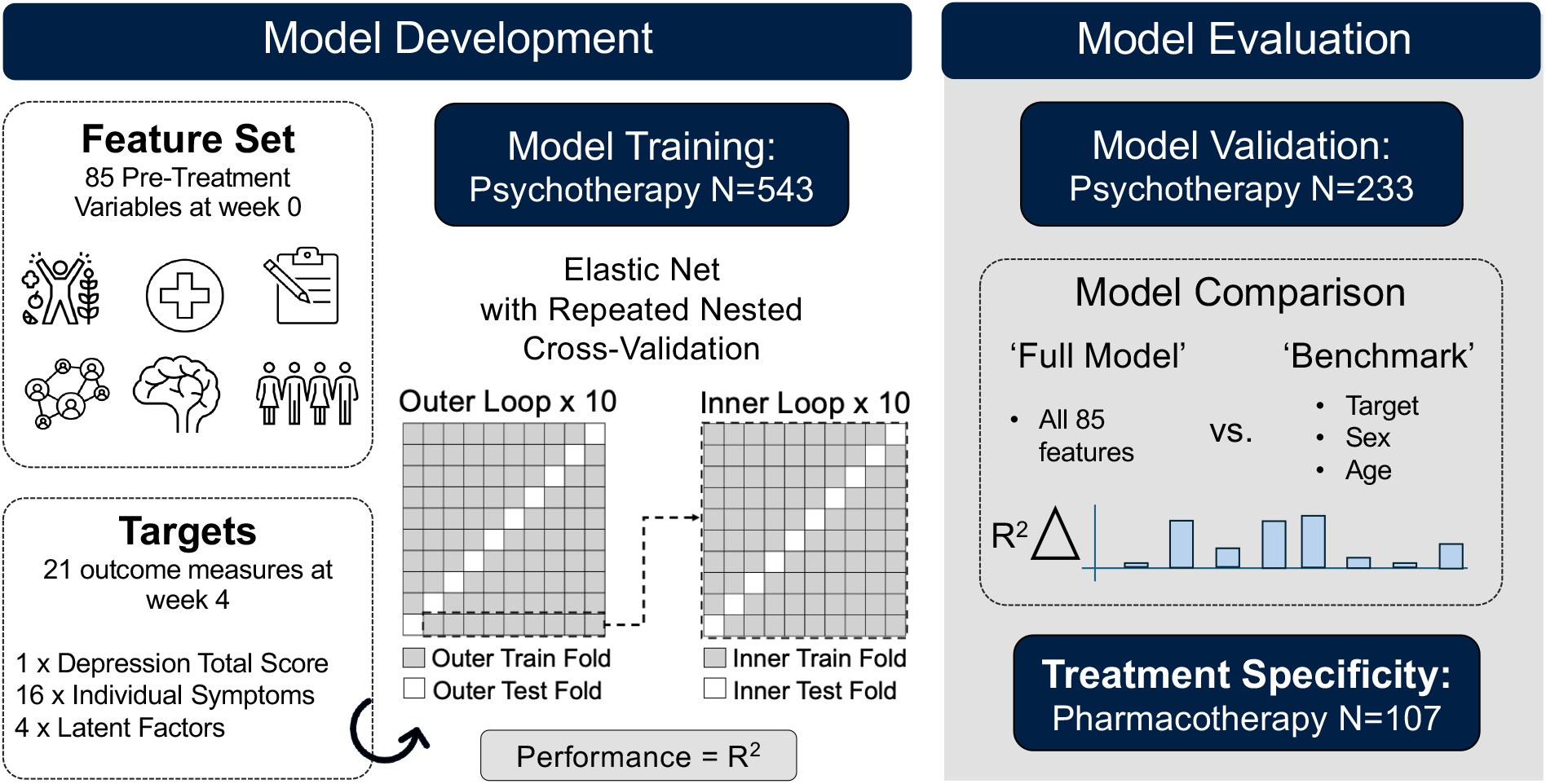
Predictive Modelling Pipeline. During model training, we used 70% of the psychotherapy sample (N=543) to train an elastic net regression model using nested repeat cross validation (nrCV) with 10 inner loops (with 10 iterations each) for hyperparameter optimisation and feature selection and 10 outer loops for internal model evaluation. A ‘full model’ was trained based on 85 predictors of interest from socio-demographics, psychosocial, physical health and lifestyle, clinical, treatment, and cognitive data to predict post-intervention scores at the 4-week final assessment for each of the 16 symptoms, 4 latent factors and 1 total score for depression. Model testing was performed on a hold-out sample, from the remaining 30% of the psychotherapy sample (N=233) and model specificity was determined using an unseen hold-out pharmacotherapy sample (N=107). Then a model comparison was conducted to determine whether the ‘full model’, using all 85 predictors of interest, offered better predictive performance - in terms of variance explained (R^2^) - than a benchmark model, using only the pre-intervention symptom scores of the outcome measure, age and sex.

### Model Evaluation

To evaluate the models, we first validated their performance by determining how well they generalize to the unseen hold-out psychotherapy sample (N=233). Then we conducted comprehensive model comparisons to determine if the predictive power of each model was driven primarily by a benchmark model containing as predictors only the pre-intervention symptom scores of the outcome measure (plus age and sex). To evaluate whether the predictive performance of the full model containing all 85 predictors significantly outperformed these benchmark models, we implemented a permutation-based significance test based on R^2^ differences (see supplementary material). Finally, for the models that survived this comparison, we further assessed their treatment-specificity using a held-out pharmacotherapy sample (N=107).

## Results

### Model Development and Validation

During model development the cross-validated performance (R^2^) for QIDS total scores in the psychotherapy training sample (n=543) was 39.3% (Figure 3). This generalised well to the psychotherapy hold-out sample (n=233) during model validation (R^2^=39.9%). For individual symptoms, model performance was highly variable. While some symptoms enabled higher predictive accuracy during cross-validation in the training dataset, such as insomnia (sleep onset) with 48.8% or suicidal ideation with 47.8%, their performance in the unseen psychotherapy validation dataset was substantially reduced to 33.2% and 35.9% respectively. Other symptom models explained less variance but generalised better to the hold-out validation sample, including hypersomnia (R^2^ = 43.8%), sad mood (R^2^ = 32.5%), and appetite increases (R^2^ = 34.2%), self-critical thoughts (R^2^ = 31.9%). The models developed to predict latent factors generally exhibited more stable generalisation, with the most variance in the psychotherapy validation sample explained by the negative affect and thought latent factor (R^2^ = 43.5%).

**Figure 3.**
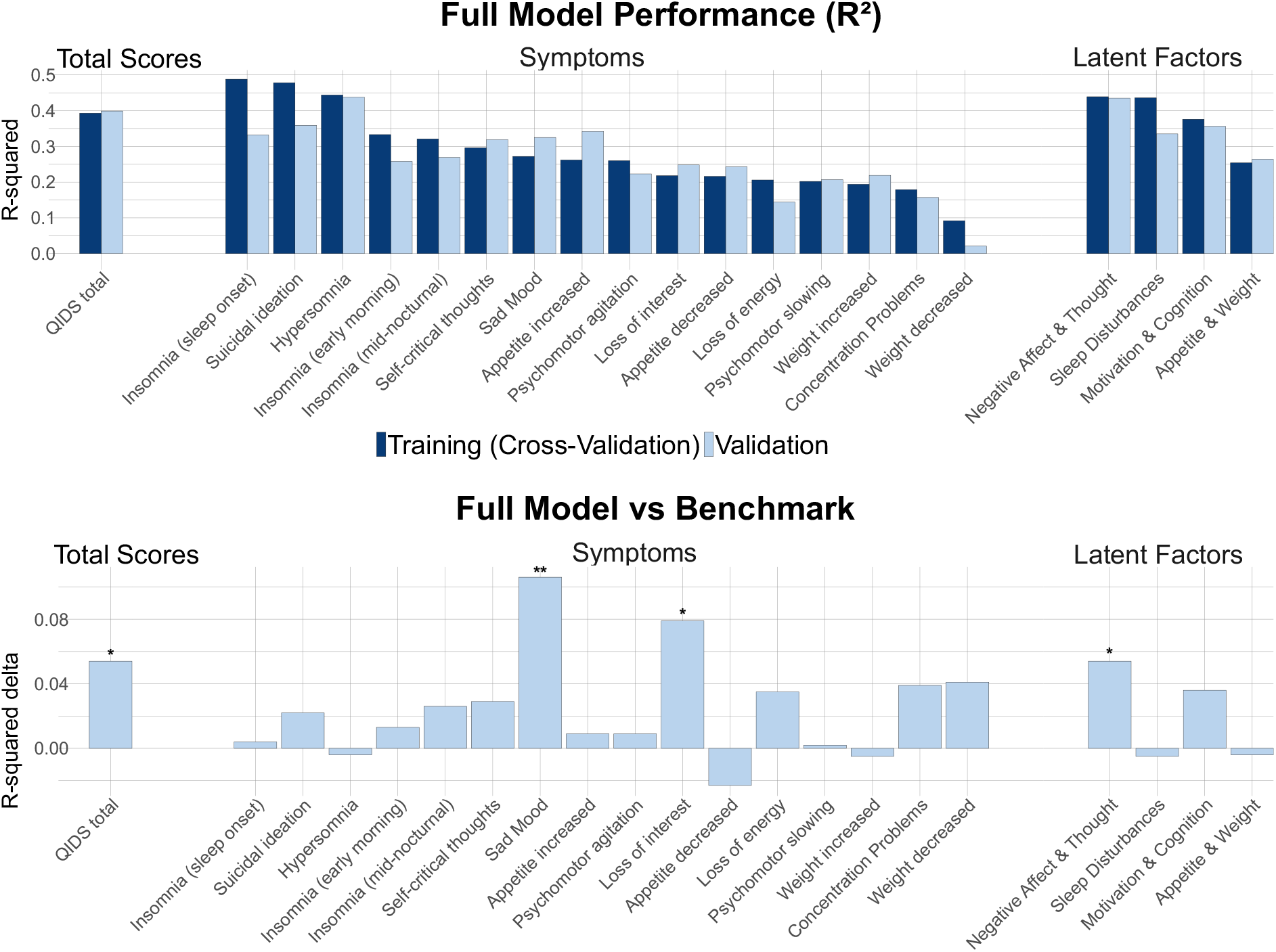
Model Performance (R-squared) for total scores, symptoms and latent factors. In the upper panel the full model performance (R-squared) is shown for the cross-validated training sample (dark blue) and the hold-out psychotherapy validation sample (light blue). The lower panel shows the difference in model performance (R-squared delta) between the full predictor models and benchmark models, which contain only the baseline outcome measure, age, and sex. The significance symbols indicate where the full model significantly outperformed the benchmark.

### Model Comparison, Treatment Specificity and Predictor Importance

All models in the psychotherapy and pharmacotherapy hold-out samples performed significantly better than chance level (see supplementary material 1). However, it is possible for the prediction of future outcomes to be driven by their baseline values and basic demographics. To test if this was the case for some of our models, we performed model comparison of the full model versus one trained on a small benchmark feature set comprising only baseline levels of the target (outcome) variable, age and sex. The model comparison in the psychotherapy validation sample was statistically significant for total scores (ΔR^2^=0.054, p=0.034), symptoms of sad mood (ΔR^2^=0.106, p=0.001) and loss of interest (ΔR^2^=0.079, p=0.021), as well as for the latent factor for negative affect and thought (ΔR^2^=0.054, p=0.038).

To further understand how properties of the target variable related to model performance, we tested if those variables with greater variability in patient responses (i.e., higher entropy in score distributions) were more accurately predicted and found this was the case (Spearman’s p=0.57, p-value=0.024, see supplementary material 1 for details).

Next, we aimed to test the treatment specificity for the four target variables that had superior performance to the benchmark model. There was evidence that the models were specific to iCBT, as they performed markedly worse on the antidepressant sample and were not superior to the benchmark (total scores; R^2^=0.221, ΔR^2^=0.04, p=0.273, sad mood; R^2^=0.09, ΔR^2^=0.004, p=0.921, loss of interest; R^2^=0.072, ΔR^2^=0.004, p=0.912, latent negative affect and thought factor; R^2^=0.278, ΔR^2^=0.066, p=0.055). Finally, we inspected the coefficients of all significant predictors for these models (Figure 4).

**Figure 4.**
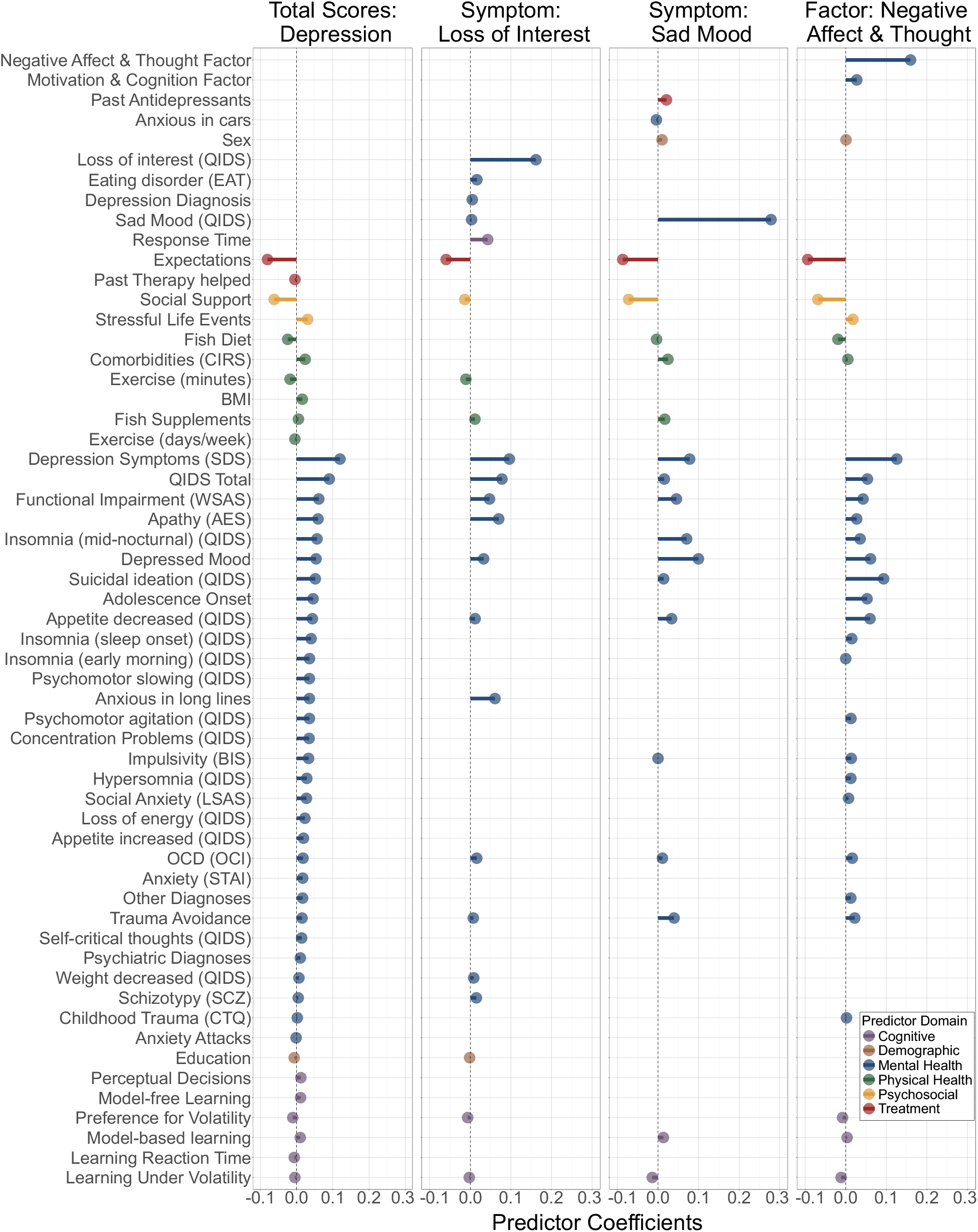
Contributions of significant predictors from elastic net models for interpretability. Predictor coefficients are shown for full models that significantly outperformed the benchmark in the psychotherapy validation sample, which includes the total score, sad mood symptom, loss of interest symptom, and negative affect & thought latent factor. This shows the contribution of predictors across the cognitive (purple), demographic (brown), mental health (blue), physical health (green), psychosocial (yellow) and treatment (red) domain.

## Discussion

We developed machine learning models for 21 different outcome measures to predict treatment response to iCBT in depression, comparing how well we could predict total scores relative to latent factors and individual symptoms. There was considerable variability in the performance of these models, but total scores were among the highest performing, only outperformed in the iCBT hold-out set by hypersomnia and a ‘negative affect and thought’ factor. However, several models performed well simply because their baseline level of the target was highly predictive on their follow-up scores (e.g. hypersomnia). We tested whether additional demographic, cognitive, psychosocial, treatment, physical and mental health measures improved prediction by comparing the performance of models trained on all available features to models trained on just baseline scores for the target variable, age and sex (i.e., the ‘benchmark’ model). Only four models predicting total scores, sad mood, loss of interest and the negative affect and thought factor significantly outperformed the benchmark models.

The winning prediction model overall was the negative affect and thought latent factor model, which had the best model validation performance out of those that demonstrated superiority to the benchmark. This latent factor captured a range of symptoms such as suicidal ideation, sad mood and self-critical thoughts, but performed substantially better than any of these in isolation. Therefore, this latent factor might capture a depression phenotype that is particularly well suited to predictive modelling in CBT. In the clinical domain, this was most strongly predicted by depression symptoms (SDS), closely followed by suicidal ideation, at baseline. This suggests that targeted monitoring of these specific symptoms may be beneficial for treatment outcomes. From an analytical perspective, latent factors such as negative affect and thought may offer a suitable target for predictive modelling by achieving better reliability than can be achieved with single symptom models, while still focusing on a more specific element of depression than is captured in the total score used in most trials. However, regardless of predictive performance, modelling approaches that take into account individual symptoms may be preferred by patients, because certain symptoms are more distressing or impairing than others (Fried & Nesse, 2014). Individual symptom models therefore allow more personalisation of treatments not just to individual patients, but the symptoms that those patients most want to tackle.

When tested on an unseen antidepressant medication sample, the full versus benchmark model comparison of the four validated models did not generalise, suggesting treatment-specificity. This is important as it confirms that model performance is not driven purely by a general tendency for symptoms to improve or worsen over time and is instead related to the type of treatment received. That said, to confirm this is the case, precision treatment allocation studies require randomised designs and counterfactual tests of model performance.

In terms of model explainability, the total depression score model retained the most predictors, while the symptom-level and factor models had more parsimonious feature sets, revealing interesting patterns of overlap and distinction in predictor importance. Across all four of these models, most predictor coefficients consistently emerged from the mental health domain. Higher baseline scores of the respective outcome measure and Zung self-rating depression scale (SDS) were most predictive of worse post-intervention outcomes. Conversely, better baseline treatment expectations and social support were strong predictors of reduced post-intervention severity, with striking consistency across all models. Treatment expectations, in particular, are increasingly recognised as a crucial determinant of patient outcomes (Constantino et al., 2018; Richler, 2022). Importantly, these can be shaped by therapists through goal setting and expectation management even before patients begin treatment. Likewise, clinicians can offer social support resources and programs alongside therapy to further improve outcomes. For the total scores and loss of interest symptom, post-intervention scores were positively predicted by apathy, which can suggest impairments in the motivation and ability to enjoy otherwise pleasurable things, and inversely by exercise, which may be a suitable lifestyle intervention to improve this outcome (JIa et al., 2025). Meanwhile, cognitive measures performed relatively poorly across all models. The exception to this was slower response time, uniquely associated with loss of interest, which may reflect a behavioural manifestation of this symptom. We were also able to demonstrate that for the remaining symptom-level models, which were non-significant in our model comparison, basic benchmark measures such as baseline scores, age and sex are sufficient to predict outcomes, with varying performance levels. These insights can be useful for future predictive modelling research and substantially reduce the burden of data collection for patients and clinicians.

In supplementary analyses, we further assessed how the distributional properties of symptom scores related to model performance. As a sanity check, we were able to show that models trained on symptoms with more variability (i.e., higher entropy) performed better. This confirms that more variance to explain in the outcome measure can enhance model discrimination, while imbalanced or rare response items may be less discriminable. It is also consistent with the idea that imbalanced response patterns can reduce the variance available to be explained by predictive models, making it more difficult to detect meaningful patterns. Linear models, such as elastic net regressions, are known to perform optimally with more normally distributed outcomes. However, the distributions of symptom-level scores in our samples suffered from sparsity and zero-inflation, with many patients reporting scores of zero. This can lead to reduced predictive power and biased parameter estimates (Rožanec et al., 2025). Interestingly, aggregating symptoms to latent factors is analytically advantageous for predictive modelling by providing more normally distributed outcome measures with a greater continuous spread in variance. This suggests that factor analysis may be a promising approach to generate insights based on symptoms while dealing with the limitation of individual symptom-level target variables having limited variance and not being normally distributed.

The present study has several limitations. We prioritized consistency and comparability across outcome measures by evaluating the same type of machine learning model, but future work may wish to conduct a more comprehensive comparison of different linear and non-linear as well as hybrid model types for each outcome measure. While we initially considered ordinal and multinomial elastic net regression models for our item-level prediction, these approaches had several conceptual limitations that made them unsuitable for our purposes. For instance, they require further assumptions about whether to model the cumulative probabilities of being at or below, at or above, or directly adjacent to a certain score. This would limit the interpretability and consistency of model predictions across individual symptoms, which have entirely different and often skewed distributions with no meaningful score thresholds. Importantly, such an approach would not allow us to compare these models with the continuous total scores used in clinical settings. While we provide evidence for treatment specificity by showing that our final models did not outperform the benchmark in our pharmacotherapy validation sample, this should not be mistaken for a significant model by treatment modality interaction, which would be more conclusive but require a larger and more balanced combined model. Finally, our data was based on a real-world observational study without placebo-controlled randomised interventions and causal effects can therefore not be inferred

## Conclusions and Clinical Impact

In this study, we developed treatment-specific machine learning models that can generate patient-level outcome predictions for total depression scores, individual symptoms and latent factors, using multimodal real-world data from treatment-seeking patients. This modelling approach is highly flexible, and can also be extended to feature sets enriched with neural data, such as functional neuroimaging (Dunlop et al., 2017) or electroencephalographic measures (Rajpurkar et al., 2020), to bridge the current gap between biological mechanisms and psychopathology (Tiego et al., 2023). This work may inspire future computational and predictive modelling approaches to provide clinicians with patient-level treatment prognosis tools based on symptom profiles. In particular, the graded approach presented in this paper may be useful for identifying specific pre-intervention predictors that may represent targets for intervention at different levels of granularity, from individual symptoms to symptom factors to diagnostic categories. We hope that this work will help opening new avenues towards a more patient-centred precision psychiatry, with the potential to improve clinical care and patient outcomes.

## Data Availability

The data used in this study can be made available upon reasonable request, at the discretion of the corresponding authors.

## Notes

### Competing Interest Statement

DR and SH are current employees of and hold shares in SilverCloud Health, Amwell. CTL became an employee of SilverCloud Health, Amwellpost-completion of this researchas part of her PhD studentship, funded by SilverCloud Health, Amwell and the Irish Research Council via an industry-academia partnership; she holds no shares in the company.CMG reports no financial relationships with commercial interests but was the primary supervisor of CTL. KES acknowledges support by the Rene and Susanne Braginsky Foundation and the ETH Foundation.

### Funding Statement

This work was funded by a fellowship awarded to CMG from MQ: transforming mental health (MQ16IP13). CMG holds additional funding from Science Foundation Irelands Frontiers for the Future Scheme (19/FFP/6418), and a European Research Council (ERC) Starting Grant (ERC-H2020-HABIT). SKB was supported by a Biotechnology and Biological Sciences Research Council [grant number: BB/T008709/1] Ph.D. Studentship.

### Author Declarations

The study obtained ethical approval from the Research Ethics Committee of School of Psychology, Trinity College Dublin and the Northwest-Greater Manchester West Research Ethics Committee of the National Health Service, Health Research Authority and Health and Care Research Wales. All methods were performed in accordance with the relevant guidelines and regulations. All participants provided informed consent to participate in the study online before they proceeded to the screening stage of the study.

### Summary of Updates

some typos in the abstract

