## Supplementary Material 1 for "Predictive Modelling of Depression Treatment Response using Individual Symptoms and Latent Factors"

**STUDY DESIGN AND PARTICIPANTS** ... 1
**DATA PROCESSING** ... 1
**SCORE SUMMARY FOR QIDS TOTAL, ITEMS AND LATENT FACTORS** ERROR! BOOKMARK NOT DEFINED.
**SUMMARY OF PREDICTORS OF INTEREST** ... 2
**EXPLORATORY FACTOR ANALYSIS** ... 4
**DISTRIBUTIONS OF QIDS TOTAL, SYMPTOM-LEVEL AND FACTOR SCORES PRE- AND POST-INTERVENTION** ... 5
**ELASTIC NET REGRESSION MODEL PERFORMANCE** ... 7
TRAINING SAMPLE (PSYCHOTHERAPY) ... 7
HOLD-OUT VALIDATION SAMPLE (PSYCHOTHERAPY) ... 12
HOLD-OUT VALIDATION SAMPLE (PHARMACOTHERAPY) ... 18
**PROPERTIES OF QIDS SYMPTOM-LEVEL DISTRIBUTIONS AND MODEL PERFORMANCE** ... 23
**MODEL COMPARISON OF FULL TO BENCHMARK MODEL** ... 25
TOTAL SCORE ... 25
SYMPTOMS ... 26
FACTORS ... 37

#### Study Design and Participants

Participants in the psychotherapy treatment group completed clinician-guided modules of cognitive behavioural therapy (CBT) on the SilverCloud platform. They were recruited from a mental health charity in Dublin, Ireland and from a National Health Service (NHS) Talking Therapies clinic in Berkshire, UK. The pharmacotherapy treatment group initiating antidepressant medication was recruited via online (Google Ads, social media platforms, mental health charities) and in-print advertisements (pharmacies, general practitioners, counselling clinics, newsletters).

Participants were not required to discontinue other treatments to be eligible for this study and could be included in the pharmacotherapy group if they were initiating or switching to a new antidepressant medication. As a result, 24% of the psychotherapy sample reported concurrent antidepressant medication use and 34% of the pharmacotherapy sample were concurrently receiving psychotherapy. This allowed us to assess treatment specific effects by collecting data pre- and post-intervention initiation with either CBT or antidepressants, while ensuring our samples reflect the clinical reality of symptom heterogeneity and concurrent treatment use (see Lee et al., 2023 for full sample characteristics).

#### Data processing

All data processing and analysis was carried out in R (2024.09.1). For consistency we used a previously established protocol to preprocess the data (Lee et al., 2025). To deal with missing predictors while avoiding target data leakage, we used the median for continuous as well as ordinal predictors and mode for categorical predictors from the training dataset, after our train-test sample split, to impute missing data in the model training and hold-out testing datasets. Nominal variables were binarized, with the exception of 'age of onset' which was dummy transformed to childhood, adolescence or adulthood. Continuous variables in the training and testing datasets were standardised by subtracting the mean and dividing by the standard deviation both computed exclusively on the training sample to avoid data leakage. Variables with rare endorsement, defined as having <10% in any response option for binary items and ≥90% of any response option for interval/continuous variables, were eliminated. Variables with high collinearity (r≥0.8) as well as variables with near zero variance were also removed. To impute missing outcome measures at follow-up, we used a linear regression model for each symptom measure to predict scores at week four based on scores at the previous week three timepoint based on the training sample data. We then applied the intercept and beta coefficient estimates from this formula to impute missing scores at follow-up based on the non-missing scores from week three in both the training and testing datasets separately, in order to avoid target data leakage.

#### Summary of Predictors of Interest

| Predictor | Coding |
|---|---|
| **Demographic** | |
| Age | Ratio |
| Sex | Binary (male, female) |
| Marital Status | Binary (single, not single) |
| Education Level | Ordinal (< third level, completed third level, > third level) |
| Employment Status | Binary (employed, unemployed) |
| Subjective Social Status | Ordinal (0-10) |
| **Physical Health** | |
| Exercise days/week | Ratio (0-7) |
| Exercise minutes/day | Ordinal (0, 10, 20, 30, 40, 50, 60+) |
| Diet Quality | Ordinal (Very poor, Poor, Fair, Good, Very good, Excellent) |
| Diet Fish Consumption | Ordinal (rarely/never, once a month, twice a month, once a week, twice a week, every second day, once a day, more than once a day) |
| Diet Fish Supplements | Ordinal (rarely/never, once a month, twice a month, once a week, twice a week, every second day, once a day, more than once a day) |
| Drug use at present | Binary (yes, no) |
| Drug use at past | Binary (yes, no) |
| Cumulative Illness Rating Scale (CIRS) total | Interval |
| Patient Health Questionnaire (PHQ-15) 5 pain items total | Interval |

| | |
|---|---|
| Smoking use at present | Binary (yes, no) |
| Smoking use at past | Binary (yes, no) |
| BMI based on height & weight | Ratio |
| **Psychosocial** | |
| Stressful Life Events (SRRS) total | Interval |
| Childhood Trauma (CTQ) total | Interval |
| Scale of Perceived Social Support (MSPSS) total | Interval |
| Perceived Stress Scale (PSS) total | Interval |
| **Mental Health** | |
| Lifetime mental health episodes | Ordinal (<2, 2-5, 5+) |
| First episode onset: Adolescence | Binary (Yes, No) |
| First episode onset: Adulthood | Binary (Yes, No) |
| Current mental health episode number of days | Ratio |
| Psychiatric Diagnoses total | Ratio (0-13) |
| Psychiatric Diagnosis Depression | Binary (Yes, No) |
| Psychiatric Diagnosis Generalised Anxiety Disorder (GAD) | Binary (Yes, No) |
| Psychiatric Diagnoses Other (excluding Depression and GAD) | Ratio (0-11) |
| Psychiatric Diagnoses (Family) | Ordinal (None, 1-2, 3+) |
| 8 Psychiatric Symptoms based on predictors in (Chekroud et al., 2016) | Binary (Yes, No) |
| Apathy Scale (AES) total | Interval |
| Alcohol Use Disorder (AUDIT) total | Interval |
| Impulsivity Scale (BIS) total | Interval |
| Eating Disorder Sale (EAT-26) total | Interval |
| Schizotypy (SSMS-R) total | Interval |
| Social Anxiety Scale (LSAS) total | Interval |
| State Anxiety Scale (STAI-T) total | Interval |
| Obsessive Compulsive Disorder Scale (OCI-R) total | Interval |
| Self-Rating Depression Scale (SDS) total | Interval |
| Quick Inventory of Depressive Symptomatology – Self-Report (QIDS-SR) total score | Interval |
| All 16 QIDS-SR item scores | Ordinal (0, 1, 2, 3) |
| Work and Social Adjustment Scale (WSAS) total | Interval |
| **Treatment** | |
| Past Psychological Therapy | Binary (Yes, No) |
| Past Antidepressant Treatment | Binary (Yes, No) |
| Past Psychological Therapy significantly improved symptoms | Ordinal (none, 1-4 times, 5+ times) |

| | |
|---|---|
| Past Antidepressant Treatment significantly improved symptoms | Ordinal (none, 1-4 times, 5+ times) |
| Current Treatment Expectations | Interval (0-9; no better to completely better) |
| **Cognitive** | |
| Confidence (Dot Discrimination Task) | Ratio |
| Metacognitive Efficiency (Dot Discrimination Task) | Ratio |
| Model-Free planning index (Two-Step Reinforcement-Learning Task) | Ratio |
| Model-based planning index (Two-Step Reinforcement-Learning Task) | Ratio |
| Choice stickiness (Two-Step Reinforcement-Learning Task) | Ratio |
| Mean Reaction Time (Two-Step Reinforcement-Learning Task) | Ratio |
| Reaction Time after transition (Two-Step Reinforcement-Learning Task) | Ratio |
| Learning rate in stable environment (Aversive learning task) | Ratio |
| Learning rate in volatile environment (Aversive learning task) | Ratio |
| Risk preference in stable environment (Aversive learning task) | Ratio |
| Risk preference in volatile environment (Aversive learning task) | Ratio |
| Choice stochasticity in stable environment (Aversive learning task) | Ratio |
| Choice stochasticity in volatile environment (Aversive learning task) | Ratio |

***Supplementary Table 1**. Predictors of Interest. A list of all the predictors included in the full model across the demographic, physical health, psychosocial, mental health, treatment and cognitive domain and their coding on a ratio, binary, ordinal or interval scale.*

#### Exploratory Factor Analysis

To identify latent factors underlying depressive symptoms measured by the 16-item Quick Inventory of Depressive Symptomatology (QIDS), we conducted an Exploratory Factor Analysis (EFA) using the psych package in R. Factor scores are then used as outcome variables for machine learning models.

EFA was performed on the training dataset to derive factor weights that were subsequently used to obtain factor scores for participants in the hold-out psychotherapy and pharmacotherapy samples to avoid data leakage. We used the fa() function from the psych package in R, with the minimum residual (minres) estimation method and oblimin rotation, allowing for correlated factors.

First, we computed a Spearman correlation matrix of all 16 QIDS symptoms at baseline that represents the degree of shared variance between every pair of observed variables. Then eigenvalues were computed as the variances of the principal components. For example, an eigenvalue of 3.5 for a factor indicates that it accounts for the equivalent of 3.5 standardized variables' worth of variance, or approximately 21.9% (3.5/16) of the total item variance.
A scree plot (see Supplementary Figure 1) was generated to visualize the point of inflection, or "elbow," beyond which additional factors contributed minimally to explained variance. Based on this and conceptual interpretability, a four-factor solution was selected.

Factors were extracted using the minimum residual (minres) estimation method (fm = "minres"), which seeks the solution that minimizes the residuals between the predicted and observed correlations. The correlation between each symptom and each factor is expressed as a "loading." Symptoms with high loadings on the same factor are understood to represent a shared underlying construct. Loadings above approximately 0.30 are typically considered meaningful.

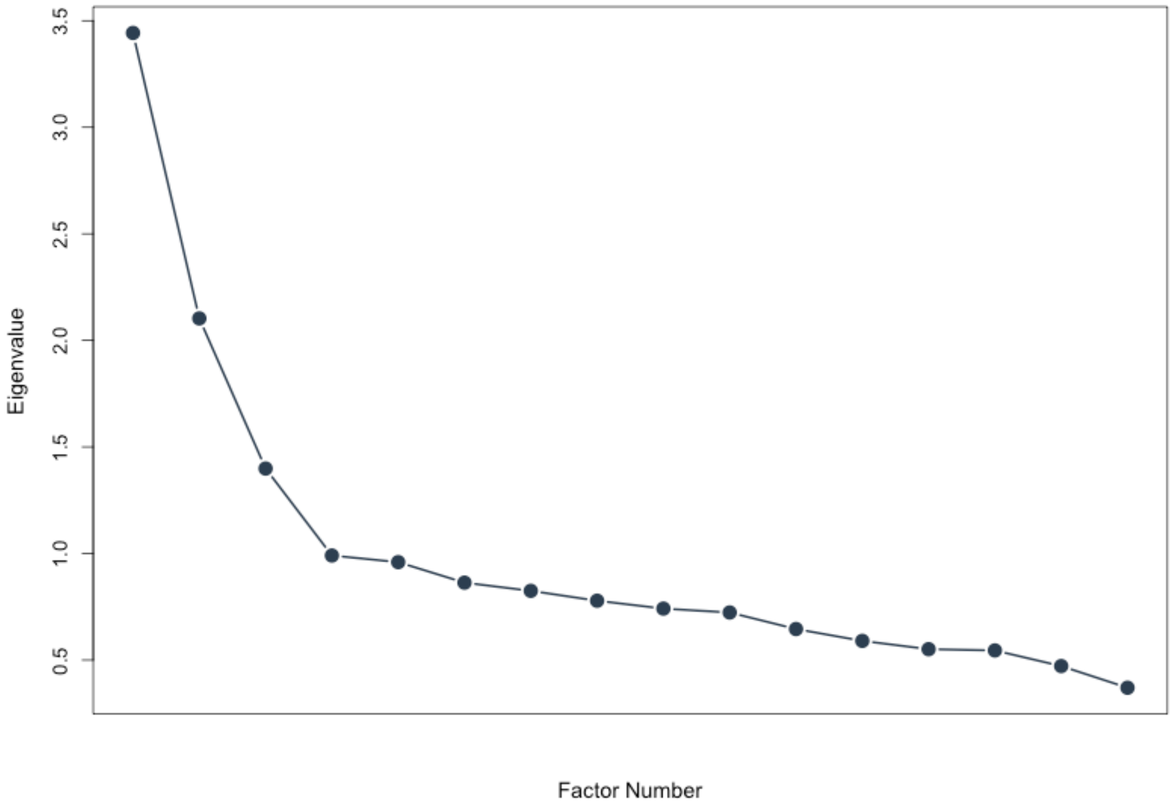


***Supplementary Figure 1.*** *Scree Plot of eigenvalues, on the y axis, showing the variance explained across different factor numbers, on the x axis. Based on the inflection point and conceptual interpretability, a four-factor solution was selected.*

#### Distributions of QIDS Total, Symptom-level and Factor Scores Pre- and Post-intervention

To provide an empirical overview of the post-intervention distributions of our outcome measures for predictive modelling and illustrate the impact of pharmacotherapy and psychotherapy, we visualised the pre- and post-intervention scores for the QIDS total, individual symptoms and latent factors across the full samples changes prior to any stratification. Understanding these distributional characteristics can be important in predictive modelling, as non-normality, skewness, and floor or ceiling effects can impact model performance, bias estimations, and obscure signal in machine learning pipelines (Miotto et al., 2016; Chekroud et al., 2016; Dinga et al., 2020).

Early treatment effects at the 4-week follow up were evident for most scores, even at the individual symptom level (see Score Summary for QIDS total, items and latent factors). The distributions of QIDS total scores were approximately normally distributed, with a shift from moderate to low depression severity post-intervention, indicating a consistent treatment effect across individuals, with few extreme scores. In contrast, the individual symptoms exhibit less symmetrical and more irregular distributions, including skewness and multimodality. The majority of patients reported minimal symptom severity for items related to appetite, weight, insomnia and hypersomnia, psychomotor changes and suicidal ideation, resulting in floor effects with heavily right-skewed distributions and less pronounced post-intervention shifts**.** Other items, such as sad mood, self-critical thoughts or loss of energy and interest, exhibited less skewed distributions with more consistent post-intervention reductions in symptom severity across patients. The latent factors represent distinct symptom clusters resembled that of the total scores. Note that factor scores are standardized around mean 0, as they reflect how far each observation is from the average on that latent factor in standard deviation units.

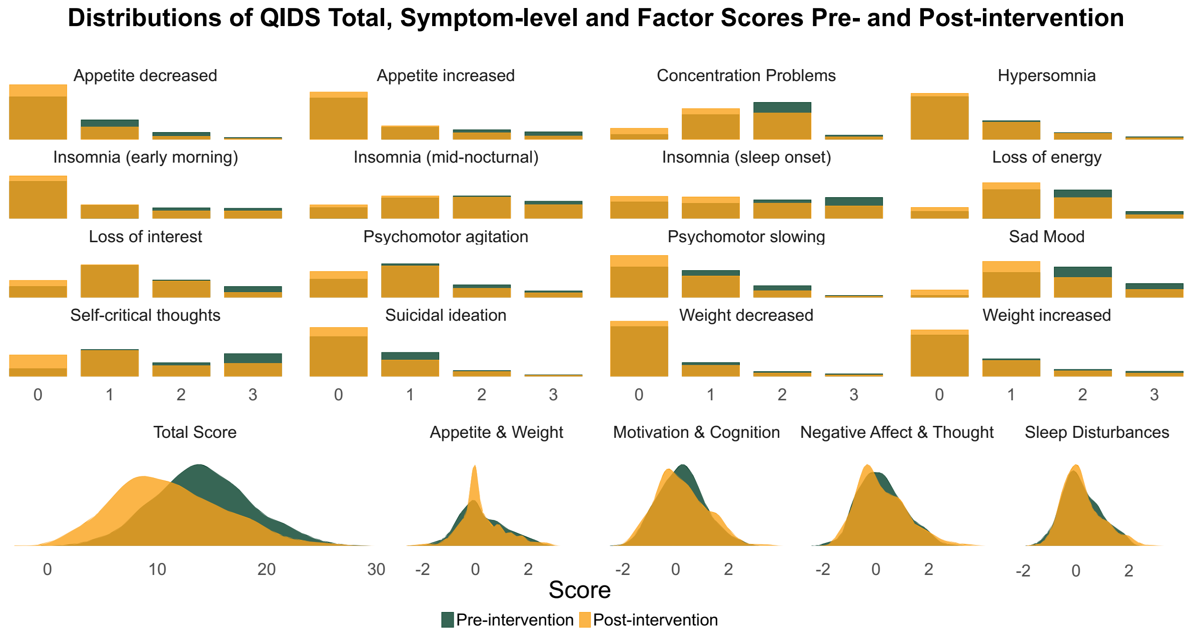


*Supplementary Figure 2. The distributions of QIDS total, symptom-level and factor scores using pre-intervention (dark green) and post-intervention (dark yellow) for the psychotherapy group.*

Total scores were approximately normally distributed, with a clear post-intervention shift from moderate to low depression severity. Individual symptom scores exhibited more irregular, non-normal distributions, including notable skewness and multimodality. Several symptoms—such as appetite and weight changes, insomnia and hypersomnia, psychomotor disturbances, and suicidal ideation—showed strong floor effects, with many participants reporting minimal severity. In contrast, symptoms like sad mood, self-critical thoughts, and loss of energy or interest exhibited less skewed distributions, with more consistent reductions post-intervention. Factor-level scores, across the latent symptom dimensions Appetite & Weight, Motivation & Cognition, Negative Affect & Thought, Sleep Disturbances, showed more symmetric and continuous distributions. Compared to individual symptom items, these composite scores mitigated the effects of item-level skewness and idiosyncratic variation by aggregating across symptoms.

#### Elastic Net Regression Model Performance

To train the models and internally evaluate their performance, we utilized 10-fold nested repeated cross-validation (nrCV). This performed hyperparameter tuning on the inner folds to find the optimal combination of α (alpha), which controls the mix between lasso ($\alpha = 1$) and ridge ($\alpha = 0$) regularization, and λ (lambda), which controls the strength of regularization, using a grid search of 20 evenly spaced values ranging from 0.0001 to 0.9999. Within the training set of each outer fold, 10 inner folds were generated. Within each inner fold, 90% of the data was used to train and tune the model using various hyperparameter configurations. Using the remaining 10% of each inner fold validation set, the best model configuration was selected based on the lowest root mean square error (RMSE). This model was then fit to each of the 10 outer fold training sets and evaluated on each outer validation set; the results were averaged to obtain the cross-validated model performance metrics. As the primary metric to evaluate model performance we used the coefficient of determination ($R^2$) which captures the proportion of variance in the outcome measure explained by the model. It was computed as: $R^2 = 1 - (SSE / SST)$, whereby SSE denotes the sum of squared errors (squared difference between predicted and observed values) and SST the total sum of squares (squared difference between the observed values and their mean).

##### Training Sample (Psychotherapy)

| QIDS Score | Metric | Benchmark Model | Full Model | Delta (Full-Benchmark) |
|---|---|---|---|---|
| **QIDS total** | Correlation Actual to Predicted | 0.563 | 0.592 | 0.029 |
| **QIDS total** | Mean Absolute Error (MAE) | 0.637 | 0.621 | -0.017 |
| **QIDS total** | R squared | 0.352 | 0.393 | 0.041 |
| **QIDS total** | Root Mean Squared Error (RMSE) | 0.804 | 0.779 | -0.026 |

| | | | | |
|---|---|---|---|---|
| **QIDS 1: Insomnia (sleep onset)** | Correlation Actual to Predicted | 0.648 | 0.703 | 0.055 |
| **QIDS 1: Insomnia (sleep onset)** | Mean Absolute Error (MAE) | 0.586 | 0.576 | -0.01 |
| **QIDS 1: Insomnia (sleep onset)** | R squared | 0.472 | 0.488 | 0.016 |
| **QIDS 1: Insomnia (sleep onset)** | Root Mean Squared Error (RMSE) | 0.726 | 0.715 | -0.011 |
| **QIDS 2: Insomnia (mid-nocturnal)** | Correlation Actual to Predicted | 0.525 | 0.542 | 0.017 |
| **QIDS 2: Insomnia (mid-nocturnal)** | Mean Absolute Error (MAE) | 0.670 | 0.686 | 0.016 |
| **QIDS 2: Insomnia (mid-nocturnal)** | R squared | 0.314 | 0.321 | 0.007 |
| **QIDS 2: Insomnia (mid-nocturnal)** | Root Mean Squared Error (RMSE) | 0.828 | 0.823 | -0.004 |
| **QIDS 3: Insomnia (early morning)** | Correlation Actual to Predicted | 0.502 | 0.549 | 0.047 |
| **QIDS 3: Insomnia (early morning)** | Mean Absolute Error (MAE) | 0.633 | 0.635 | 0.001 |
| **QIDS 3: Insomnia (early morning)** | R squared | 0.310 | 0.333 | 0.023 |
| **QIDS 3: Insomnia (early morning)** | Root Mean Squared Error (RMSE) | 0.830 | 0.816 | -0.014 |
| **QIDS 4: Hypersomnia** | Correlation Actual to Predicted | 0.579 | 0.551 | -0.028 |
| **QIDS 4: Hypersomnia** | Mean Absolute Error (MAE) | 0.512 | 0.544 | 0.031 |
| **QIDS 4: Hypersomnia** | R squared | 0.455 | 0.444 | -0.011 |
| **QIDS 4: Hypersomnia** | Root Mean Squared Error (RMSE) | 0.738 | 0.745 | 0.007 |
| **QIDS 5: Sad Mood** | Correlation Actual to Predicted | 0.428 | 0.494 | 0.066 |

| | | | | |
|---|---|---|---|---|
| **QIDS 5: Sad Mood** | Mean Absolute Error (MAE) | 0.680 | 0.683 | 0.002 |
| **QIDS 5: Sad Mood** | R squared | 0.234 | 0.272 | 0.038 |
| **QIDS 5: Sad Mood** | Root Mean Squared Error (RMSE) | 0.874 | 0.852 | -0.022 |
| **QIDS 6: Appetite decreased** | Correlation Actual to Predicted | 0.433 | 0.414 | -0.019 |
| **QIDS 6: Appetite decreased** | Mean Absolute Error (MAE) | 0.606 | 0.620 | 0.014 |
| **QIDS 6: Appetite decreased** | R squared | 0.223 | 0.216 | -0.007 |
| **QIDS 6: Appetite decreased** | Root Mean Squared Error (RMSE) | 0.881 | 0.885 | 0.004 |
| **QIDS 7: Appetite increased** | Correlation Actual to Predicted | 0.407 | 0.412 | 0.006 |
| **QIDS 7: Appetite increased** | Mean Absolute Error (MAE) | 0.616 | 0.635 | 0.019 |
| **QIDS 7: Appetite increased** | R squared | 0.283 | 0.262 | -0.021 |
| **QIDS 7: Appetite increased** | Root Mean Squared Error (RMSE) | 0.846 | 0.858 | 0.012 |
| **QIDS 8: Weight decreased** | Correlation Actual to Predicted | 0.166 | 0.256 | 0.091 |
| **QIDS 8: Weight decreased** | Mean Absolute Error (MAE) | 0.688 | 0.691 | 0.003 |
| **QIDS 8: Weight decreased** | R squared | 0.082 | 0.092 | 0.01 |
| **QIDS 8: Weight decreased** | Root Mean Squared Error (RMSE) | 0.957 | 0.952 | -0.005 |
| **QIDS 9: Weight increased** | Correlation Actual to Predicted | 0.369 | 0.383 | 0.014 |
| **QIDS 9: Weight increased** | Mean Absolute Error (MAE) | 0.689 | 0.685 | -0.004 |
| **QIDS 9: Weight increased** | R squared | 0.175 | 0.194 | 0.019 |

| | | | | |
|---|---|---|---|---|
| **QIDS 9: Weight increased** | Root Mean Squared Error (RMSE) | 0.907 | 0.897 | -0.01 |
| **QIDS 10: Concentration Problems** | Correlation Actual to Predicted | 0.327 | 0.429 | 0.102 |
| **QIDS 10: Concentration Problems** | Mean Absolute Error (MAE) | 0.731 | 0.724 | -0.006 |
| **QIDS 10: Concentration Problems** | R squared | 0.163 | 0.179 | 0.017 |
| **QIDS 10: Concentration Problems** | Root Mean Squared Error (RMSE) | 0.914 | 0.905 | -0.009 |
| **QIDS 11: Self-critical thoughts** | Correlation Actual to Predicted | 0.416 | 0.536 | 0.12 |
| **QIDS 11: Self-critical thoughts** | Mean Absolute Error (MAE) | 0.709 | 0.676 | -0.033 |
| **QIDS 11: Self-critical thoughts** | R squared | 0.232 | 0.296 | 0.064 |
| **QIDS 11: Self-critical thoughts** | Root Mean Squared Error (RMSE) | 0.875 | 0.838 | -0.037 |
| **QIDS 12: Suicidal ideation** | Correlation Actual to Predicted | 0.583 | 0.599 | 0.016 |
| **QIDS 12: Suicidal ideation** | Mean Absolute Error (MAE) | 0.464 | 0.486 | 0.021 |
| **QIDS 12: Suicidal ideation** | R squared | 0.476 | 0.478 | 0.002 |
| **QIDS 12: Suicidal ideation** | Root Mean Squared Error (RMSE) | 0.723 | 0.722 | -0.001 |
| **QIDS 13: Loss of interest** | Correlation Actual to Predicted | 0.357 | 0.454 | 0.097 |
| **QIDS 13: Loss of interest** | Mean Absolute Error (MAE) | 0.699 | 0.709 | 0.009 |
| **QIDS 13: Loss of interest** | R squared | 0.176 | 0.218 | 0.042 |
| **QIDS 13: Loss of interest** | Root Mean Squared Error (RMSE) | 0.907 | 0.883 | -0.023 |

| | | | | |
|---|---|---|---|---|
| **QIDS 14: Loss of energy** | Correlation Actual to Predicted | 0.363 | 0.449 | 0.086 |
| **QIDS 14: Loss of energy** | Mean Absolute Error (MAE) | 0.709 | 0.702 | -0.007 |
| **QIDS 14: Loss of energy** | R squared | 0.173 | 0.206 | 0.034 |
| **QIDS 14: Loss of energy** | Root Mean Squared Error (RMSE) | 0.909 | 0.890 | -0.019 |
| **QIDS 15: Psychomotor slowing** | Correlation Actual to Predicted | 0.455 | 0.511 | 0.056 |
| **QIDS 15: Psychomotor slowing** | Mean Absolute Error (MAE) | 0.690 | 0.681 | -0.008 |
| **QIDS 15: Psychomotor slowing** | R squared | 0.193 | 0.202 | 0.008 |
| **QIDS 15: Psychomotor slowing** | Root Mean Squared Error (RMSE) | 0.897 | 0.893 | -0.005 |
| **QIDS 16: Psychomotor agitation** | Correlation Actual to Predicted | 0.442 | 0.491 | 0.05 |
| **QIDS 16: Psychomotor agitation** | Mean Absolute Error (MAE) | 0.677 | 0.676 | -0.001 |
| **QIDS 16: Psychomotor agitation** | R squared | 0.233 | 0.260 | 0.027 |
| **QIDS 16: Psychomotor agitation** | Root Mean Squared Error (RMSE) | 0.875 | 0.859 | -0.015 |
| **Latent Factor: Motivation & Cognition** | Correlation Actual to Predicted | 0.582 | 0.592 | 0.009 |
| **Latent Factor: Motivation & Cognition** | Mean Absolute Error (MAE) | 0.602 | 0.599 | -0.003 |
| **Latent Factor: Motivation & Cognition** | R squared | 0.354 | 0.376 | 0.022 |
| **Latent Factor: Motivation & Cognition** | Root Mean Squared Error (RMSE) | 0.770 | 0.757 | -0.013 |
| **Latent Factor: Appetite & Weight** | Correlation Actual to Predicted | 0.452 | 0.451 | -0.001 |

| | | | | |
|---|---|---|---|---|
| **Latent Factor: Appetite & Weight** | Mean Absolute Error (MAE) | 0.529 | 0.523 | -0.005 |
| **Latent Factor: Appetite & Weight** | R squared | 0.257 | 0.254 | -0.003 |
| **Latent Factor: Appetite & Weight** | Root Mean Squared Error (RMSE) | 0.706 | 0.708 | 0.001 |
| **Latent Factor: Negative Affect & Thought** | Correlation Actual to Predicted | 0.578 | 0.625 | 0.047 |
| **Latent Factor: Negative Affect & Thought** | Mean Absolute Error (MAE) | 0.570 | 0.553 | -0.017 |
| **Latent Factor: Negative Affect & Thought** | R squared | 0.396 | 0.439 | 0.043 |
| **Latent Factor: Negative Affect & Thought** | Root Mean Squared Error (RMSE) | 0.723 | 0.697 | -0.026 |
| **Latent Factor: Sleep Disturbances** | Correlation Actual to Predicted | 0.622 | 0.615 | -0.006 |
| **Latent Factor: Sleep Disturbances** | Mean Absolute Error (MAE) | 0.437 | 0.441 | 0.003 |
| **Latent Factor: Sleep Disturbances** | R squared | 0.436 | 0.436 | 0 |
| **Latent Factor: Sleep Disturbances** | Root Mean Squared Error (RMSE) | 0.558 | 0.558 | 0 |

*Supplementary Table 2. Model Training Performance in the psychotherapy sample. This table includes various performance metrics that can be used to evaluate the performance of model training such as the correlation of actual to predicted values, mean absolute error (MAE), R squared and root mean squared error (RMSE). These metrics are reported separately for the benchmark model, which included as predictors only the pre-intervention symptom scores of the outcome measure (plus age and sex), as well as the full model, which contains all 85 predictors of interest. The delta denotes the difference value for each metric between the full model and the benchmark model.*

#### Hold-out Validation Sample (Psychotherapy)

| QIDS Score | Metric | Benchmark Model | Full Model | Delta Full-Benchmark) |
|---|---|---|---|---|

| | | | | |
|---|---|---|---|---|
| **QIDS total** | Correlation Actual to Predicted | 0.567 | 0.636 | 0.068 |
| **QIDS total** | Mean Absolute Error (MAE) | 0.602 | 0.590 | -0.013 |
| **QIDS total** | R squared | 0.345 | 0.399 | 0.054 |
| **QIDS total** | R squared Alternative | 0.346 | 0.405 | 0.059 |
| **QIDS total** | Root Mean Squared Error (RMSE) | 0.774 | 0.741 | -0.033 |
| **QIDS 1: Insomnia (sleep onset)** | Correlation Actual to Predicted | 0.562 | 0.564 | 0.002 |
| **QIDS 1: Insomnia (sleep onset)** | Mean Absolute Error (MAE) | 0.645 | 0.640 | -0.005 |
| **QIDS 1: Insomnia (sleep onset)** | R squared | 0.328 | 0.332 | 0.004 |
| **QIDS 1: Insomnia (sleep onset)** | Root Mean Squared Error (RMSE) | 0.797 | 0.795 | -0.003 |
| **QIDS 2: Insomnia (mid-nocturnal)** | Correlation Actual to Predicted | 0.477 | 0.498 | 0.021 |
| **QIDS 2: Insomnia (mid-nocturnal)** | Mean Absolute Error (MAE) | 0.687 | 0.699 | 0.012 |
| **QIDS 2: Insomnia (mid-nocturnal)** | R squared | 0.243 | 0.269 | 0.026 |
| **QIDS 2: Insomnia (mid-nocturnal)** | Root Mean Squared Error (RMSE) | 0.847 | 0.833 | -0.015 |
| **QIDS 3: Insomnia (early morning)** | Correlation Actual to Predicted | 0.431 | 0.475 | 0.044 |
| **QIDS 3: Insomnia (early morning)** | Mean Absolute Error (MAE) | 0.611 | 0.614 | 0.003 |
| **QIDS 3: Insomnia (early morning)** | R squared | 0.245 | 0.258 | 0.013 |
| **QIDS 3: Insomnia (early morning)** | Root Mean Squared Error (RMSE) | 0.819 | 0.812 | -0.007 |

| | | | | |
|---|---|---|---|---|
| **QIDS 4: Hypersomnia** | Correlation Actual to Predicted | 0.597 | 0.562 | -0.034 |
| **QIDS 4: Hypersomnia** | Mean Absolute Error (MAE) | 0.487 | 0.507 | 0.02 |
| **QIDS 4: Hypersomnia** | R squared | 0.442 | 0.438 | -0.004 |
| **QIDS 4: Hypersomnia** | Root Mean Squared Error (RMSE) | 0.717 | 0.719 | 0.002 |
| **QIDS 5: Sad Mood** | Correlation Actual to Predicted | 0.423 | 0.571 | 0.148 |
| **QIDS 5: Sad Mood** | Mean Absolute Error (MAE) | 0.650 | 0.623 | -0.026 |
| **QIDS 5: Sad Mood** | R squared | 0.219 | 0.325 | 0.106 |
| **QIDS 5: Sad Mood** | Root Mean Squared Error (RMSE) | 0.836 | 0.777 | -0.059 |
| **QIDS 6: Appetite decreased** | Correlation Actual to Predicted | 0.397 | 0.385 | -0.012 |
| **QIDS 6: Appetite decreased** | Mean Absolute Error (MAE) | 0.554 | 0.569 | 0.015 |
| **QIDS 6: Appetite decreased** | R squared | 0.266 | 0.243 | -0.023 |
| **QIDS 6: Appetite decreased** | Root Mean Squared Error (RMSE) | 0.824 | 0.836 | 0.013 |
| **QIDS 7: Appetite increased** | Correlation Actual to Predicted | 0.573 | 0.547 | -0.026 |
| **QIDS 7: Appetite increased** | Mean Absolute Error (MAE) | 0.630 | 0.633 | 0.003 |
| **QIDS 7: Appetite increased** | R squared | 0.334 | 0.342 | 0.009 |
| **QIDS 7: Appetite increased** | Root Mean Squared Error (RMSE) | 0.852 | 0.846 | -0.006 |
| **QIDS 8: Weight decreased** | Correlation Actual to Predicted | 0.174 | 0.224 | 0.05 |

| | | | | |
|---|---|---|---|---|
| **QIDS 8: Weight decreased** | Mean Absolute Error (MAE) | 0.640 | 0.632 | -0.007 |
| **QIDS 8: Weight decreased** | R squared | -0.020 | 0.021 | 0.041 |
| **QIDS 8: Weight decreased** | Root Mean Squared Error (RMSE) | 0.873 | 0.856 | -0.018 |
| **QIDS 9: Weight increased** | Correlation Actual to Predicted | 0.467 | 0.476 | 0.009 |
| **QIDS 9: Weight increased** | Mean Absolute Error (MAE) | 0.698 | 0.717 | 0.019 |
| **QIDS 9: Weight increased** | R squared | 0.224 | 0.219 | -0.005 |
| **QIDS 9: Weight increased** | Root Mean Squared Error (RMSE) | 0.943 | 0.945 | 0.003 |
| **QIDS 10: Concentration Problems** | Correlation Actual to Predicted | 0.369 | 0.431 | 0.061 |
| **QIDS 10: Concentration Problems** | Mean Absolute Error (MAE) | 0.753 | 0.740 | -0.013 |
| **QIDS 10: Concentration Problems** | R squared | 0.119 | 0.157 | 0.039 |
| **QIDS 10: Concentration Problems** | Root Mean Squared Error (RMSE) | 0.930 | 0.909 | -0.021 |
| **QIDS 11: Self-critical thoughts** | Correlation Actual to Predicted | 0.543 | 0.577 | 0.034 |
| **QIDS 11: Self-critical thoughts** | Mean Absolute Error (MAE) | 0.674 | 0.652 | -0.023 |
| **QIDS 11: Self-critical thoughts** | R squared | 0.290 | 0.319 | 0.029 |
| **QIDS 11: Self-critical thoughts** | Root Mean Squared Error (RMSE) | 0.822 | 0.805 | -0.017 |
| **QIDS 12: Suicidal ideation** | Correlation Actual to Predicted | 0.538 | 0.594 | 0.056 |

| | | | | |
|---|---|---|---|---|
| **QIDS 12: Suicidal ideation** | Mean Absolute Error (MAE) | 0.555 | 0.561 | 0.006 |
| **QIDS 12: Suicidal ideation** | R squared | 0.336 | 0.359 | 0.022 |
| **QIDS 12: Suicidal ideation** | Root Mean Squared Error (RMSE) | 0.858 | 0.844 | -0.015 |
| **QIDS 13: Loss of interest** | Correlation Actual to Predicted | 0.426 | 0.502 | 0.076 |
| **QIDS 13: Loss of interest** | Mean Absolute Error (MAE) | 0.639 | 0.642 | 0.003 |
| **QIDS 13: Loss of interest** | R squared | 0.168 | 0.248 | 0.079 |
| **QIDS 13: Loss of interest** | Root Mean Squared Error (RMSE) | 0.872 | 0.830 | -0.043 |
| **QIDS 14: Loss of energy** | Correlation Actual to Predicted | 0.352 | 0.402 | 0.051 |
| **QIDS 14: Loss of energy** | Mean Absolute Error (MAE) | 0.740 | 0.722 | -0.018 |
| **QIDS 14: Loss of energy** | R squared | 0.109 | 0.144 | 0.035 |
| **QIDS 14: Loss of energy** | Root Mean Squared Error (RMSE) | 0.947 | 0.928 | -0.019 |
| **QIDS 15: Psychomotor slowing** | Correlation Actual to Predicted | 0.434 | 0.487 | 0.053 |
| **QIDS 15: Psychomotor slowing** | Mean Absolute Error (MAE) | 0.677 | 0.696 | 0.019 |
| **QIDS 15: Psychomotor slowing** | R squared | 0.205 | 0.207 | 0.002 |
| **QIDS 15: Psychomotor slowing** | Root Mean Squared Error (RMSE) | 0.876 | 0.875 | -0.001 |
| **QIDS 16: Psychomotor agitation** | Correlation Actual to Predicted | 0.514 | 0.516 | 0.001 |
| **QIDS 16: Psychomotor agitation** | Mean Absolute Error (MAE) | 0.669 | 0.661 | -0.008 |

| | | | | |
|---|---|---|---|---|
| **QIDS 16: Psychomotor agitation** | R squared | 0.214 | 0.223 | 0.009 |
| **QIDS 16: Psychomotor agitation** | Root Mean Squared Error (RMSE) | 0.860 | 0.855 | -0.005 |
| **Sleep Disturbances** | Correlation Actual to Predicted | 0.561 | 0.577 | 0.016 |
| **Sleep Disturbances** | Mean Absolute Error (MAE) | 0.412 | 0.417 | 0.005 |
| **Sleep Disturbances** | R squared | 0.340 | 0.335 | -0.005 |
| **Sleep Disturbances** | Root Mean Squared Error (RMSE) | 0.569 | 0.571 | 0.002 |
| **Motivation & Cognition** | Correlation Actual to Predicted | 0.547 | 0.593 | 0.046 |
| **Motivation & Cognition** | Mean Absolute Error (MAE) | 0.603 | 0.594 | -0.009 |
| **Motivation & Cognition** | R squared | 0.319 | 0.356 | 0.036 |
| **Motivation & Cognition** | Root Mean Squared Error (RMSE) | 0.774 | 0.753 | -0.021 |
| **Appetite & Weight** | Correlation Actual to Predicted | 0.460 | 0.456 | -0.004 |
| **Appetite & Weight** | Mean Absolute Error (MAE) | 0.507 | 0.506 | -0.001 |
| **Appetite & Weight** | R squared | 0.269 | 0.264 | -0.004 |
| **Appetite & Weight** | Root Mean Squared Error (RMSE) | 0.692 | 0.695 | 0.002 |
| **Negative Affect & Thought** | Correlation Actual to Predicted | 0.606 | 0.661 | 0.055 |
| **Negative Affect & Thought** | Mean Absolute Error (MAE) | 0.556 | 0.546 | -0.01 |
| **Negative Affect & Thought** | R squared | 0.381 | 0.435 | 0.054 |

| | | | | |
|---|---|---|---|---|
| **Negative Affect & Thought** | Root Mean Squared Error (RMSE) | 0.715 | 0.683 | -0.032 |

***Supplementary Table 3. Model Training Performance in the hold-out psychotherapy validation sample****. This table includes various performance metrics that can be used to evaluate the performance of model validation in the hold-out psychotherapy sample such as the correlation of actual to predicted values, mean absolute error (MAE), R squared and root mean squared error (RMSE). These metrics are reported separately for the benchmark model, which included as predictors only the pre-intervention symptom scores of the outcome measure (plus age and sex), as well as the full model, which contains all 85 predictors of interest. The delta denotes the difference value for each metric between the full model and the benchmark model.*

#### Hold-out Validation Sample (Pharmacotherapy)

| QIDS Score | Metric | Benchmark Model | Full Model | Delta Full-Benchmark) |
|---|---|---|---|---|
| **QIDS total** | Correlation Actual to Predicted | 0.431 | 0.467 | 0.036 |
| **QIDS total** | Mean Absolute Error (MAE) | 0.744 | 0.745 | 0.002 |
| **QIDS total** | R squared | 0.181 | 0.221 | 0.04 |
| **QIDS total** | R squared Alternative | 0.216 | 0.255 | 0.038 |
| **QIDS total** | Root Mean Squared Error (RMSE) | 0.940 | 0.917 | -0.023 |
| **QIDS 1: Insomnia (sleep onset)** | Correlation Actual to Predicted | 0.571 | 0.572 | 0 |
| **QIDS 1: Insomnia (sleep onset)** | Mean Absolute Error (MAE) | 0.672 | 0.689 | 0.018 |
| **QIDS 1: Insomnia (sleep onset)** | R squared | 0.348 | 0.324 | -0.024 |
| **QIDS 1: Insomnia (sleep onset)** | Root Mean Squared Error (RMSE) | 0.827 | 0.842 | 0.015 |
| **QIDS 2: Insomnia (mid-nocturnal)** | Correlation Actual to Predicted | 0.330 | 0.330 | 0.001 |

| | | | | |
|---|---|---|---|---|
| **QIDS 2: Insomnia (mid-nocturnal)** | Mean Absolute Error (MAE) | 0.827 | 0.821 | -0.007 |
| **QIDS 2: Insomnia (mid-nocturnal)** | R squared | 0.042 | 0.062 | 0.02 |
| **QIDS 2: Insomnia (mid-nocturnal)** | Root Mean Squared Error (RMSE) | 1.011 | 1.000 | -0.011 |
| **QIDS 3: Insomnia (early morning)** | Correlation Actual to Predicted | 0.492 | 0.510 | 0.017 |
| **QIDS 3: Insomnia (early morning)** | Mean Absolute Error (MAE) | 0.656 | 0.679 | 0.023 |
| **QIDS 3: Insomnia (early morning)** | R squared | 0.325 | 0.309 | -0.016 |
| **QIDS 3: Insomnia (early morning)** | Root Mean Squared Error (RMSE) | 0.837 | 0.847 | 0.01 |
| **QIDS 4: Hypersomnia** | Correlation Actual to Predicted | 0.570 | 0.559 | -0.011 |
| **QIDS 4: Hypersomnia** | Mean Absolute Error (MAE) | 0.621 | 0.650 | 0.029 |
| **QIDS 4: Hypersomnia** | R squared | 0.401 | 0.403 | 0.001 |
| **QIDS 4: Hypersomnia** | Root Mean Squared Error (RMSE) | 0.870 | 0.869 | -0.001 |
| **QIDS 5: Sad Mood** | Correlation Actual to Predicted | 0.380 | 0.406 | 0.026 |
| **QIDS 5: Sad Mood** | Mean Absolute Error (MAE) | 0.824 | 0.828 | 0.004 |
| **QIDS 5: Sad Mood** | R squared | 0.086 | 0.090 | 0.004 |
| **QIDS 5: Sad Mood** | Root Mean Squared Error (RMSE) | 1.000 | 0.997 | -0.002 |
| **QIDS 6: Appetite decreased** | Correlation Actual to Predicted | 0.258 | 0.326 | 0.068 |
| **QIDS 6: Appetite decreased** | Mean Absolute Error (MAE) | 0.862 | 0.852 | -0.01 |

| | | | | |
|---|---|---|---|---|
| **QIDS 6: Appetite decreased** | R squared | -0.005 | 0.058 | 0.063 |
| **QIDS 6: Appetite decreased** | Root Mean Squared Error (RMSE) | 1.105 | 1.070 | -0.035 |
| **QIDS 7: Appetite increased** | Correlation Actual to Predicted | 0.330 | 0.298 | -0.032 |
| **QIDS 7: Appetite increased** | Mean Absolute Error (MAE) | 0.618 | 0.627 | 0.009 |
| **QIDS 7: Appetite increased** | R squared | 0.007 | 0.023 | 0.016 |
| **QIDS 7: Appetite increased** | Root Mean Squared Error (RMSE) | 0.815 | 0.808 | -0.007 |
| **QIDS 8: Weight decreased** | Correlation Actual to Predicted | 0.364 | 0.388 | 0.024 |
| **QIDS 8: Weight decreased** | Mean Absolute Error (MAE) | 0.838 | 0.859 | 0.022 |
| **QIDS 8: Weight decreased** | R squared | 0.061 | 0.056 | -0.005 |
| **QIDS 8: Weight decreased** | Root Mean Squared Error (RMSE) | 1.199 | 1.202 | 0.003 |
| **QIDS 9: Weight increased** | Correlation Actual to Predicted | 0.372 | 0.347 | -0.025 |
| **QIDS 9: Weight increased** | Mean Absolute Error (MAE) | 0.678 | 0.680 | 0.003 |
| **QIDS 9: Weight increased** | R squared | 0.161 | 0.168 | 0.007 |
| **QIDS 9: Weight increased** | Root Mean Squared Error (RMSE) | 0.924 | 0.920 | -0.004 |
| **QIDS 10: Concentration Problems** | Correlation Actual to Predicted | 0.323 | 0.347 | 0.024 |
| **QIDS 10: Concentration Problems** | Mean Absolute Error (MAE) | 0.831 | 0.836 | 0.005 |

| | | | | |
|---|---|---|---|---|
| **QIDS 10: Concentration Problems** | R squared | 0.092 | 0.096 | 0.004 |
| **QIDS 10: Concentration Problems** | Root Mean Squared Error (RMSE) | 1.007 | 1.005 | -0.002 |
| **QIDS 11: Self-critical thoughts** | Correlation Actual to Predicted | 0.295 | 0.472 | 0.177 |
| **QIDS 11: Self-critical thoughts** | Mean Absolute Error (MAE) | 0.848 | 0.792 | -0.057 |
| **QIDS 11: Self-critical thoughts** | R squared | 0.073 | 0.172 | 0.099 |
| **QIDS 11: Self-critical thoughts** | Root Mean Squared Error (RMSE) | 1.001 | 0.946 | -0.055 |
| **QIDS 12: Suicidal ideation** | Correlation Actual to Predicted | 0.474 | 0.521 | 0.047 |
| **QIDS 12: Suicidal ideation** | Mean Absolute Error (MAE) | 0.904 | 0.899 | -0.005 |
| **QIDS 12: Suicidal ideation** | R squared | 0.230 | 0.266 | 0.036 |
| **QIDS 12: Suicidal ideation** | Root Mean Squared Error (RMSE) | 1.186 | 1.158 | -0.028 |
| **QIDS 13: Loss of interest** | Correlation Actual to Predicted | 0.288 | 0.323 | 0.034 |
| **QIDS 13: Loss of interest** | Mean Absolute Error (MAE) | 0.888 | 0.906 | 0.019 |
| **QIDS 13: Loss of interest** | R squared | 0.068 | 0.072 | 0.004 |
| **QIDS 13: Loss of interest** | Root Mean Squared Error (RMSE) | 1.073 | 1.071 | -0.002 |
| **QIDS 14: Loss of energy** | Correlation Actual to Predicted | 0.360 | 0.368 | 0.008 |
| **QIDS 14: Loss of energy** | Mean Absolute Error (MAE) | 0.900 | 0.890 | -0.01 |
| **QIDS 14: Loss of energy** | R squared | 0.075 | 0.074 | 0 |

| | | | | |
|---|---|---|---|---|
| **QIDS 14: Loss of energy** | Root Mean Squared Error (RMSE) | 1.077 | 1.077 | 0 |
| **QIDS 15: Psychomotor slowing** | Correlation Actual to Predicted | 0.468 | 0.448 | -0.02 |
| **QIDS 15: Psychomotor slowing** | Mean Absolute Error (MAE) | 0.737 | 0.767 | 0.03 |
| **QIDS 15: Psychomotor slowing** | R squared | 0.194 | 0.175 | -0.019 |
| **QIDS 15: Psychomotor slowing** | Root Mean Squared Error (RMSE) | 0.953 | 0.964 | 0.011 |
| **QIDS 16: Psychomotor agitation** | Correlation Actual to Predicted | 0.270 | 0.294 | 0.024 |
| **QIDS 16: Psychomotor agitation** | Mean Absolute Error (MAE) | 0.874 | 0.843 | -0.032 |
| **QIDS 16: Psychomotor agitation** | R squared | 0.028 | 0.084 | 0.056 |
| **QIDS 16: Psychomotor agitation** | Root Mean Squared Error (RMSE) | 1.095 | 1.063 | -0.032 |
| **Sleep Disturbances** | Correlation Actual to Predicted | 0.534 | 0.506 | -0.028 |
| **Sleep Disturbances** | Mean Absolute Error (MAE) | 0.529 | 0.536 | 0.007 |
| **Sleep Disturbances** | R squared | 0.329 | 0.280 | -0.049 |
| **Sleep Disturbances** | Root Mean Squared Error (RMSE) | 0.635 | 0.658 | 0.023 |
| **Motivation & Cognition** | Correlation Actual to Predicted | 0.444 | 0.475 | 0.031 |
| **Motivation & Cognition** | Mean Absolute Error (MAE) | 0.768 | 0.735 | -0.033 |
| **Motivation & Cognition** | R squared | 0.156 | 0.195 | 0.038 |
| **Motivation & Cognition** | Root Mean Squared Error (RMSE) | 0.958 | 0.936 | -0.022 |

| | | | | |
|---|---|---|---|---|
| **Appetite & Weight** | Correlation Actual to Predicted | 0.396 | 0.397 | 0.001 |
| **Appetite & Weight** | Mean Absolute Error (MAE) | 0.667 | 0.668 | 0.001 |
| **Appetite & Weight** | R squared | 0.098 | 0.099 | 0.001 |
| **Appetite & Weight** | Root Mean Squared Error (RMSE) | 0.837 | 0.836 | -0.001 |
| **Negative Affect & Thought** | Correlation Actual to Predicted | 0.484 | 0.528 | 0.044 |
| **Negative Affect & Thought** | Mean Absolute Error (MAE) | 0.756 | 0.739 | -0.017 |
| **Negative Affect & Thought** | R squared | 0.212 | 0.278 | 0.066 |
| **Negative Affect & Thought** | Root Mean Squared Error (RMSE) | 0.941 | 0.901 | -0.04 |

***Supplementary Table 4. Model Training Performance in the pharmacotherapy sample***. *This table includes various performance metrics that can be used to evaluate the performance of model validation in the hold-out pharmacotherapy sample such as the correlation of actual to predicted values, mean absolute error (MAE), R squared and root mean squared error (RMSE). These metrics are reported separately for the benchmark model, which included as predictors only the pre-intervention symptom scores of the outcome measure (plus age and sex), as well as the full model, which contains all 85 predictors of interest. The delta denotes the difference value for each metric between the full model and the benchmark model.*

#### Properties of QIDS symptom-level Distributions and Model Performance

To account for the variability across QIDS symptom-level item response distributions when interpreting model training performance, we inspected the relationship between $R^2$, comparing the full to benchmark model, and entropy of the post-intervention scores predicted by the models. For the variability in responses we used a descriptive measure of entropy. Shannon entropy was computed using the entropy() function from the entropy R package to capture the variability of responses across scores. Higher entropy suggests more variability in the distribution, while lower entropy indicates that responses are more skewed or clustered around certain scores.

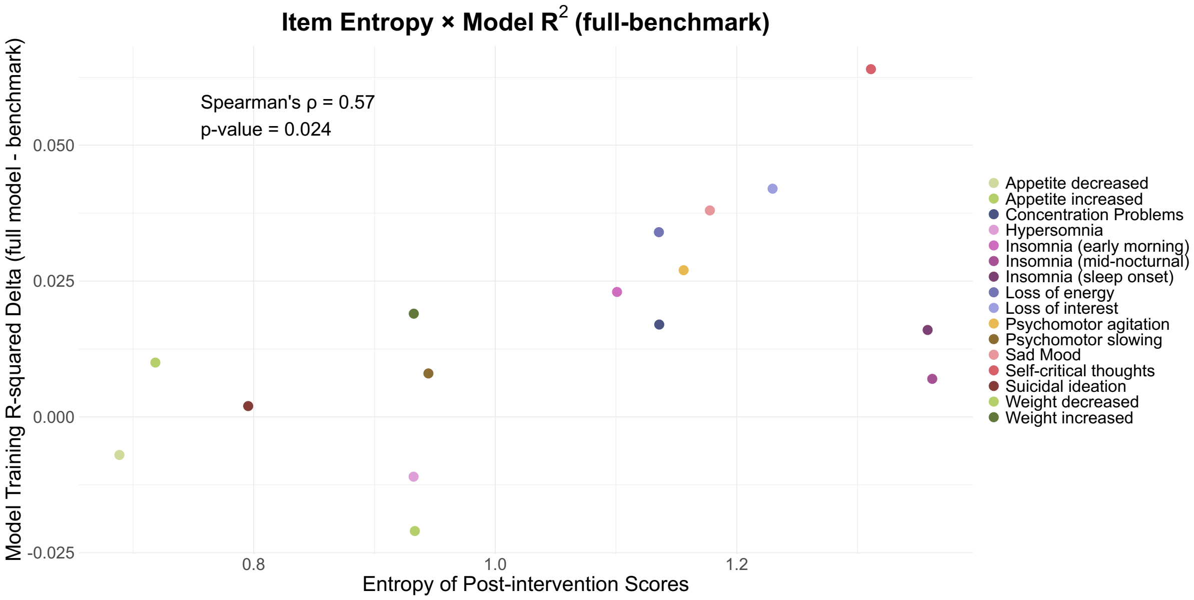


***Supplementary Figure 3. The relationship between item entropy and model training performance.*** *This figure illustrate the relationship between properties of the symptom-level distributions and model training performance which was examined by correlating the delta of* $R^2$ *(full - benchmark model) with item entropy, a measure of distributional spread. Items with more variable score distributions (higher entropy) are associated with greater model prediction improvements.*

We observed a positive correlation between entropy and model performance (Spearman's p=0.57, p-value=0.024), indicating that items with more variability in responses were more accurately predicted. This suggests that more variance to explain in the outcome measure can enhance model discrimination, while imbalanced or rare response items may be less discriminable. When there are stronger ceiling or floor effects, the model may not be able to distinguish between subjects clustered at one end. Meanwhile flatter distributions may yield more reliable predictions due to more consistent relationships between features and outcome scores. As such, entropy can help contextualise the interpretability of model performance variations across QIDS symptom-level scores when comparing the full to benchmark model.

### Model Comparison of Full to Benchmark Model

For each hold-out validation sample, the predicted scores were obtained from both the full and benchmark models for all participants. In each of 1,000 permutations, two permuted models were constructed by randomly selecting predictions from the full and benchmark model for each participant. This procedure ensured that both permuted models contained predictions for the same set of participants, thereby preserving subject-level alignment. The difference in $R^2$ between the two permuted models was calculated at each iteration, generating a null distribution of $R^2$ differences under the assumption that the full and benchmark models perform equivalently. The observed $R^2$ difference between the full and benchmark models was then compared to this null distribution. A p-value was estimated as the proportion of permuted $R^2$ differences that were as extreme or more extreme than the observed $\Delta R^2$. This provided an empirical test of whether the full model offered a statistically significant improvement in predictive performance over the benchmark model.

#### Total Score

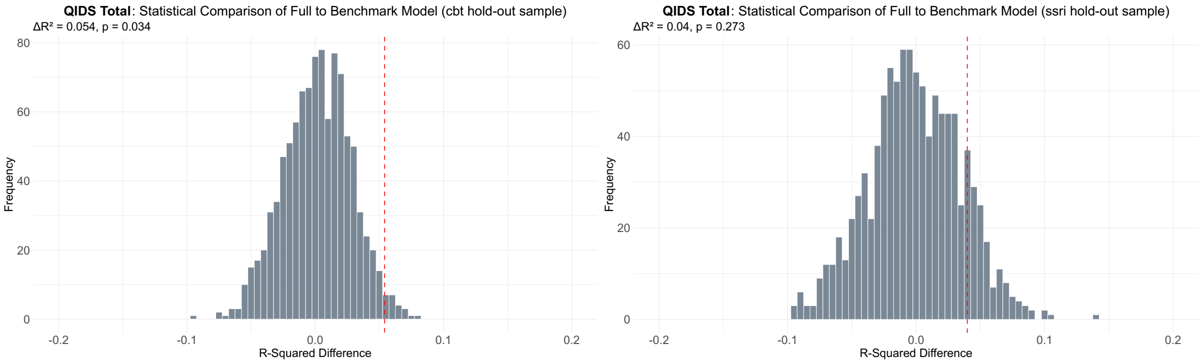


***Supplementary Figure 4. Statistical comparison of the full versus benchmark model for QIDS total scores.*** *This is shown for the psychotherapy (cbt) hold-out sample (left) and pharmacotherapy (ssri) hold-out sample (right). The distributions represent the frequency of R-squared difference values expected by chance based on 1,000 randomized permutations and the red dotted line on the plot indicates the true R-squared difference value between the full and benchmark model.*

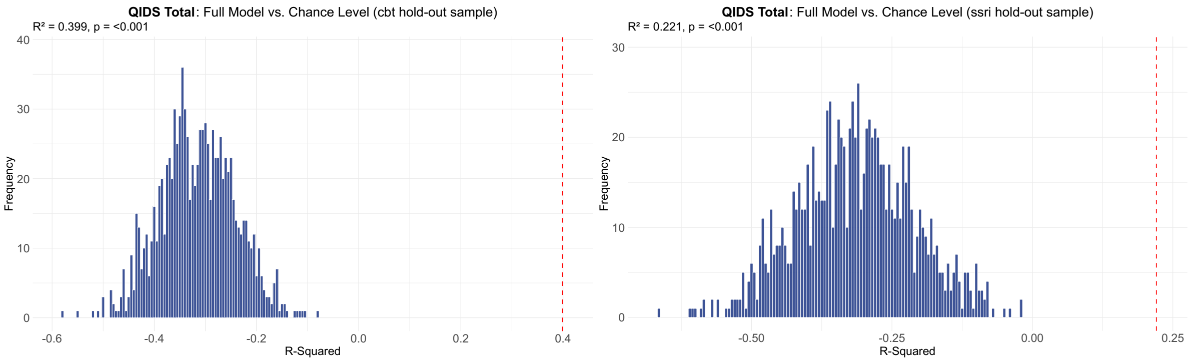


***Supplementary Figure 5. Statistical comparison of the full model versus chance level for QIDS total scores.*** *This is shown for the psychotherapy (cbt) hold-out sample (left) and pharmacotherapy (ssri) hold-out sample (right). The distributions represent the frequency of*

*R-squared values expected by chance based on 1,000 randomized permutations and the red dotted line on the plot indicates the true R-squared value of the full model.*

#### Symptoms

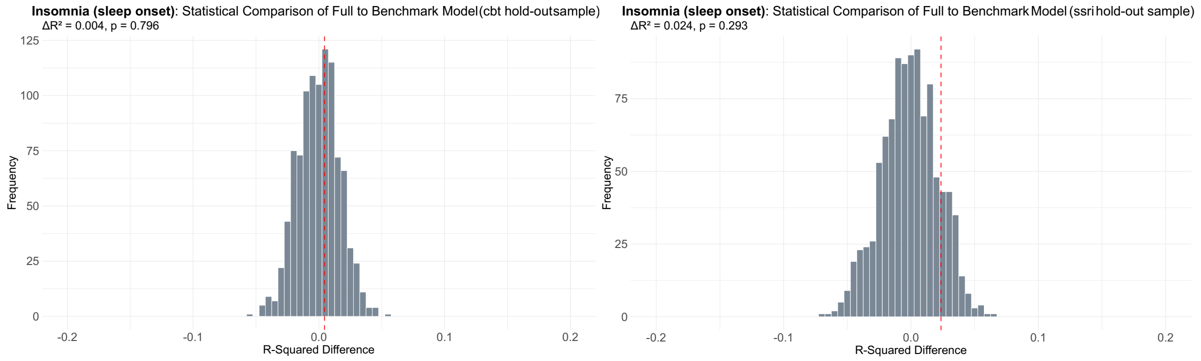


***Supplementary Figure 6. Statistical comparison of the full versus benchmark model for Insomnia (sleep onset) scores.*** *This is shown for the psychotherapy (cbt) hold-out sample (left) and pharmacotherapy (ssri) hold-out sample (right). The distributions represent the frequency of R-squared difference values expected by chance based on 1,000 randomized permutations and the red dotted line on the plot indicates the true R-squared difference value between the full and benchmark model.*

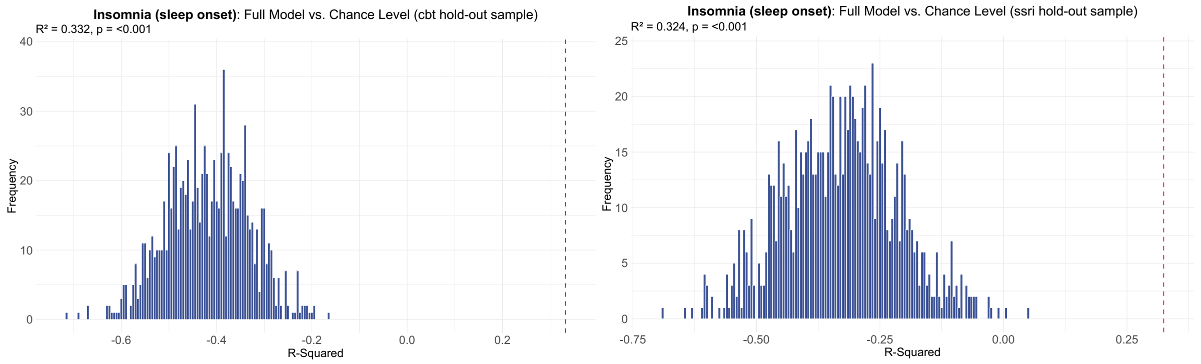


***Supplementary Figure 7. Statistical comparison of the full model versus chance level for Insomnia (sleep onset) scores.*** *This is shown for the psychotherapy (cbt) hold-out sample (left) and pharmacotherapy (ssri) hold-out sample (right). The distributions represent the frequency of R-squared values expected by chance based on 1,000 randomized permutations and the red dotted line on the plot indicates the true R-squared value of the full model.*

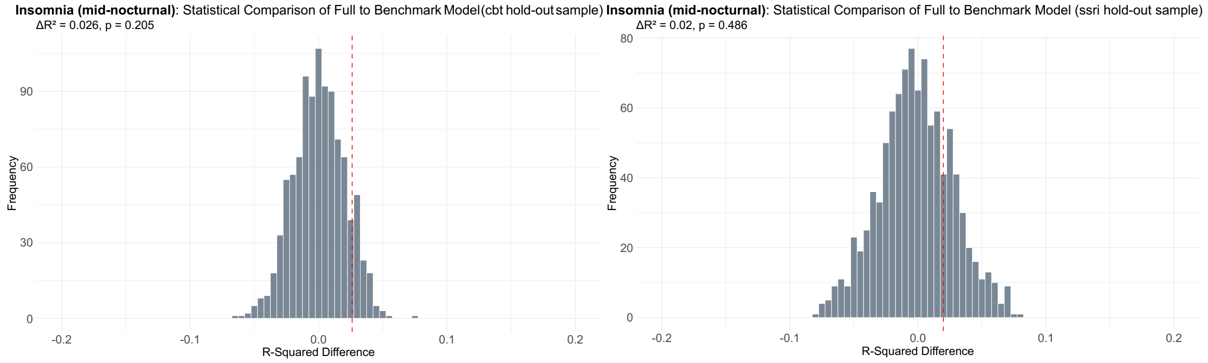

*Supplementary Figure 8. Statistical comparison of the full versus benchmark model for* ***Insomnia (mid-nocturnal) scores.*** *This is shown for the psychotherapy (cbt) hold-out sample (left) and pharmacotherapy (ssri) hold-out sample (right). The distributions represent the frequency of R-squared difference values expected by chance based on 1,000 randomized permutations and the red dotted line on the plot indicates the true R-squared difference value between the full and benchmark model.*

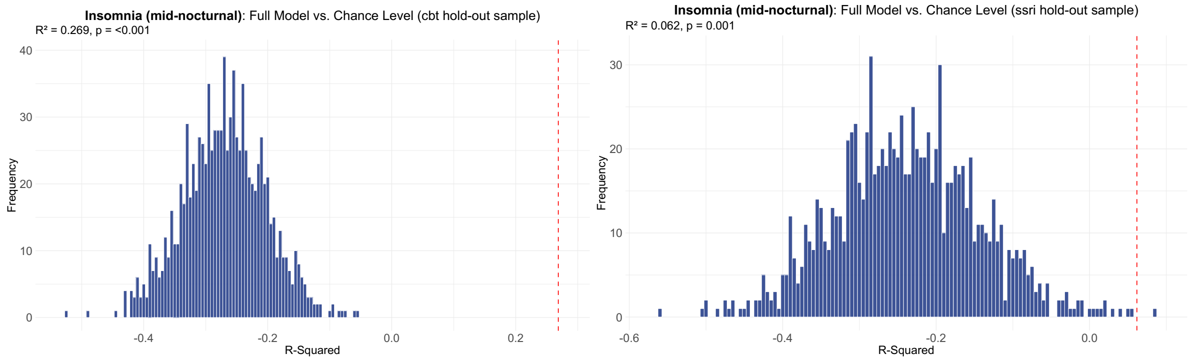


***Supplementary Figure 9. Statistical comparison of the full model versus chance level for Insomnia (mid-nocturnal) scores.*** *This is shown for the psychotherapy (cbt) hold-out sample (left) and pharmacotherapy (ssri) hold-out sample (right). The distributions represent the frequency of R-squared values expected by chance based on 1,000 randomized permutations and the red dotted line on the plot indicates the true R-squared value of the full model.*

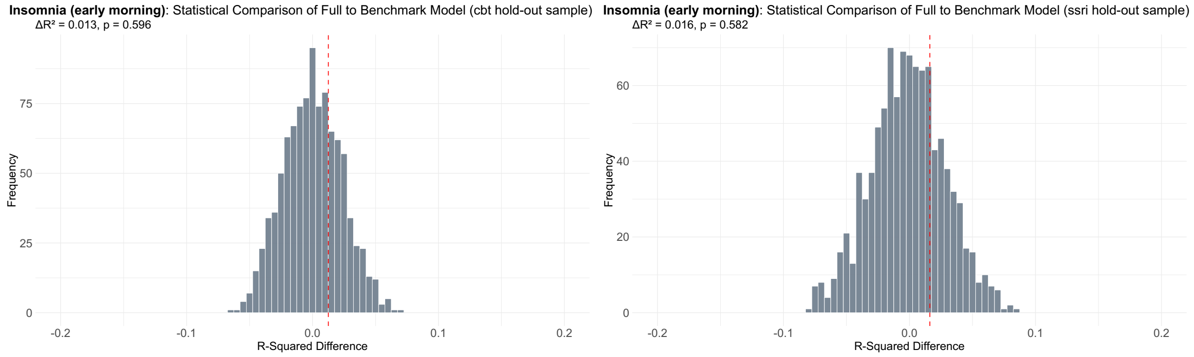


***Supplementary Figure 10. Statistical comparison of the full versus benchmark model for Insomnia (early morning) scores.*** *This is shown for the psychotherapy (cbt) hold-out sample (left) and pharmacotherapy (ssri) hold-out sample (right). The distributions represent the frequency of R-squared difference values expected by chance based on 1,000 randomized permutations and the red dotted line on the plot indicates the true R-squared difference value between the full and benchmark model.*

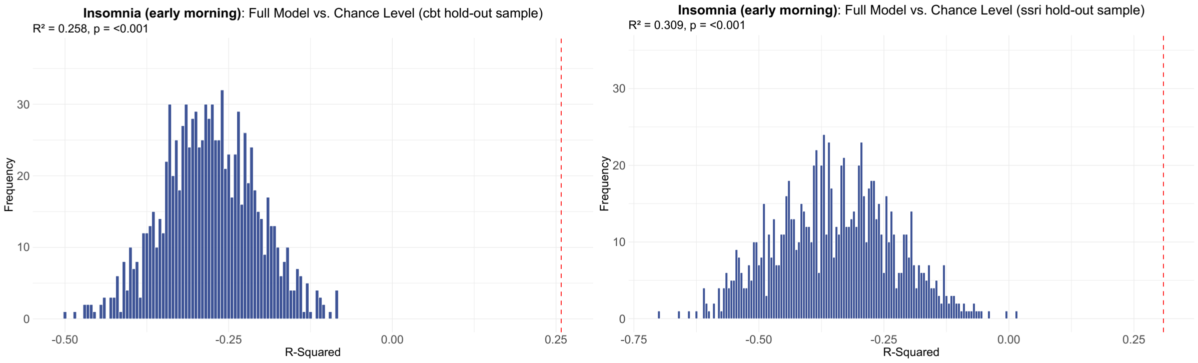


***Supplementary Figure 11. Statistical comparison of the full model versus chance level for Insomnia (early morning) scores.*** *This is shown for the psychotherapy (cbt) hold-out sample (left) and pharmacotherapy (ssri) hold-out sample (right). The distributions represent the frequency of R-squared values expected by chance based on 1,000 randomized permutations and the red dotted line on the plot indicates the true R-squared value of the full model.*

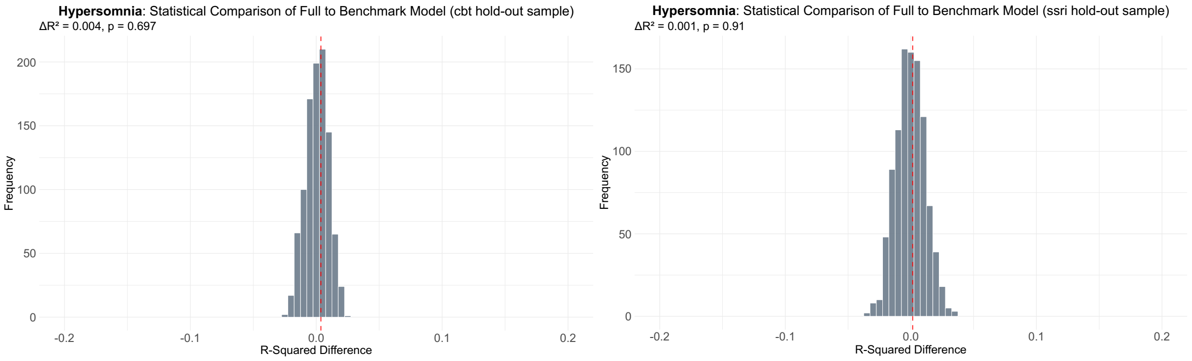


***Supplementary Figure 12. Statistical comparison of the full versus benchmark model for hypersomnia scores.*** *This is shown for the psychotherapy (cbt) hold-out sample (left) and pharmacotherapy (ssri) hold-out sample (right). The distributions represent the frequency of R-squared difference values expected by chance based on 1,000 randomized permutations and the red dotted line on the plot indicates the true R-squared difference value between the full and benchmark model.*

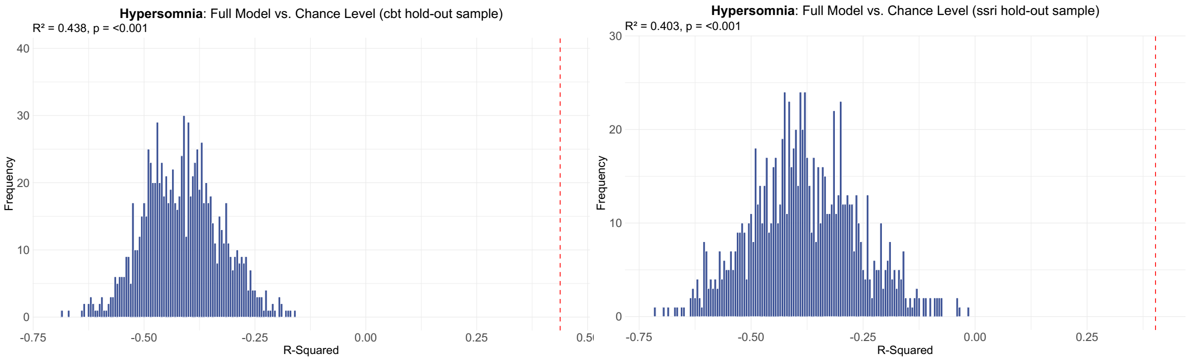


***Supplementary Figure 13. Statistical comparison of the full model versus chance level for hypersomnia scores.*** *This is shown for the psychotherapy (cbt) hold-out sample (left) and pharmacotherapy (ssri) hold-out sample (right). The distributions represent the frequency of R-squared values expected by chance based on 1,000 randomized permutations and the red dotted line on the plot indicates the true R-squared value of the full model.*

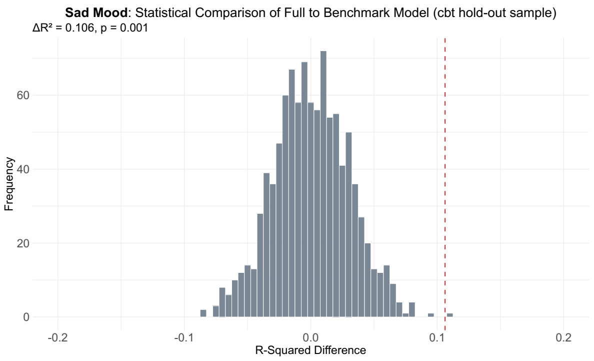


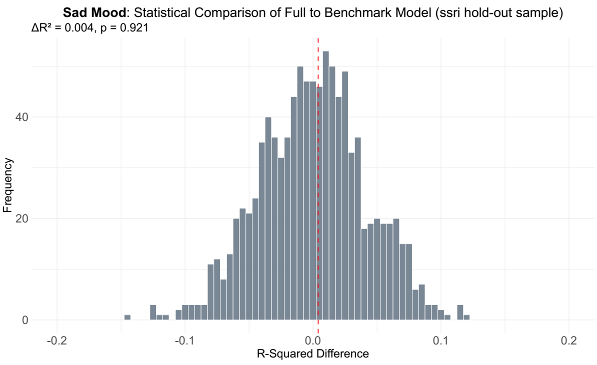


***Supplementary Figure 14. Statistical comparison of the full versus benchmark model for sad mood scores.*** *This is shown for the psychotherapy (cbt) hold-out sample (left) and pharmacotherapy (ssri) hold-out sample (right). The distributions represent the frequency of R-squared difference values expected by chance based on 1,000 randomized permutations and the red dotted line on the plot indicates the true R-squared difference value between the full and benchmark model.*

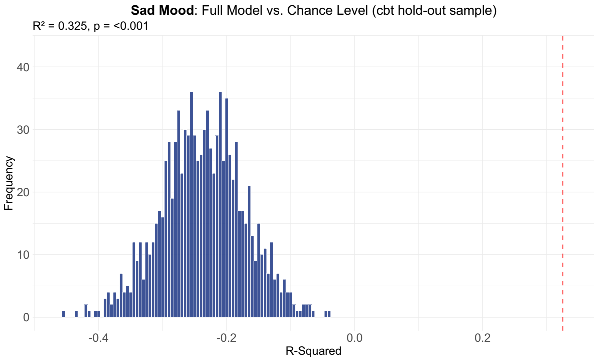


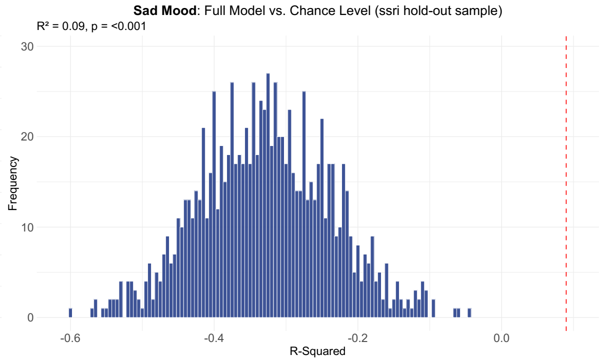


***Supplementary Figure 15. Statistical comparison of the full model versus chance level for sad mood scores.*** *This is shown for the psychotherapy (cbt) hold-out sample (left) and pharmacotherapy (ssri) hold-out sample (right). The distributions represent the frequency of R-squared values expected by chance based on 1,000 randomized permutations and the red dotted line on the plot indicates the true R-squared value of the full model.*

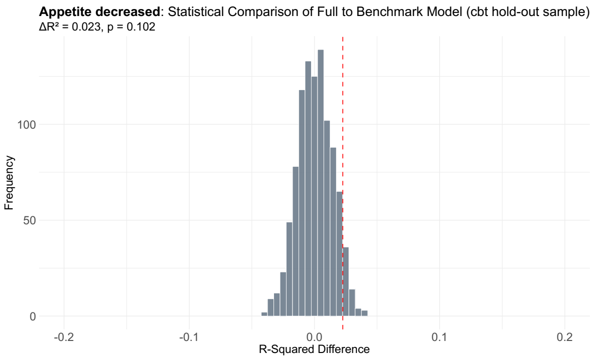


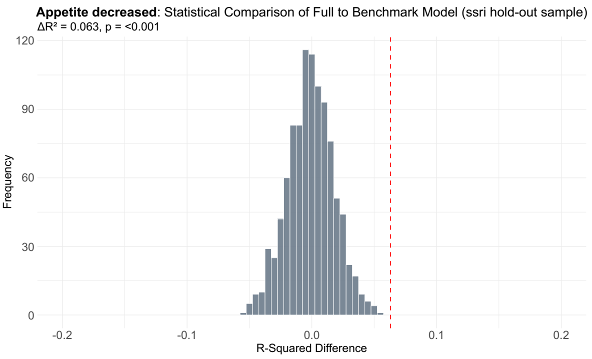


***Supplementary Figure 16. Statistical comparison of the full versus benchmark model for appetite decreased scores.*** *This is shown for the psychotherapy (cbt) hold-out sample (left) and pharmacotherapy (ssri) hold-out sample (right). The distributions represent the frequency of R-squared difference values expected by chance based on 1,000 randomized permutations and the red dotted line on the plot indicates the true R-squared difference value between the full and benchmark model.*

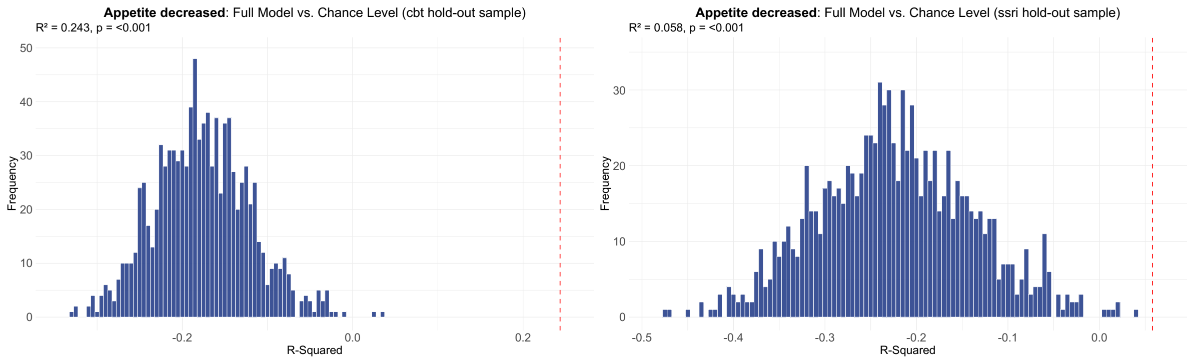


***Supplementary Figure 17. Statistical comparison of the full model versus chance level for appetite decreased scores.*** *This is shown for the psychotherapy (cbt) hold-out sample (left) and pharmacotherapy (ssri) hold-out sample (right). The distributions represent the frequency of R-squared values expected by chance based on 1,000 randomized permutations and the red dotted line on the plot indicates the true R-squared value of the full model.*

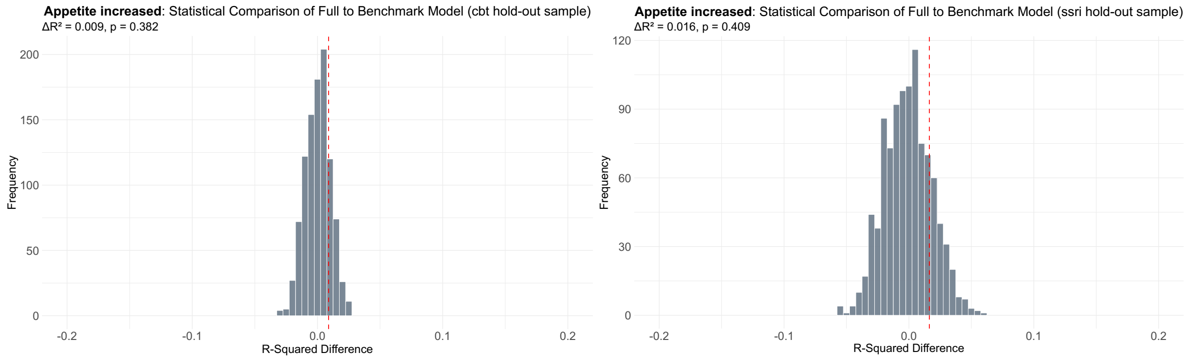


***Supplementary Figure 18. Statistical comparison of the full versus benchmark model for appetite increased scores.*** *This is shown for the psychotherapy (cbt) hold-out sample (left) and pharmacotherapy (ssri) hold-out sample (right). The distributions represent the frequency of R-squared difference values expected by chance based on 1,000 randomized permutations and the red dotted line on the plot indicates the true R-squared difference value between the full and benchmark model.*

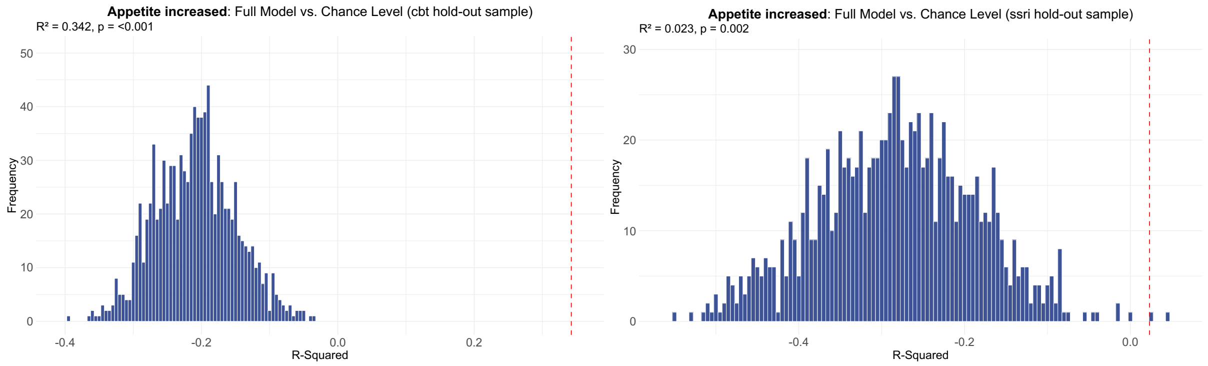


***Supplementary Figure 19. Statistical comparison of the full model versus chance level for appetite increased scores.*** *This is shown for the psychotherapy (cbt) hold-out sample (left) and pharmacotherapy (ssri) hold-out sample (right). The distributions represent the frequency of R-squared values expected by chance based on 1,000 randomized permutations and the red dotted line on the plot indicates the true R-squared value of the full model.*

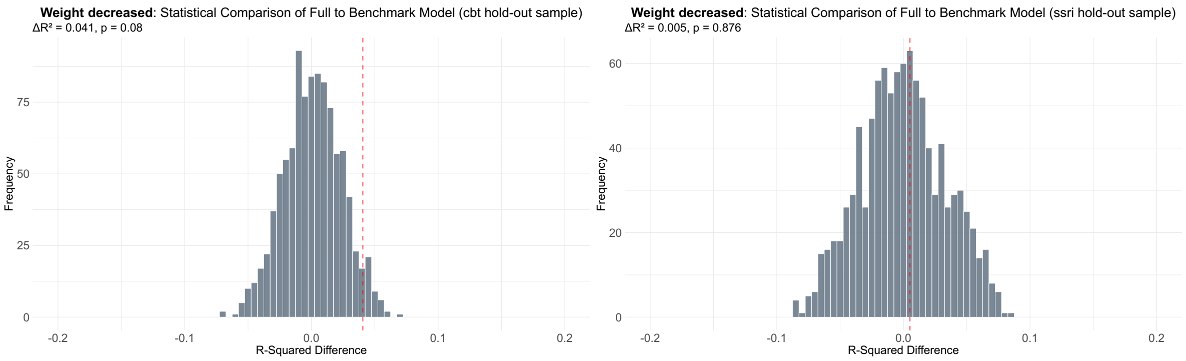


***Supplementary Figure 20. Statistical comparison of the full versus benchmark model for weight decreased scores.*** *This is shown for the psychotherapy (cbt) hold-out sample (left) and pharmacotherapy (ssri) hold-out sample (right). The distributions represent the frequency of R-squared difference values expected by chance based on 1,000 randomized permutations and the red dotted line on the plot indicates the true R-squared difference value between the full and benchmark model.*

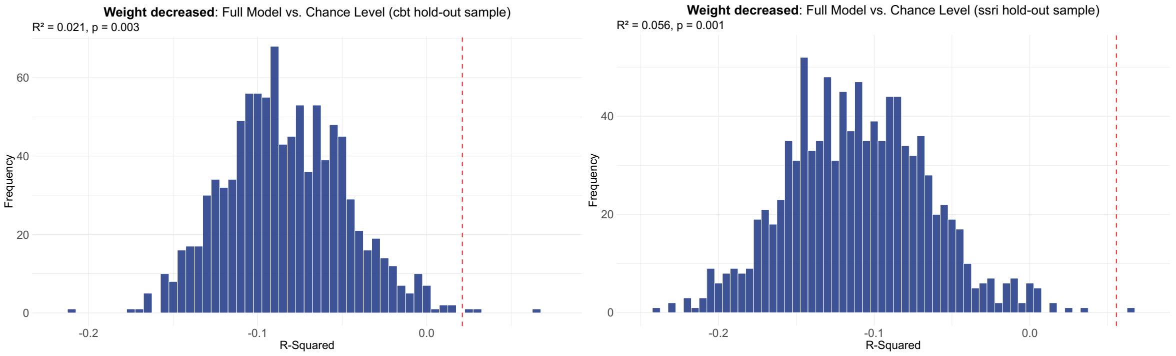


***Supplementary Figure 21. Statistical comparison of the full model versus chance level for weight decreased scores.*** *This is shown for the psychotherapy (cbt) hold-out sample (left) and pharmacotherapy (ssri) hold-out sample (right). The distributions represent the frequency of R-squared values expected by chance based on 1,000 randomized permutations and the red dotted line on the plot indicates the true R-squared value of the full model.*

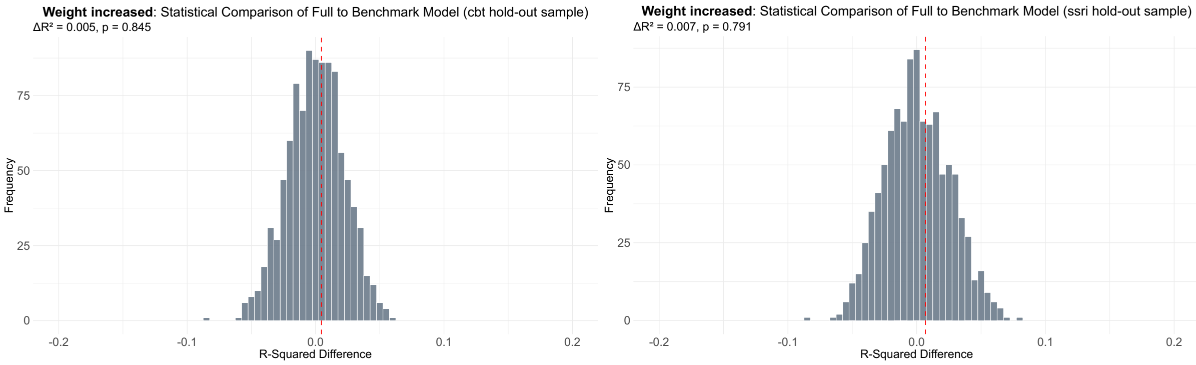


***Supplementary Figure 22. Statistical comparison of the full versus benchmark model for weight increased scores.*** *This is shown for the psychotherapy (cbt) hold-out sample (left) and pharmacotherapy (ssri) hold-out sample (right). The distributions represent the frequency of R-squared difference values expected by chance based on 1,000 randomized permutations and the red dotted line on the plot indicates the true R-squared difference value between the full and benchmark model.*

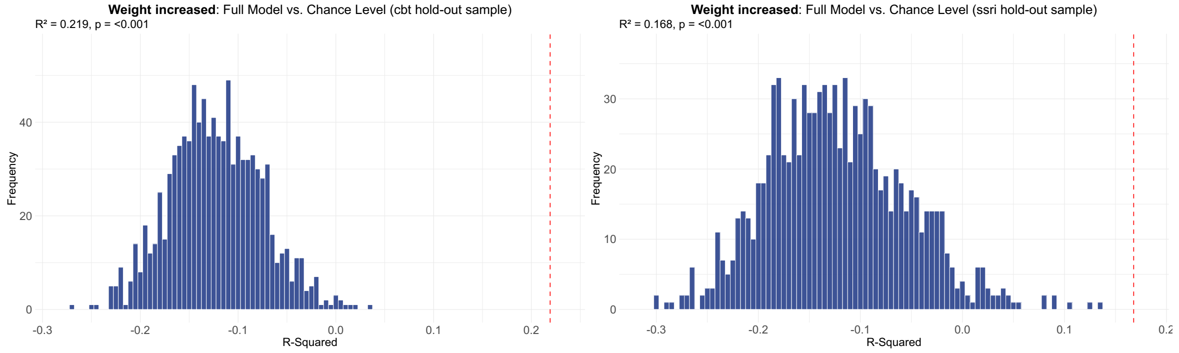


***Supplementary Figure 23. Statistical comparison of the full model versus chance level for weight increased scores.** This is shown for the psychotherapy (cbt) hold-out sample (left) and pharmacotherapy (ssri) hold-out sample (right). The distributions represent the frequency of R-squared values expected by chance based on 1,000 randomized permutations and the red dotted line on the plot indicates the true R-squared value of the full model.*

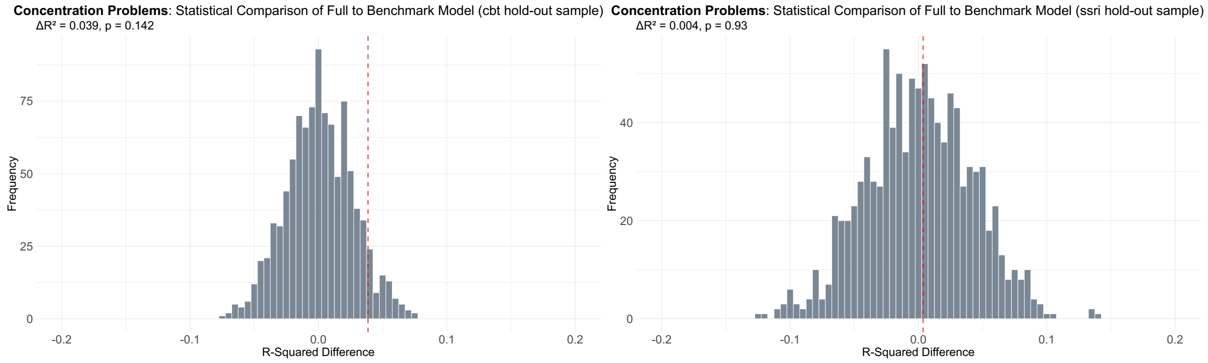


***Supplementary Figure 24. Statistical comparison of the full versus benchmark model for concentration problems scores.** This is shown for the psychotherapy (cbt) hold-out sample (left) and pharmacotherapy (ssri) hold-out sample (right). The distributions represent the frequency of R-squared difference values expected by chance based on 1,000 randomized permutations and the red dotted line on the plot indicates the true R-squared difference value between the full and benchmark model.*

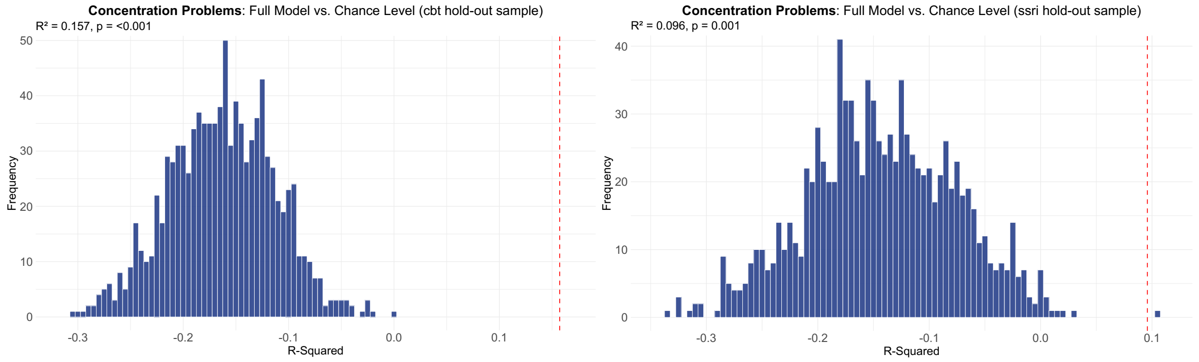


***Supplementary Figure 25. Statistical comparison of the full model versus chance level for concentration problems scores.** This is shown for the psychotherapy (cbt) hold-out sample (left) and pharmacotherapy (ssri) hold-out sample (right). The distributions represent the frequency of R-squared values expected by chance based on 1,000 randomized permutations and the red dotted line on the plot indicates the true R-squared value of the full model.*

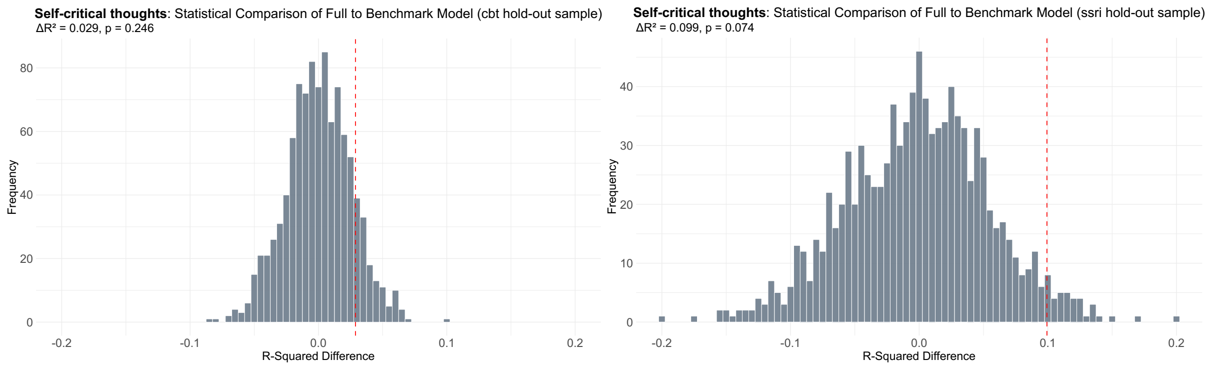


*Supplementary Figure 26. **Statistical comparison of the full versus benchmark model for self-critical thoughts scores.** This is shown for the psychotherapy (cbt) hold-out sample (left) and pharmacotherapy (ssri) hold-out sample (right). The distributions represent the frequency of R-squared difference values expected by chance based on 1,000 randomized permutations and the red dotted line on the plot indicates the true R-squared difference value between the full and benchmark model.*

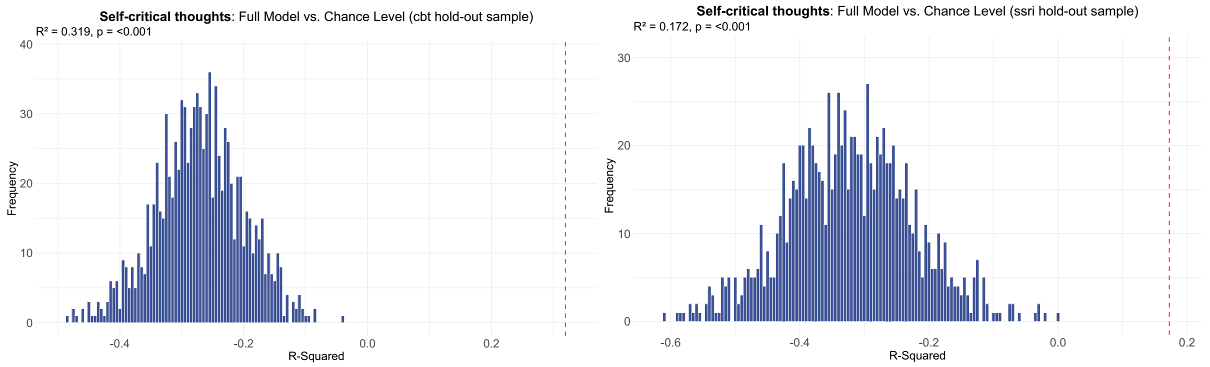


*Supplementary Figure 27. **Statistical comparison of the full model versus chance level for concentration problems scores.** This is shown for the psychotherapy (cbt) hold-out sample (left) and pharmacotherapy (ssri) hold-out sample (right). The distributions represent the frequency of R-squared values expected by chance based on 1,000 randomized permutations and the red dotted line on the plot indicates the true R-squared value of the full model.*

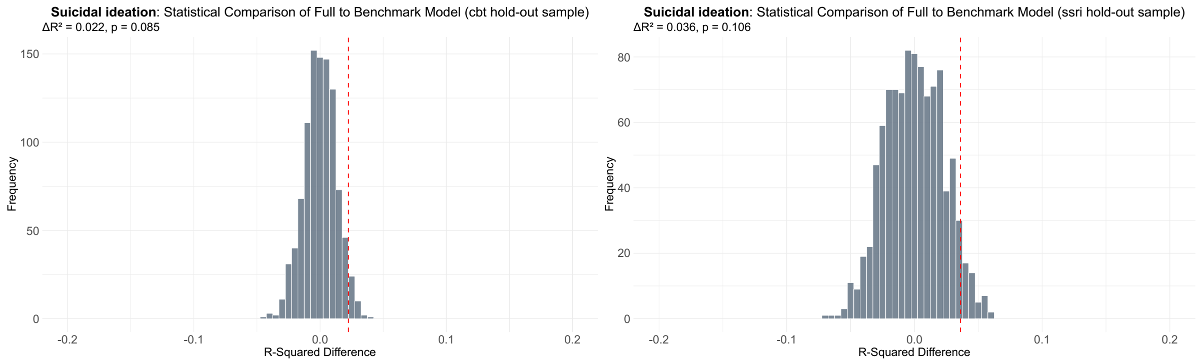


*Supplementary Figure 28. **Statistical comparison of the full versus benchmark model for suicidal ideation scores.** This is shown for the psychotherapy (cbt) hold-out sample (left) and pharmacotherapy (ssri) hold-out sample (right). The distributions represent the frequency of R-squared difference values expected by chance based on 1,000 randomized permutations and the red dotted line on the plot indicates the true R-squared difference value between the full and benchmark model.*

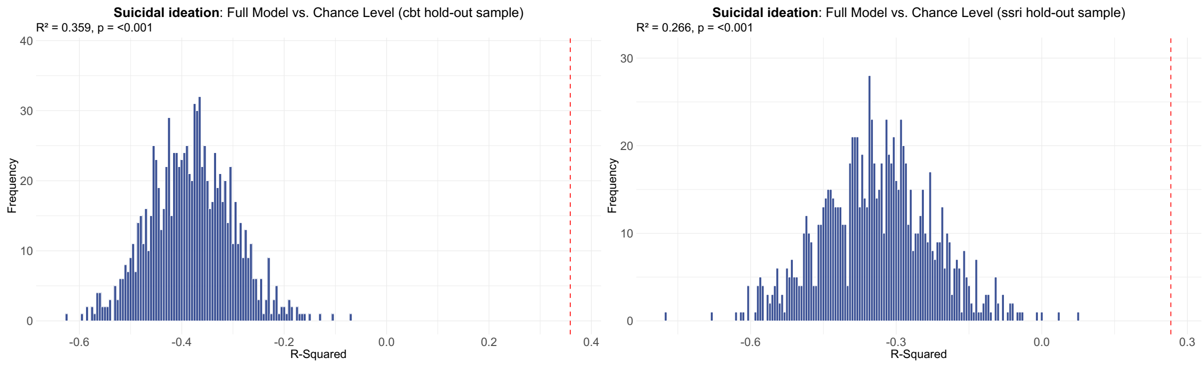


***Supplementary Figure 29. Statistical comparison of the full model versus chance level for suicidal ideation scores.*** *This is shown for the psychotherapy (cbt) hold-out sample (left) and pharmacotherapy (ssri) hold-out sample (right). The distributions represent the frequency of R-squared values expected by chance based on 1,000 randomized permutations and the red dotted line on the plot indicates the true R-squared value of the full model.*

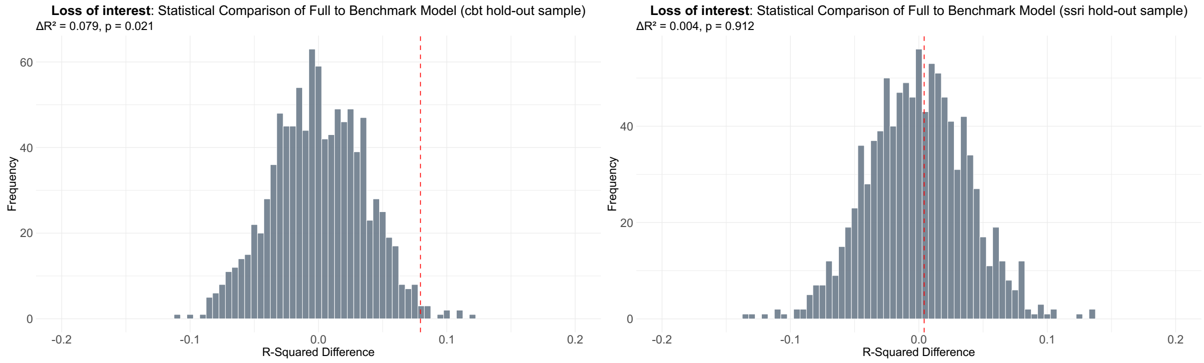


***Supplementary Figure 30. Statistical comparison of the full versus benchmark model for loss of interest scores.*** *This is shown for the psychotherapy (cbt) hold-out sample (left) and pharmacotherapy (ssri) hold-out sample (right). The distributions represent the frequency of R-squared difference values expected by chance based on 1,000 randomized permutations and the red dotted line on the plot indicates the true R-squared difference value between the full and benchmark model.*

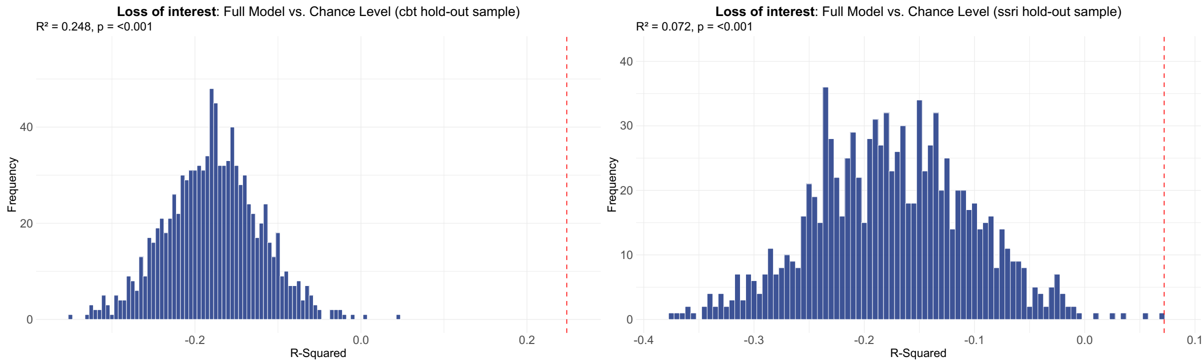


***Supplementary Figure 31. Statistical comparison of the full model versus chance level for loss of interest scores.*** *This is shown for the psychotherapy (cbt) hold-out sample (left) and pharmacotherapy (ssri) hold-out sample (right). The distributions represent the frequency of R-squared values expected by chance based on 1,000 randomized permutations and the red dotted line on the plot indicates the true R-squared value of the full model.*

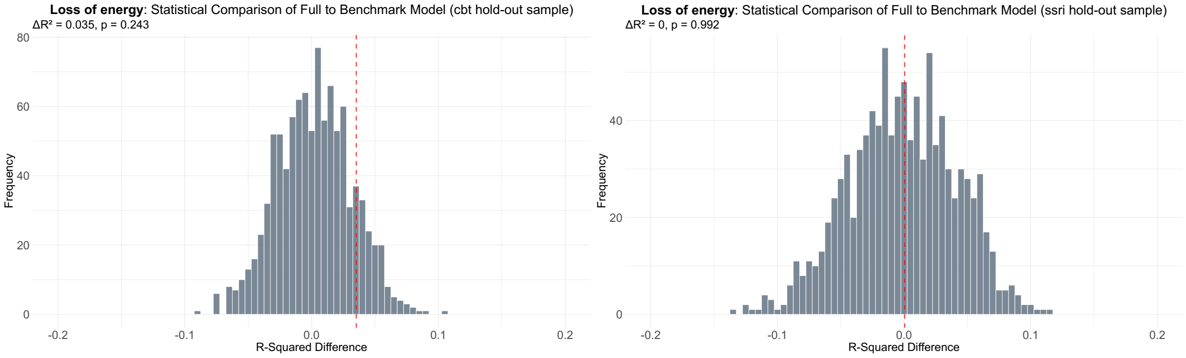


***Supplementary Figure 32. Statistical comparison of the full versus benchmark model for loss of energy scores.*** *This is shown for the psychotherapy (cbt) hold-out sample (left) and pharmacotherapy (ssri) hold-out sample (right). The distributions represent the frequency of R-squared difference values expected by chance based on 1,000 randomized permutations and the red dotted line on the plot indicates the true R-squared difference value between the full and benchmark model.*

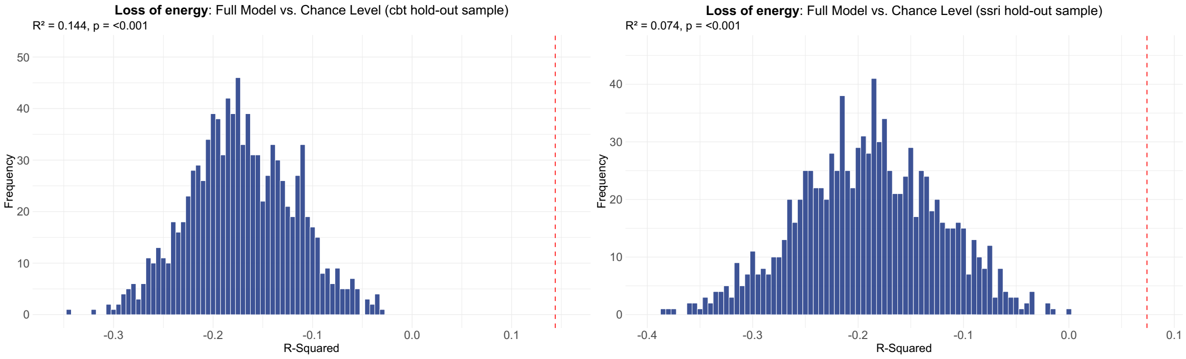


***Supplementary Figure 33. Statistical comparison of the full model versus chance level for loss of energy scores.*** *This is shown for the psychotherapy (cbt) hold-out sample (left) and pharmacotherapy (ssri) hold-out sample (right). The distributions represent the frequency of R-squared values expected by chance based on 1,000 randomized permutations and the red dotted line on the plot indicates the true R-squared value of the full model.*

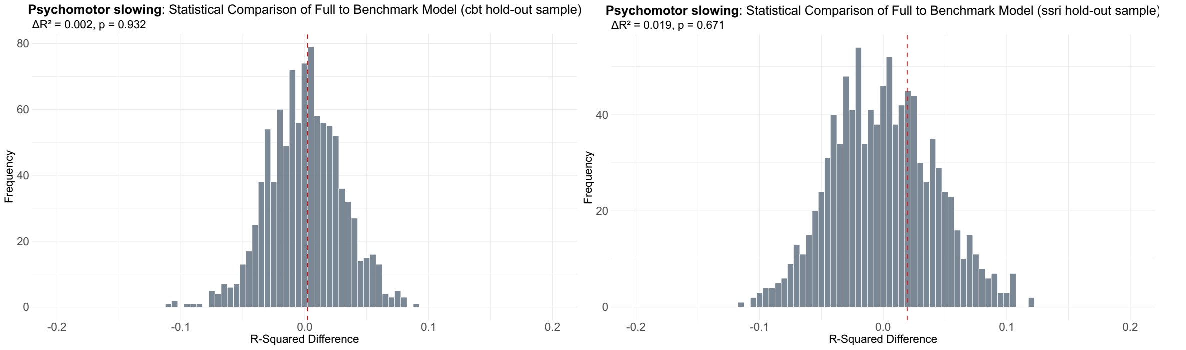


***Supplementary Figure 34. Statistical comparison of the full versus benchmark model for psychomotor slowing scores.*** *This is shown for the psychotherapy (cbt) hold-out sample (left) and pharmacotherapy (ssri) hold-out sample (right). The distributions represent the frequency of R-squared difference values expected by chance based on 1,000 randomized permutations and the red dotted line on the plot indicates the true R-squared difference value between the full and benchmark model.*

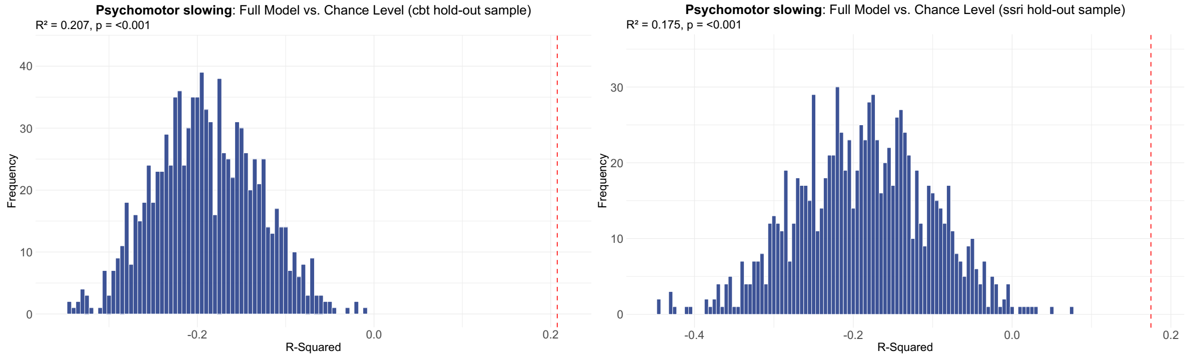


***Supplementary Figure 35. Statistical comparison of the full model versus chance level for psychomotor slowing scores.*** *This is shown for the psychotherapy (cbt) hold-out sample (left) and pharmacotherapy (ssri) hold-out sample (right). The distributions represent the frequency of R-squared values expected by chance based on 1,000 randomized permutations and the red dotted line on the plot indicates the true R-squared value of the full model.*

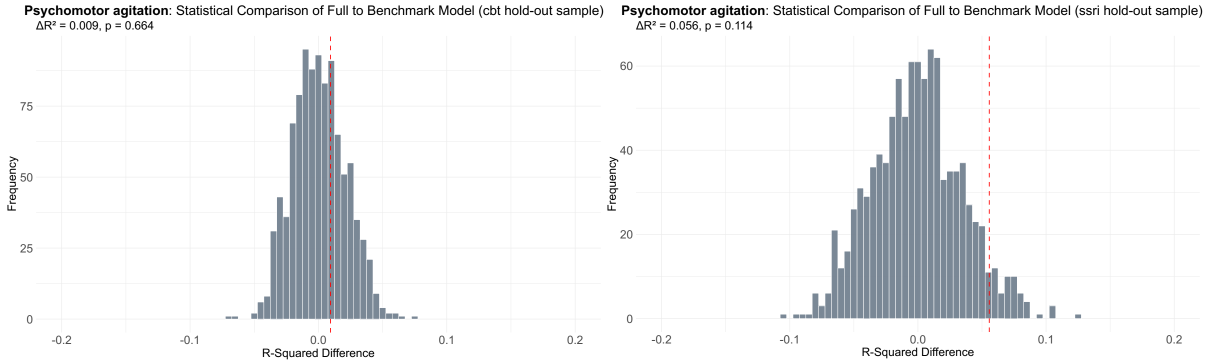


***Supplementary Figure 36. Statistical comparison of the full versus benchmark model for psychomotor agitation scores.*** *This is shown for the psychotherapy (cbt) hold-out sample (left) and pharmacotherapy (ssri) hold-out sample (right). The distributions represent the frequency of R-squared difference values expected by chance based on 1,000 randomized permutations and the red dotted line on the plot indicates the true R-squared difference value between the full and benchmark model.*

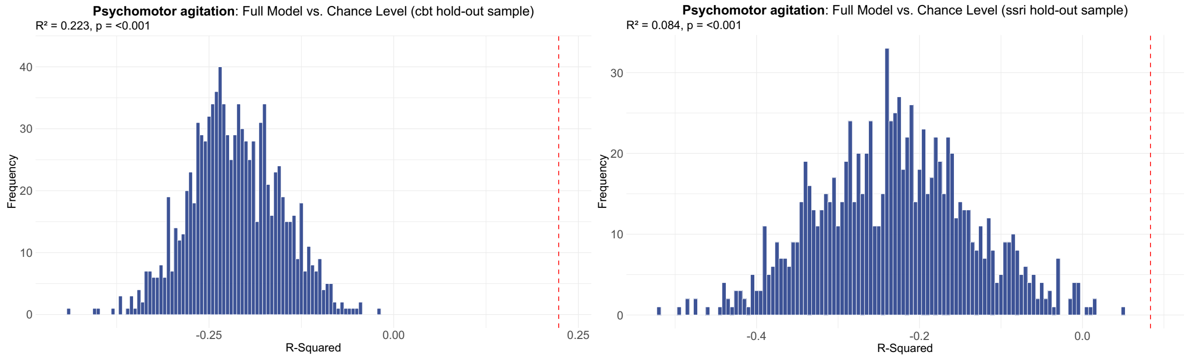


***Supplementary Figure 37. Statistical comparison of the full model versus chance level for psychomotor agitation scores.*** *This is shown for the psychotherapy (cbt) hold-out sample (left) and pharmacotherapy (ssri) hold-out sample (right). The distributions represent the frequency of R-squared values expected by chance based on 1,000 randomized permutations and the red dotted line on the plot indicates the true R-squared value of the full model.*

#### Factors

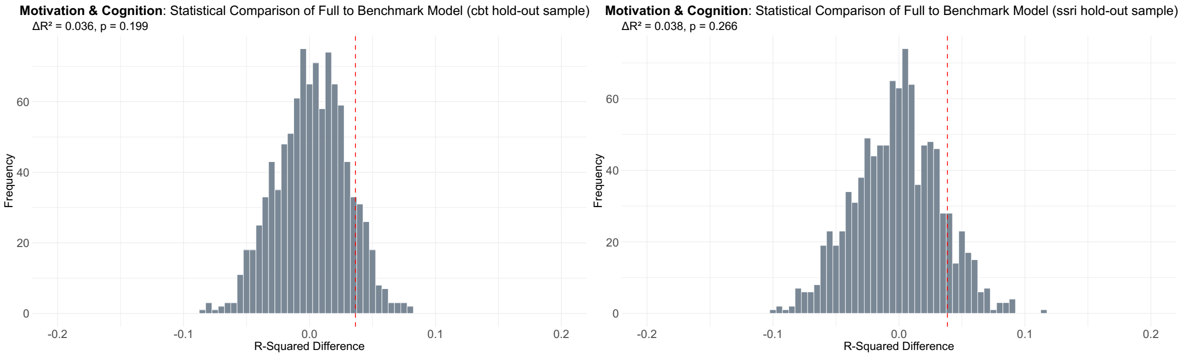


***Supplementary Figure 38. Statistical comparison of the full versus benchmark model for motivation and cognition factor scores.*** *This is shown for the psychotherapy (cbt) hold-out sample (left) and pharmacotherapy (ssri) hold-out sample (right). The distributions represent the frequency of R-squared difference values expected by chance based on 1,000 randomized permutations and the red dotted line on the plot indicates the true R-squared difference value between the full and benchmark model.*

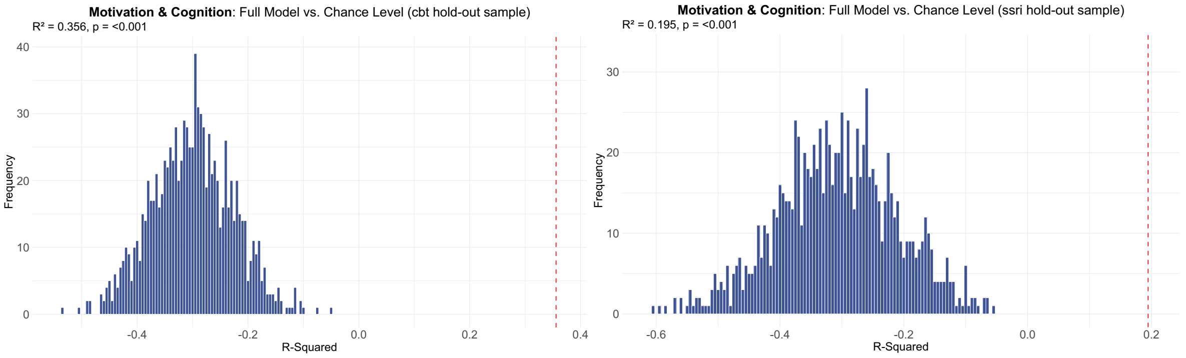


***Supplementary Figure 39. Statistical comparison of the full model versus chance level for motivation and cognition factor scores.*** *This is shown for the psychotherapy (cbt) hold-out sample (left) and pharmacotherapy (ssri) hold-out sample (right). The distributions represent the frequency of R-squared values expected by chance based on 1,000 randomized permutations and the red dotted line on the plot indicates the true R-squared value of the full model.*

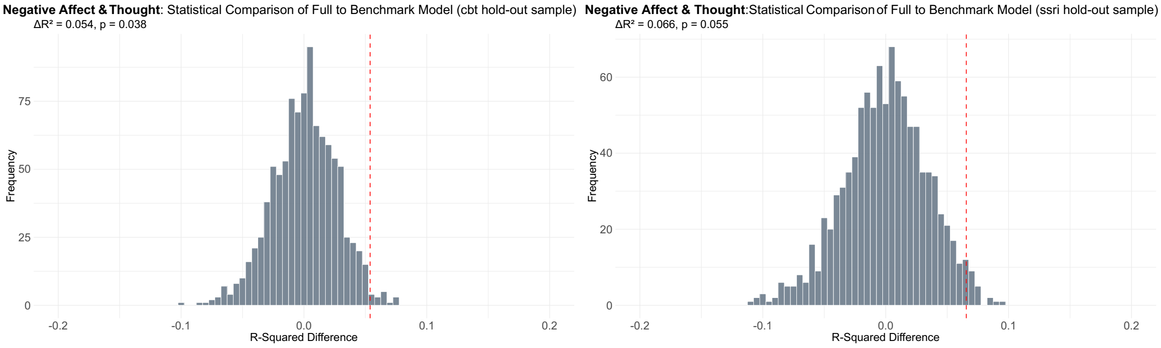


***Supplementary Figure 40. Statistical comparison of the full versus benchmark model for negative affect and thought factor scores.*** *This is shown for the psychotherapy (cbt) hold-out sample (left) and pharmacotherapy (ssri) hold-out sample (right). The distributions represent the frequency of R-squared difference values expected by chance based on 1,000*

*randomized permutations and the red dotted line on the plot indicates the true R-squared difference value between the full and benchmark model.*

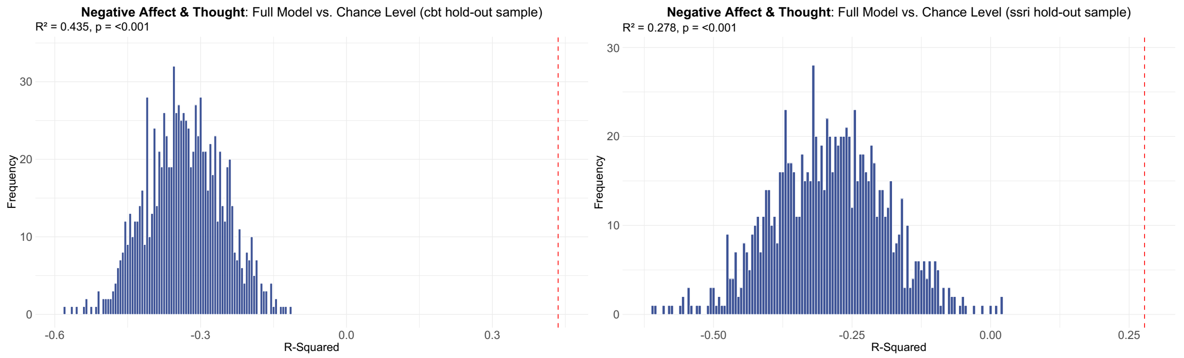


***Supplementary Figure 41. Statistical comparison of the full model versus chance level for negative affect and thought factor scores.*** *This is shown for the psychotherapy (cbt) hold-out sample (left) and pharmacotherapy (ssri) hold-out sample (right). The distributions represent the frequency of R-squared values expected by chance based on 1,000 randomized permutations and the red dotted line on the plot indicates the true R-squared value of the full model.*

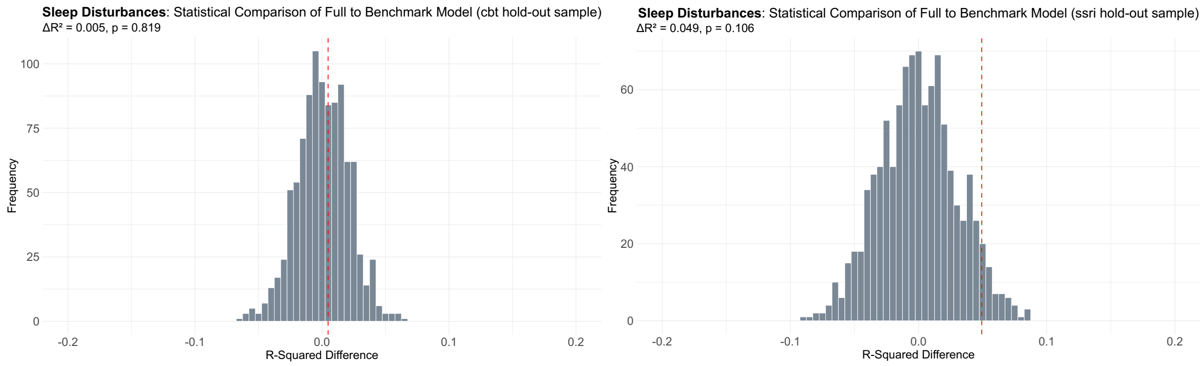


***Supplementary Figure 42. Statistical comparison of the full versus benchmark model for sleep disturbances scores.*** *This is shown for the psychotherapy (cbt) hold-out sample (left) and pharmacotherapy (ssri) hold-out sample (right). The distributions represent the frequency of R-squared difference values expected by chance based on 1,000 randomized permutations and the red dotted line on the plot indicates the true R-squared difference value between the full and benchmark model.*

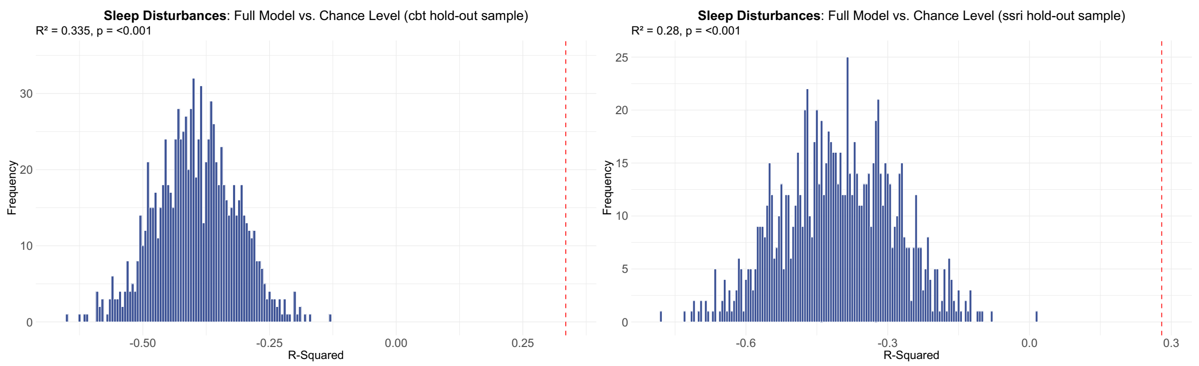


***Supplementary Figure 43. Statistical comparison of the full model versus chance level for sleep disturbances scores.*** *This is shown for the psychotherapy (cbt) hold-out sample (left)*

*and pharmacotherapy (ssri) hold-out sample (right). The distributions represent the frequency of R-squared values expected by chance based on 1,000 randomized permutations and the red dotted line on the plot indicates the true R-squared value of the full model.*

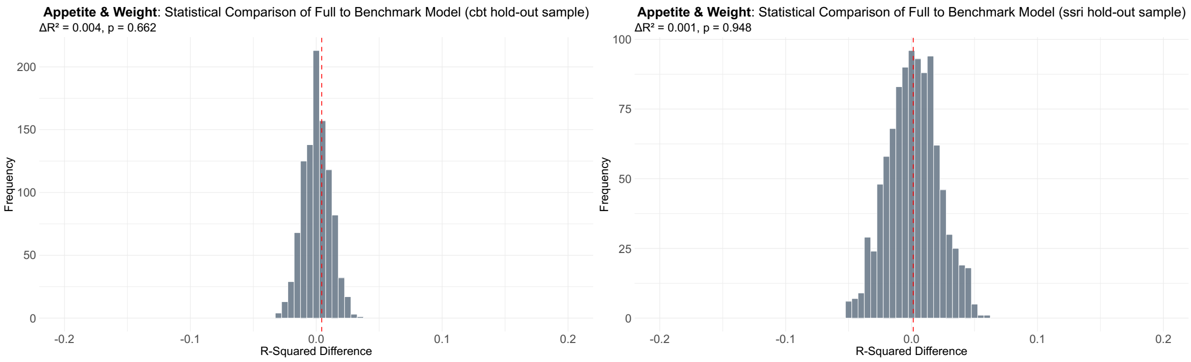


***Supplementary Figure 44. Statistical comparison of the full versus benchmark model for appetite and weight change scores.*** *This is shown for the psychotherapy (cbt) hold-out sample (left) and pharmacotherapy (ssri) hold-out sample (right). The distributions represent the frequency of R-squared difference values expected by chance based on 1,000 randomized permutations and the red dotted line on the plot indicates the true R-squared difference value between the full and benchmark model.*

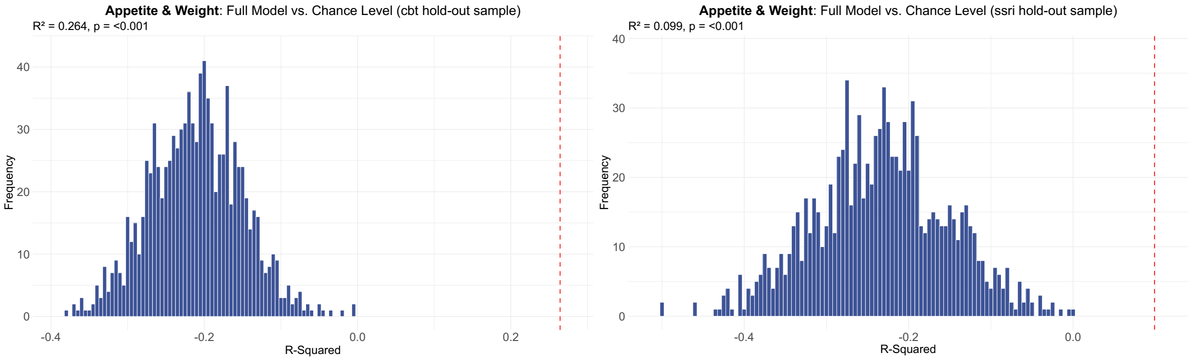


***Supplementary Figure 45. Statistical comparison of the full model versus chance level for appetite and weight change scores.*** *This is shown for the psychotherapy (cbt) hold-out sample (left) and pharmacotherapy (ssri) hold-out sample (right). The distributions represent the frequency of R-squared values expected by chance based on 1,000 randomized permutations and the red dotted line on the plot indicates the true R-squared value of the full model.*
