## Supplementary Material 2 for "Predictive Modelling of Depression Treatment Response using Individual Symptoms and Latent Factors"

### Feature Set: Detailed Predictor Variable Directory

| Predictor Domain: Demographic | | |
|---|---|---|
| **Feature** | **Item wording** | **Response Option** |
| age | "What is your age?" | 18-70 (numeric) |
| Sex | "What is your sex?" | Female (0), Male (1), Intersex (2), Male to Female (3), Female to Male (4) |
| MStatus | "What is your marital status?" | Single (0), Married (1), In a relationship (2), Separated (3), Divorced (4), Widowed (5) |
| Education | "What is the highest level of education that you've completed?" | No schooling completed (0), Some early primary (1), Completed primary school (2), some secondary education (3), completed secondary education (4), trade/technical/vocational training (5), some undergraduate education (6), completed undergraduate education (7), some postgraduate education (8), master's degree (9), doctorate degree (10) |
| Employment | "What best describes your current employment status?" | Unemployed not looking (0), unemployed looking (1), full-time employed (2), part-time employed (3), self-employed (4), retired (5) |
| SSStatus | MacArthur Scale of Subjective Social Status ("At the top of the ladder are the people who are the best off, those who have the most money, most education, and best jobs. At the bottom are the people who are the worst off, those who have the least money, least education, worst jobs, or no job. Please place an 'X' on the rung that best represents where you think you stand on the ladder.") | 0-10 (the higher the number, the higher the subjective status) |

| Predictor Domain: Physical Health | | |
|---|---|---|
| **Feature** | **Item wording** | **Response Option** |
| Exercise_days | "On average, how many days per week do you engage in moderate to strenuous exercise? (e.g. a brisk walk, a run, swimming, cycling, weight lifting, team sports etc.)". (Physical Activity Vital Sign; PAVS) | 0-7 |
| Exercise_min | "On these days, on average, how many minutes do you engage in exercise at this level?" | 0 min, 10 min, 20 min, 30 min, 40 min, 50 min, 60+ min |
| Diet_quality | "In general, how healthy is your overall diet?" | Very poor (0), Poor (1), Fair (2), Good (3), Very good (4), Excellent (5) |

| | | |
|---|---|---|
| Diet_fish | "How often do you usually eat fresh or canned fish? (NOT including fish and chips)" | rarely/never (0), once a month (1), twice a month (2), once a week (3), twice a week (4), every second day (5), once a day (6), more than once a day (7) |
| Diet_fishsupp | "Do you regularly take diet supplements that contain fish oils or omega 3 fatty acids?" | rarely/never (0), once a month (1), twice a month (2), once a week (3), twice a week (4), every second day (5), once a day (6), more than once a day (7) |
| Drug Use | Marijuana, Ecstasy/MDMA, Stimulant, Opiates, Street Opiates, Sedatives, Street Sedatives (48 variables, 1 variable on current street sedative use has no info) | |
| | Q2."How frequently did you take it in the past?" | If (1) above<br>Less than once a month (0), Every month (1), Every week (2), Daily or almost daily (3), More than once a day (4) |
| | Q5."How frequently do you take it?" | Less than once a month (0), Every month (1), Every week (2), Daily or almost daily (3), More than once a day (4) |
| Physical Health Comorbidities (CIRS) | Cumulative Illness Rating Scale (CIRS)<br>"These questions ask you about symptoms and impairment relating to different systems in your body." | None (0), Mild (1), Moderate (2), Severe (3), Extremely severe (4) |
| | 1. Cardiac (heart only) | |
| | 2. Hypertension (rating is based on severity; affected systems are rated separately) | |
| | 3. Vascular (blood, blood vessels and cells, marrow, spleen, lymphatics) | |
| | 4. Respiratory (lungs, bronchi, trachea below the larynx) | |
| | 5. ENT (eye, ear, nose, throat, larynx) | |
| | 6. Upper GI (esophagus, stomach, duodenum. Biliar and parcreatic trees; do not include diabetes) | |
| | 7. Lower GI (intestines, colon, bowel, hernias) | |
| | 8. Hepatic (liver only) | |
| | 9. Renal (kidneys only) | |
| | 10. Other GU (ureters, bladder, urethra, prostate, genitals) | |
| | 11. Musculo-Skeletal-Integumentary (muscles, bone, skin) | |
| | 12. Neurological (brain, spinal cord, nerves; do not include dementia) | |
| | 13. Endocrine-Metabolic (includes diabetes, thyroid dysfunction, diffuse infections, infections, toxicity) | |
| | CIRS_total | A total score exists but summing all 13 items. |

| | | |
|---|---|---|
| Pain | Patient Health Questionnaire (PHQ-15)<br>“During the past 7 days, how much have you been bothered by any of the following?” | Not bothered at all (0), Bothered a little (1), Bothered a lot (2). |
| | 1. Stomach pain | |
| | 2. Back pain | |
| | 3. Pain in your arms, legs, or joints (knees, hips, etc.) | |
| | 4. Headaches | |
| | 5. Chest pain | |
| | PAIN_total | A total score exists but summing all 5 items. |
| Smoking | Cigarettes, Vaping | |
| | Q2.“Please select the option which best applies to you.” | If (1) above<br>Less than once per day (0), 1-5 times (1), 5-10 times (2), 10-20 times (3), 20-30 times (4), more than 30 times (5) |
| | Q5. “How frequently do you smoke? (include instances where you have only taken one or two puffs) | Less than once per day (0), 1-5 times (1), 5-10 times (2), 10-20 times (3), 20-30 times (4), more than 30 times (5) |
| currentweight | “What is your weight at present (in lbs) (please give your best estimate)” | Free numeric field |
| FeetDropdownId +<br>InchesDropdownId | “What is your height (in feet and inches)?” | 1-8 feet<br>0-11 inches |

| Predictor Domain: Psychosocial | | |
|---|---|---|
| **Feature** | **Item wording** | **Response Option** |
| Stressful Life Events (SRRS) | Social Readjustment Rating Scale (SRRS)<br>“Indicate whether true or false these stressful life events over the last 12 months” | False (0), True (1) |
| | 1.partnerDeath | |
| | 2.Divorce | |
| | 3.Separation | |
| | 4.Jail | |
| | 5.familyDeath | |
| | 6.majorInjury | |

| | |
|---|---|
| | 7.Marriage |
| | 8.marReconciliation |
| | 9.Fired |
| | 10.Retirement |
| | 11.familyHealth |
| | 12.Pregnancy |
| | 13.sexualIssues |
| | 14.newFamMember |
| | 15.businessChange |
| | 16.financialChange |
| | 17.friendDeath |
| | 18.workChange |
| | 19.arguingPartner |
| | 20.newMortgage |
| | 21.mortgageForeclosure |
| | 22.workResponsibilities |
| | 23.childDeparture |
| | 24.inLaws |
| | 25.personalAchievement |
| | 26.partnerWork |
| | 27.schoolingChange |
| | 28.homeChange |
| | 29.habitChange |
| | 30.bossTroubles |
| | 31.workDuration |
| | 32.residenceChange |
| | 33.newSchool |
| | 34.recreationChange |
| | 35.churchChange |
| | 36.socialisingChange |
| | 37.Loan |
| | 38.sleepChange |
| | 39.familyGatherings |
| | 40.eatingChange |

| | | |
|---|---|---|
| | 41.Vacation | |
| | 42.majorHolidays | |
| | 43.lawViolations | |
| | SRRS_total | A total score is yielded by multiplying the ‘True’ answers by a specific weight. Some life events are thus classes as more stressful than others. |
| Childhood Trauma (CTQ) | Childhood Trauma Questionnaire (CTQ)<br>“Please select an answer that best describes how much each statement is true for you. While I was growing up, during the first 18 years of life:” | Never true (0), Rarely true (1), Sometimes true (2), Often true (3), Very often true (4). |
| | 1.I didn't have enough to eat | |
| | 2.I knew there was someone to take care of me and protect me. | |
| | 3.People in my family called me “stupid,” “lazy,” or “ugly.” | |
| | 4.My parents were too drunk or high to take care of the family. | |
| | 5.There was someone in my family who helped me feel that I was important or special. | |
| | 6. I had to wear dirty clothes. | |
| | 7. I felt loved. | |
| | 8. I thought that my parents wished I had never been born. | |
| | 9. Got hit so hard that I had to see a doctor or go to the hospital. | |
| | 10. There was nothing I wanted to change about my family. | |
| | 11. Family hit me so hard that it left me with bruises or marks. | |
| | 12. I was punished with a belt/board/cord other hard object. | |
| | 13. People in my family looked out for each other. | |
| | 14. People in my family said hurtful or insulting things to me. | |
| | 15. I believe that I was physically abused. | |
| | 16. I had the perfect childhood. | |
| | 17. I got hit or beaten so badly it was noticed by a teacher/neighbor/doctor. | |
| | 18. I felt that someone in my family hated me. | |
| | 19. People in my family felt close to each other. | |
| | 20. Someone tried to touch me in a sexual way or tried to make me touch them. | |
| | 21. Someone threatened to hurt me or tell lies about me unless I did something sexual. | |
| | 22. I had the best family in the world. | |

| | | |
|---|---|---|
| | 23. Someone tried to make me do or watch sexual things. | |
| | 24. Someone molested me. | |
| | 25. I believe that I was emotionally abused. | |
| | 26. There was someone to take me to the doctor if I needed it. | |
| | 27. I believe that I was sexually abused. | |
| | 28. My family was a source of strength and support. | |
| | CTQ_total | A total score and 5 subscale scores are yielded by summing respective item responses. |
| | CTQ_emabuse | |
| | CTQ_physabuse | |
| | CTQ_sexalbuse | |
| | CTQ_emneglect | |
| | CTQ_physneglect | |
| Perceived Social Support (SoSu) | Multidimensional Scale of Perceived Social Support (MSPSS)<br>"We are interested in how you feel about the following statements. Read each statement carefully. Indicate how you feel about each statement." | Very strongly disagree (0), Strongly disagree (1), Mildly disagree (2), Neutral (3), Mildly agree (4), Strongly Agree (5), Very strongly Agree (6) |
| | 1. There is a special person who is around when I am in need. | |
| | 2. There is a special person with whom I can share my joys and sorrows. | |
| | 3. My family really tries to help me. | |
| | 4. I get the emotional help and support I need from my family. | |
| | 5. I have a special person who is a real source of comfort to me. | |
| | 6. My friends really try to help me. | |
| | 7. I can count on my friends when things go wrong. | |
| | 8. I can talk about my problems with my family. | |
| | 9. I have friends with whom I can share my joys and sorrows. | |
| | 10. There is a special person in my life who cares about my feelings. | |
| | 11. My family is willing to help me make decisions. | |
| | 12. I can talk about my problems with my friends. | |
| | SoSu_total | A total score is yielded by summing respective item responses. |
| | Perceived Stress Scale (PSS) | Never (0), Almost Never (1), Sometimes (2), Fairly often (3), Very often (4). |

| | | |
|---|---|---|
| Perceived Stress (PSS) | "Indicate how often you felt/thought certain way during last month:" | |
| | 1.upset because of something that happened unexpectedly? | |
| | 2.felt that you were unable to control the important things in your life? | |
| | 3.nervous and "stressed"? | |
| | 4.confident about your ability to handle your personal problems? | |
| | 5.felt that things were going your way? | |
| | 6.found that you could not cope with all the things that you had to do? | |
| | 7.able to control irritations in your life? | |
| | 8.felt that you were on top of things? | |
| | 9.angered because of things that were outside of your control? | |
| | 10.felt difficulties were piling up so high that you could not overcome them? | |
| | PSS_total | A total score is yielded by summing respective item responses. |

| Predictor Domain: Mental Health | | |
|---|---|---|
| **Feature** | **Item wording** | **Response Option** |
| TFLTE_episodes | "How many times in your life have you experienced an episode of poor mental health?" | 0-10 times (10 being 10+ times) |
| TFLTE_ageonset | "What age were you when you experienced your first episode of poor mental health?" | 0-70 |
| TFLTE_currentonset | "When did this current episode of poor mental health start?" | DD/MM/YY |
| Psychiatric Diagnoses (Self) | "I currently have a diagnosis of… (Please select all that apply)" | Yes (1) and No (0) for 13 diagnoses. |
| | 1.Diagnosis_Dep | |
| | 2.Diagnosis_OCD | |
| | 3.Diagnosis_GAD | |
| | 4.Diagnosis_PanD | |
| | 5.Diagnosis_PTSD | |
| | 6.Diagnosis_BPD | |
| | 7.Diagnosis_Schiz | |
| | 8.Diagnosis_PersD | |

| | | |
|---|---|---|
| | 9.Diagnosis_SubD | |
| | 10.Diagnosis_AnoD | |
| | 11.Diagnosis_BulD | |
| | 12.Diagnosis_BinD | |
| | 13.Diagnosis_TicD | |
| | Diagnoses_total | A total score is yielded by summing the number of ‘YES for all diagnoses. |
| | Diagnoses_other | This is a composite total score variable that counts up the number of diagnoses one has that is not depression or anxiety. |
| Psychiatric Diagnosis (Family) | “Do you have any close relatives who ever received a diagnosis of a mental health disorder? (Please only count your biological parents, biological siblings or biological children)” | 0-10+ |
| Miscellaneous Psychiatric Symptoms (LP_items) | Top predictors in Chekroud and colleagues’ 2016 study “During the past week…” | Yes (1,) No (0) |
| | 1. Have you experienced depressed mood most of the day, nearly every day? | |
| | 2.Have you been bothered by aches and pains in many different parts of your body? | |
| | 3. Did reminders of a traumatic event make you shake, break out into a sweat, or have a racing heart? | |
| | 4.Did you try to avoid activities, places, or people that reminded you of a traumatic event? | |
| | 5. Did you have attacks of anxiety that caused you to avoid certain situations or to change your behaviour or normal routine? | |
| | 6. Did standing in long lines make you feel fearful, anxious, or nervous? | |
| | 7.Did driving or riding in a car make you feel fearful, anxious, or nervous? | |
| | 8. Have you ever witnessed a traumatic event such as rape, assault, someone dying in an accident, or any other extremely upsetting incident? | |
| | LP_total | A total score is yielded by summing respective item responses. This is not included. |
| Apathy (AES) | Apathy Evaluation Scale (AES) | A 4-point Likert scale for 18 items: Not at all (0), Slightly (1), Somewhat (2), A lot (3). |

| | | |
|---|---|---|
| | "For each statement, select the answer that best describes your thoughts, feelings, and activity in the past 4 weeks." | |
| | 1. I am interested in things | |
| | 2. I get things done during the day | |
| | 3. Getting things started on my own is important to me | |
| | 4. I am interested in having new experiences | |
| | 5. I am interested in learning new things | |
| | 6. I put little effort into anything | |
| | 7. I approach life with intensity | |
| | 8. Seeing a job through to the end is important to me | |
| | 9. I spend time doing things that interest me | |
| | 10. Someone has to tell me what to do each day | |
| | 11. I am less concerned about my problems than I should be | |
| | 12. I have friends | |
| | 13. Getting together with friends is important to me | |
| | 14. When something good happens, I get excited | |
| | 15. I have an accurate understanding of my problems | |
| | 16. Getting things done during the day is important to me | |
| | 17. I have initiative | |
| | 18. I have motivation | |
| | AES_total | A total score is yielded by summing respective item responses. This is not included. |
| Alcoholism (AUDIT) | Alcohol Use Disorder Identification Test (AUDIT)<br>"We will now ask some questions about your use of alcohol. Please select the option that best describes your answer to each question." | A 5-point Likert Scale for 8 items (0-4) and a 3-point Likert Scale for 2 items. |
| | 1. How often do you have a drink containing alcohol? | |
| | 2. How many drinks containing alcohol do you have on a typical day when you are drinking? | |
| | 3. How often do you have six or more drinks on one occasion? | |
| | 4. How often during the last year have you found that you were not able to stop drinking once you had started? | |
| | 5. How often during the last year have you failed to do what was normally expected of you because of drinking? | |

| | | |
|---|---|---|
| | 6. How often during the last year have you needed a first drink in the morning to get yourself going after a heavy drinking session? | |
| | 7. How often during the last year have you had a feeling of guilt or remorse after drinking? | |
| | 8. How often during the last year have you been unable to remember what happened the night before because of your drinking? | |
| | 9. Have you or someone else been injured because of your drinking? | |
| | 10. Has a relative, friend, doctor, or other health care worker been concerned about your drinking or suggested you cut down? | |
| | AUDIT_total | A total score is yielded by summing respective item responses. |
| Impulsivity (BIS) | Barratt Impulsivity Scale (BIS)<br>“People differ in the ways they act and think in different situations. This is a test to measure some of the ways in which you act and think. Read each statement and select the answer that describes you best. Do not spend too much time on any statement. Answer quickly and honestly.” | A 4-point Likert scale for 30 items: Rarely/Never (1), Occasionally (2), Often 3), Almost always/Always (4). |
| | 1. I plan tasks carefully. | |
| | 2. I do things without thinking. | |
| | 3. I make-up my mind quickly. | |
| | 4. I am happy-go-lucky. | |
| | 5. I don’t “pay attention.” | |
| | 6. I have “racing” thoughts. | |
| | 7. I plan trips well ahead of time. * | |
| | 8. I am self controlled. | |
| | 9.I concentrate easily. | |
| | 10.I save regularly. | |
| | 11. I “squirm” at plays or lectures. | |
| | 12. I am a careful thinker. | |
| | 13. I plan for job security. | |
| | 14. I say things without thinking. | |
| | 15. I like to think about complex problems. | |
| | 16. I change jobs. | |
| | 17. I act “on impulse.” | |

| | | |
|---|---|---|
| | 18. I get easily bored when solving thought problems | |
| | 19. I act on the spur of the moment. | |
| | 20. I am a steady thinker. | |
| | 21. I change residences. | |
| | 22. I buy things on impulse. | |
| | 23. I can only think about one thing at a time. | |
| | 24. I change hobbies. | |
| | 25. I spend or charge more than I earn. | |
| | 26. I often have extraneous thoughts when thinking. | |
| | 27. I am more interested in the present than the future. | |
| | 28. I am restless at the theater or lectures. | |
| | 29. I like puzzles. | |
| | 30. I am future oriented. | |
| | BIS_total | A total score is yielded by summing respective item responses. |
| Eating disorder (EAT) | Eating Attitudes Test (EAT-26)<br>“Please fill out the below form as accurately, honestly and completely as possible. There are no right or wrong answers. All of your responses are confidential. Please check a response for each of the following statements:” | A 6-point Likert scale for 26 items: Never (1), Rarely (2), Sometimes (3), Often (4), Usually (5), Always (6). |
| | 1.I am terrified about being overweight. | |
| | 2. I avoid eating when I am hungry. | |
| | 3. I find myself preoccupied with food. | |
| | 4.I have gone on eating binges where I feel that I may not be able to stop. | |
| | 5.I cut my food into small pieces. | |
| | 6. I am aware of the calorie content of foods I eat. | |
| | 7.I particularly avoid foods with high carbohydrate content. | |
| | 8.I feel that others would prefer if I ate more. | |
| | 9. I vomit after I have eaten. | |
| | 10. I feel extremely guilty after eating. | |
| | 11.I am preoccupied with a desire to be thinner. | |
| | 12. I think about burning up calories when I exercise. | |
| | 13. Other people think that I am too thin. | |
| | 14. I am preoccupied with the thought of having fat on my body | |

| | | |
|---|---|---|
| | 15. I take longer than others to eat meals. | |
| | 16. I avoid foods with sugar in them. | |
| | 17. I eat diet foods. | |
| | 18. I feel that food controls my life. | |
| | 19. I display self-control around food. | |
| | 20. I feel that others pressure me to eat. | |
| | 21. I give too much time and thought to food. | |
| | 22. I feel uncomfortable after eating sweets. | |
| | 23. I engage in dieting behaviour. | |
| | 24. I like my stomach to be empty. | |
| | 25. I have the impulse to vomit after meals. | |
| | 26. I enjoy trying new rich foods. | |
| | 27. In the past 6 months have you have you gone on eating binges where you feel that you may not be able to stop? | (5=once a day, 4=2-6 times a week, 3=once a week, 2=2-3 times a month, 1=once a month or less, 0=never) |
| | 28. In the past 6 months, ever made yourself sick (vomited) to control your weight or shape? | (5=once a day, 4=2-6 times a week, 3=once a week, 2=2-3 times a month, 1=once a month or less, 0=never) |
| | 29. In the past 6 months, ever used laxatives, diet pills, or diuretics (Water pills) to control your weight or shape? | (5=once a day, 4=2-6 times a week, 3=once a week, 2=2-3 times a month, 1=once a month or less, 0=never) |
| | 30. In the past 6 months, exercised more than 60 minutes a day to lose or to control your weight? | (5=once a day, 4=2-6 times a week, 3=once a week, 2=2-3 times a month, 1=once a month or less, 0=never) |
| | 31. Lost 20 pounds or more in the past 6 months | (0=no, 1=yes) |
| | EAT_total | A total score is yielded by summing respective item responses (1-26). |
| Social anxiety (LSAS) | Liebowitz Social Anxiety Scale (LSAS)<br>“Read each situation carefully and answer how anxious or fearful you feel in the situation. If you come across a situation that you ordinarily do not experience, we ask that you imagine "what if you were faced with that situation," and then rate the degree to which you would fear this hypothetical situation. Please base your ratings on the way that the situations have affected you in the last week. | A 4-point Likert scale for 24 items: None (0), Mild (1), Moderate (2), Severe (3). |
| | 1. Telephoning in public | |
| | 2. Participating in small groups | |
| | 3. Eating in public places. | |

| | | |
|---|---|---|
| | 4. Drinking with others in public places. | |
| | 5.Talking to people in authority. | |
| | 6. Acting, performing or giving a talk in front of an audience. | |
| | 7. Going to a party. | |
| | 8. Working while being observed. | |
| | 9. Writing while being observed. | |
| | 10. Calling someone you don’t know very well. | |
| | 11. Talking with people you don’t know very well. | |
| | 12. Meeting strangers. | |
| | 13. Urinating in a public bathroom. | |
| | 14. Entering a room when others are already seated. | |
| | 15. Being the center of attention. | |
| | 16. Speaking up at a meeting. | |
| | 17. Taking a test. | |
| | 18. Expressing a disagreement or disapproval to people you don't know very well. | |
| | 19. Looking at people you don’t know very well in the eyes. | |
| | 20. Giving a report to a group. | |
| | 21. Trying to pick up someone. | |
| | 22. Returning goods to a store. | |
| | 23. Giving a party. | |
| | 24. Resisting a high pressure salesperson. | |
| | LSAS_total | A total score is yielded by summing respective item responses (1-24). |
| Schizotypy (SSMS) | Short Scales for Measuring Schizotypy (SSMS)<br>“This questionnaire contains questions that may relate to your thoughts, feelings, experiences and preferences. There are no right or wrong answers or trick questions so please be as honest as possible. Do not spend too much time deliberating over each question, and choose answers that best reflect your own experiences.” | No (0) and Yes (1) |
| | 1.When in the dark do you often see shapes and forms even though there is nothing there? | |
| | 2.Are your thoughts sometimes so strong that you can almost hear them? | |

| | | |
|---|---|---|
| | 3.Have you ever thought that you had special, almost magical powers? | |
| | 4.Have you sometimes sensed an evil presence around you, even though you could not see it? | |
| | 5.Do you think that you could learn to read other's minds if you wanted to? | |
| | 6.When you look in the mirror does your face sometimes seem quite different from usual? | |
| | 7.Do ideas and insights sometimes come to you so fast that you cannot express them all? | |
| | 8.Can some people make you aware of them just by thinking about you? | |
| | 9. Does a passing thought ever seem so real it frightens you? | |
| | 10. Do you feel that your accidents are caused by mysterious forces? | Excluded rarely endorsed <10% |
| | 11. Do you ever have a sense of vague danger or sudden dread for reasons that you do not understand? | |
| | 12.Does your sense of smell sometimes become unusually strong? | |
| | 13. Are you easily confused if too much happens at the same time? | |
| | 14. Do you frequently have difficulty in starting to do things? | |
| | 15. Are you a person whose mood goes up and down easily? | |
| | 16. Do you dread going into a room by yourself where other people have already gathered and are talking? | |
| | 17. Do you find it difficult to keep interested in the same thing for a long time? | |
| | 18. Do you often have difficulties in controlling your thoughts? | |
| | 19. Are you easily distracted from work by daydreams? | |
| | 20. Do you ever feel that your speech is difficult to understand because the words are all mixed up and don't make sense? | |
| | 21. Are you easily distracted when you read or talk to someone? | |
| | 22. Is it hard for you to make decisions? | |
| | 23.When in a crowded room, do you often have difficulty in following a conversation? | |
| | 24. Are there very few things that you have ever enjoyed doing? | |
| | 25. Are you much too independent to get involved with other people? | |
| | 26. Do you love having your back massaged?* | |
| | 27. Do you find the bright lights of a city exciting to look at? * | |

| | | |
|---|---|---|
| | 28. Do you feel very close to your friends? * | |
| | 29. Has dancing or the idea of it always seemed dull to you? | |
| | 30. Do you like mixing with people? * | |
| | 31. Is trying new foods something you have always enjoyed? | |
| | 32. Have you often felt uncomfortable when your friends touch you? | |
| | 33. Do you prefer watching television to going out with people? | |
| | 34. Do you consider yourself to be pretty much an average sort of person? * | |
| | 35. Would you like other people to be afraid of you? | Excluded rarely endorsed <10% |
| | 36. Do you often feel the impulse to spend money which you know you can't afford? | |
| | 37. Are you usually in an average kind of mood, not too high and not too low? * | |
| | 38. Do you at times have an urge to do something harmful or shocking? | |
| | 39. Do you stop to think things over before doing anything? * | |
| | 40. Do you often overindulge in alcohol or food? | |
| | 41. Do you ever have the urge to break or smash things? | |
| | 42. Have you ever felt the urge to injure yourself? | |
| | 43. Do you often feel like doing the opposite of what other people suggest even though you know they are right? | |
| | SCZ_total | A total score is yielded by summing respective item responses (1-43). |
| Obsessive-Compulsive Disorder (OCI) | Revised Obsessive-Compulsive Inventory (OCI-R) "The following statements refer to experiences that many people have in their everyday lives. For each question, select an answer that best describes how much that experience has distressed or bothered you during the past month." | A 5-point Likert scale for 18 items: Not at all (0), A little (1), Moderately (2), A lot (3), Extremely (4). |
| | 1.I have saved up so many things that they get in the way. | |
| | 2.I check things more often than necessary. | |
| | 3.I get upset if objects are not arranged properly. | |
| | 4.I feel compelled to count while I am doing things. | |
| | 5.I find it difficult to touch an object when I know it has been touched by strangers or certain people. | |
| | 6.I find it difficult to control my own thoughts. | |

| | | |
|---|---|---|
| | 7.I collect things I don’t need. | |
| | 8.I repeatedly check doors, windows, drawers, etc. | |
| | 9.I get upset if others change the way I have arranged things. | |
| | 10.I feel I have to repeat certain numbers. | |
| | 11.I sometimes have to wash or clean myself simply because I feel contaminated. | |
| | 12.I am upset by unpleasant thoughts that come into my mind against my will. | |
| | 13.I avoid throwing things away because I am afraid I might need them later. | |
| | 14.I repeatedly check gas and water taps and light switches after turning them off. | |
| | 15.I need things to be arranged in a particular way. | |
| | 16.I feel that there are good and bad numbers. | |
| | 17.I wash my hands more often and longer than necessary. | |
| | 18.I frequently get nasty thoughts and have difficulty in getting rid of them. | |
| | OCI_total | A total score is yielded by summing respective item responses. |
| State Anxiety (STAI) | State-Trait Anxiety Inventory (STAI)<br>“Please read the following statements and then select the option that best describes how often you felt or behaved this way during the past several days.” | A 4-point Likert scale for 20 items: Not at all (1), Somewhat (2), Moderately so (3), Very much so (4). |
| | 1.I feel pleasant. | |
| | 2.I feel nervous and restless. | |
| | 3.I feel satisfied with myself. | |
| | 4.I wish I could be as happy as others seem to be. | |
| | 5.I feel like a failure. | |
| | 6.I feel rested. | |
| | 7.I am "calm, cool, and collected". | |
| | 8.I feel that difficulties are piling up so that I cannot overcome them. | |
| | 9.I worry too much over something that really doesn't matter. | |
| | 10.I am happy. | |
| | 11.I have disturbing thoughts. | |
| | 12.I lack self-confidence. | |

| | | |
|---|---|---|
| | 13.I feel secure. | |
| | 14.I make decisions easily. | |
| | 15.I feel inadequate. | |
| | 16.I am content. | |
| | 17.Some unimportant thought runs through my mind and bothers me. | |
| | 18.I take disappointments so keenly that I can't put them out of my mind. | |
| | 19.I am a steady person. | |
| | 20.I get in a state of tension or turmoil as I think over my recent concerns and interests. | |
| | STAI_total | A total score is yielded by summing respective item responses. This is not included. |
| Depression (QIDS-SR) | Quick Inventory of Depressive Symptomatology – Self-Report (QIDS-SR)<br>“Please select the response to each item that best describes you in relation to the past seven days.” | A 4-point Likert scale for 16 items (0 to 3). |
| | 1.falling asleep | |
| | 2.sleep during the night | |
| | 3.waking up too early | |
| | 4. sleeping too much | |
| | 5. feeling sad | |
| | 6. decreased appetite | |
| | 7. increased appetite | |
| | 8. decreased weight | |
| | 9. increased weight | |
| | 10.concentration / decision-making | |
| | 11.view of myself | |
| | 12. thoughts of death / suicide | |
| | 13. general interest | |
| | 14. energy level | |
| | 15. feeling slowed down | |
| | 16. feeling restless | |
| | QIDS_total | A total score is yielded by summing respective item responses (1-4 together, 6-9 together, 15-16 together). This is not included. |

| | | |
|---|---|---|
| Functional Impairment (WSAS) | Work and Social Adjustment Scale (WSAS) | A 8-point Likert scale for 5 items: Not at all (0), Slightly (2), Definitely (4), Markedly (6), Very severely (8). |
| | 1. my ability to work is impaired | |
| | 2. my home management (cleaning, shopping, cooking, looking after children, paying bills) is impaired | |
| | 3. my social leisure activities (with other people e.g. parties, bars, clubs, visits, dating) are impaired | |
| | 4. my private leisure activities (done alone e.g. reading, gardening, sewing, walking alone) are impaired | |
| | 5. my ability to form and maintain close relationships with others, including those I live with, is impaired | |
| | WSAS_total | A total score is yielded by summing respective item responses. |
| Depression (SDS) | Self-Rating Depression Scale (SDS).<br>"Please read the following statements and then select the option that best describes how often you felt or behaved this way during the past several days." | A 4 Likert scale for 20 items: A little of the time (1), Some of the time (2), Good part of the time (3), Most of the time (4). |
| | 1.I feel down-hearted and blue. | |
| | 2.Morning is when I feel the best. | |
| | 3.I have crying spells or feel like it. | |
| | 4.I have trouble sleeping at night. | |
| | 5.I eat as much as I used to. | |
| | 6.I still enjoy sex. | |
| | 7.I notice that I am losing weight. | |
| | 8.I have trouble with constipation. | |
| | 9.My heart beats faster than normal. | |
| | 10.I get tired for no reason. | |
| | 11.My mind is as clear as it used to be. | |
| | 12.I find it easy to do the things I used to do. | |
| | 13.I am restless and can't keep still. | |
| | 14.I feel hopeful about the future. | |
| | 15.I am more irritable than usual. | |
| | 16.I find it easy to make decisions. | |
| | 17.I feel that I am useful and needed. | |
| | 18.My life is pretty full. | |
| | 19.I feel that others would be better off if I were dead. | |

| | | |
|---|---|---|
| | 20.I still enjoy the things I used to do. | |
| | SDS_total | A total score is yielded by summing respective item responses. |

| Category – Treatment | | |
|---|---|---|
| **Feature** | **Item wording** | **Response Option** |
| TFLTE_ADpast / TFLTE_PTpast | Items asked separately for past antidepressant medication treatment and psychotherapy treatment | |
| | Q.1 “Have you ever in the past completed a course of treatment?” (TFLTE_ADpast / TFLTE_PTpast) | No (0), Yes (1) |
| TFLTE_ADpast_help ed / TFLTE_PTpast_help ed | If Yes (1) on Q.1<br>Q.3 “How many times would you say this kind of treatment has resulted in a significant improvement in your symptoms? “ (TFLTE_ADpast_helped / TFLTE_PTpast_helped) | 0-10+ times |
| TMT_expectations | “What point on this 10-point scale best describes your expectations about what is likely to happen as a result of your current antidepressant treatment?” | “I don’t expect to feel any better" (0) to "I expect to feel completely better" (9) |

| Predictor Domain - Cognitive | | |
|---|---|---|
| **Feature** | **Item wording** | **Response Option** |
| Perceptual Decision-Making Task | Dot Discrimination Task. A perceptual decision-making task which dissociates between decision-formation and two components of metacognitive evaluation. | |
| | mratio | Numeric |
| | Confidence | Numeric (0-6) |
| Two-Step Reinforcement-Learning Task | Two-step reinforcement-learning task. A reinforcement learning task where two sequential decisions are made on each trial with a goal of maximising reward that is probabilistically associated with stimuli in the second stage choice. | |
| | Reward | Numeric |
| | Mbi | Numeric |
| | Stay | Numeric |
| | Rt_inter | Numeric |
| | RT_trans | Numeric |
| Learning Under Volatility Task | An aversive learning task. To measure how well subjects learning rates adjust to environmental volatility | |
| | Stable_lr | Numeric |
| | Vol_lr | Numeric |
| | Stable_a | Numeric |
| | Vol_a | Numeric |
| | Stable_beta | Numeric |
| | Vol_beta | Numeric |

### Elastic Net Model Results: Predictor Coefficients

| predictor | Total Scores | Appetite & Weight Factor | Sleep Disturbances Factor | Negative Affect & Thought Factor | Motivation & Cognition Factor | Insomnia (sleep onset) | Insomnia (mid-nocturnal) | Insomnia (early morning) | Hypersomnia | Sad Mood | Appetite decreased | Appetite increased | Weight decreased | Weight increased | Concentration or Decision-Making Problems | Self-critical thoughts | Suicidal ideation | Loss of interest | Loss of energy | Psychomotor slowing | Psychomotor agitation |
|---|---|---|---|---|---|---|---|---|---|---|---|---|---|---|---|---|---|---|---|---|---|
| Total Scores | 0.09 | 0.00 | 0.00 | 0.05 | 0.04 | 0.00 | 0.00 | 0.00 | 0.00 | 0.01 | 0.00 | 0.00 | 0.00 | 0.00 | 0.00 | 0.00 | 0.00 | 0.08 | 0.00 | 0.00 | 0.00 |
| Baseline Appetite & Weight Factor | 0.00 | 0.35 | 0.00 | 0.00 | 0.00 | 0.00 | 0.00 | 0.00 | 0.00 | 0.00 | 0.00 | 0.00 | 0.00 | 0.00 | 0.00 | 0.00 | 0.00 | 0.00 | 0.00 | 0.00 | 0.00 |
| Baseline Sleep Disturbances Factor | 0.00 | 0.00 | 0.35 | 0.00 | 0.00 | 0.00 | 0.00 | 0.00 | 0.00 | 0.00 | 0.00 | 0.00 | 0.00 | 0.00 | 0.00 | 0.00 | 0.00 | 0.00 | 0.00 | 0.00 | 0.00 |
| Baseline Negative Affect & Thought Factor | 0.00 | 0.00 | 0.00 | 0.16 | 0.01 | 0.00 | 0.00 | 0.00 | 0.00 | 0.00 | 0.00 | 0.00 | 0.00 | 0.00 | 0.00 | 0.00 | 0.00 | 0.00 | 0.00 | 0.00 | 0.00 |
| Baseline Motivation & Cognition Factor | 0.00 | 0.00 | 0.00 | 0.03 | 0.14 | 0.00 | 0.00 | 0.00 | 0.00 | 0.00 | 0.00 | 0.00 | 0.00 | 0.00 | 0.00 | 0.00 | 0.00 | 0.00 | 0.00 | 0.00 | 0.00 |
| Baseline Insomnia (sleep onset) | 0.04 | 0.00 | 0.02 | 0.01 | 0.03 | 0.60 | 0.00 | 0.00 | 0.00 | 0.00 | 0.03 | 0.00 | 0.00 | 0.00 | 0.00 | 0.00 | 0.00 | 0.00 | 0.02 | 0.00 | 0.00 |
| Baseline Insomnia (mid-nocturnal) | 0.06 | 0.00 | 0.08 | 0.04 | 0.04 | 0.00 | 0.46 | 0.13 | 0.00 | 0.07 | 0.01 | 0.00 | 0.00 | 0.00 | 0.00 | 0.04 | 0.00 | 0.00 | 0.01 | 0.00 | 0.00 |
| Baseline Insomnia (early morning) | 0.04 | 0.00 | 0.08 | 0.00 | 0.03 | 0.02 | 0.04 | 0.45 | 0.00 | 0.00 | 0.02 | 0.00 | 0.00 | 0.00 | 0.00 | 0.00 | 0.00 | 0.00 | 0.02 | 0.00 | 0.01 |
| Baseline Hypersomnia | 0.03 | 0.00 | -0.06 | 0.01 | 0.06 | 0.00 | 0.00 | 0.00 | 0.61 | 0.00 | 0.00 | 0.00 | 0.00 | 0.00 | 0.00 | 0.01 | 0.00 | 0.00 | 0.01 | 0.00 | 0.01 |
| Baseline Sad Mood | 0.00 | 0.00 | 0.00 | 0.00 | 0.00 | 0.00 | 0.00 | 0.00 | 0.00 | 0.28 | 0.00 | 0.00 | 0.00 | 0.00 | 0.00 | 0.00 | 0.00 | 0.00 | 0.00 | 0.00 | 0.00 |
| Baseline Appetite decreased | 0.04 | 0.00 | 0.04 | 0.06 | 0.03 | 0.05 | 0.02 | 0.02 | 0.00 | 0.03 | 0.35 | 0.00 | 0.07 | 0.00 | 0.00 | 0.00 | 0.01 | 0.01 | 0.00 | 0.00 | 0.03 |
| Baseline Appetite increased | 0.02 | 0.08 | 0.00 | 0.00 | 0.00 | 0.00 | 0.00 | 0.00 | 0.00 | 0.00 | 0.00 | 0.42 | 0.00 | 0.16 | 0.00 | 0.00 | 0.00 | 0.00 | 0.00 | 0.00 | 0.00 |
| Baseline Weight decreased | 0.01 | 0.00 | 0.00 | 0.00 | 0.00 | 0.00 | 0.00 | 0.00 | 0.00 | 0.00 | 0.04 | 0.00 | 0.19 | 0.00 | 0.00 | 0.00 | 0.00 | 0.01 | 0.00 | 0.00 | 0.00 |
| Baseline Weight increased | 0.00 | 0.00 | 0.00 | 0.00 | 0.00 | 0.00 | 0.00 | 0.00 | 0.00 | 0.00 | 0.00 | 0.04 | 0.00 | 0.25 | 0.00 | 0.00 | 0.00 | 0.00 | 0.00 | 0.00 | 0.00 |
| Baseline Concentration or Decision-Making Problems | 0.04 | 0.00 | 0.00 | 0.00 | 0.02 | 0.00 | 0.00 | 0.00 | 0.00 | 0.00 | 0.00 | 0.00 | 0.00 | 0.00 | 0.24 | 0.01 | 0.00 | 0.00 | 0.00 | 0.06 | 0.02 |
| Baseline Self-critical thoughts | 0.01 | 0.00 | 0.00 | 0.00 | 0.00 | 0.00 | 0.00 | 0.00 | 0.00 | 0.00 | -0.00 | 0.00 | 0.00 | 0.00 | 0.00 | 0.31 | 0.00 | 0.00 | 0.00 | 0.00 | 0.00 |
| Baseline Suicidal ideation | 0.05 | 0.00 | 0.00 | 0.09 | 0.00 | 0.01 | 0.00 | 0.00 | 0.02 | 0.01 | 0.00 | 0.00 | 0.00 | 0.00 | 0.00 | 0.02 | 0.59 | 0.00 | 0.00 | 0.00 | 0.00 |
| Baseline Loss of interest | 0.00 | 0.00 | 0.00 | 0.00 | 0.00 | 0.00 | 0.00 | 0.00 | 0.00 | 0.00 | 0.00 | 0.00 | 0.00 | 0.00 | 0.00 | 0.00 | 0.00 | 0.16 | 0.00 | 0.00 | 0.00 |
| Baseline Loss of energy | 0.02 | 0.00 | 0.00 | 0.00 | 0.03 | 0.00 | 0.00 | 0.00 | 0.00 | 0.00 | 0.04 | 0.00 | 0.00 | 0.00 | 0.00 | 0.00 | 0.00 | 0.00 | 0.24 | 0.00 | 0.00 |
| Baseline Psychomotor slowing | 0.04 | 0.01 | 0.00 | 0.00 | 0.03 | 0.00 | 0.00 | 0.00 | 0.00 | 0.00 | 0.00 | 0.00 | 0.00 | 0.00 | 0.05 | 0.01 | 0.00 | 0.00 | 0.00 | 0.26 | 0.00 |
| Baseline Psychomotor agitation | 0.04 | 0.00 | 0.00 | 0.01 | 0.00 | 0.03 | 0.00 | 0.00 | 0.00 | 0.00 | 0.00 | 0.00 | 0.00 | 0.00 | 0.04 | 0.00 | 0.00 | 0.00 | 0.00 | 0.00 | 0.37 |

| predictor | Total Scores | Appetite & Weight Factor | Sleep Disturbances Factor | Negative Affect & Thought Factor | Motivation & Cognition Factor | Insomnia (sleep onset) | Insomnia (mid-nocturnal) | Insomnia (early morning) | Hypersomnia | Sad Mood | Appetite decreased | Appetite increased | Weight decreased | Weight increased | Concentration or Decision-Making Problems | Self-critical thoughts | Suicidal ideation | Loss of interest | Loss of energy | Psychomotor slowing | Psychomotor agitation |
|---|---|---|---|---|---|---|---|---|---|---|---|---|---|---|---|---|---|---|---|---|---|
| MStatus | 0.00 | 0.04 | 0.00 | 0.00 | 0.00 | 0.00 | 0.00 | 0.00 | 0.00 | 0.00 | -0.04 | 0.00 | 0.00 | 0.00 | 0.00 | 0.00 | 0.00 | 0.00 | 0.00 | 0.00 | 0.00 |
| age | 0.00 | 0.00 | 0.03 | 0.00 | 0.00 | 0.00 | 0.02 | 0.03 | 0.00 | 0.00 | -0.01 | 0.00 | 0.00 | 0.00 | 0.00 | 0.00 | -0.00 | 0.00 | 0.00 | 0.03 | 0.00 |
| SoSu_total | -0.06 | -0.00 | -0.02 | -0.07 | -0.06 | 0.00 | 0.00 | -0.03 | 0.00 | -0.07 | 0.00 | -0.01 | 0.00 | -0.01 | -0.03 | -0.02 | -0.05 | -0.01 | -0.03 | -0.04 | -0.00 |
| SDS_total | 0.12 | 0.00 | 0.04 | 0.13 | 0.14 | 0.02 | 0.08 | 0.03 | 0.02 | 0.08 | 0.03 | 0.00 | 0.00 | 0.00 | 0.07 | 0.16 | 0.03 | 0.10 | 0.10 | 0.15 | 0.03 |
| TMT_expectations | -0.08 | 0.00 | -0.04 | -0.09 | -0.06 | -0.02 | 0.00 | -0.01 | 0.00 | -0.09 | -0.05 | 0.00 | -0.01 | 0.00 | -0.01 | -0.02 | -0.05 | -0.06 | 0.00 | 0.00 | -0.08 |
| LP_1.moodDSM | 0.05 | 0.00 | 0.00 | 0.06 | 0.02 | 0.00 | 0.00 | 0.00 | 0.00 | 0.10 | 0.00 | 0.00 | 0.00 | 0.00 | 0.00 | 0.00 | 0.00 | 0.03 | 0.00 | 0.00 | 0.01 |
| MH_ageonset_adolescence | 0.05 | 0.00 | 0.04 | 0.05 | 0.04 | 0.00 | 0.00 | 0.02 | 0.00 | 0.00 | 0.11 | 0.00 | 0.00 | 0.00 | 0.00 | 0.09 | 0.00 | 0.00 | 0.00 | 0.00 | 0.00 |
| WSAS_total | 0.06 | 0.00 | 0.00 | 0.04 | 0.07 | 0.01 | 0.00 | 0.00 | 0.00 | 0.04 | 0.00 | 0.00 | 0.00 | 0.00 | 0.09 | 0.03 | 0.00 | 0.05 | 0.06 | 0.00 | 0.00 |
| AES_total | 0.06 | 0.00 | 0.00 | 0.03 | 0.07 | 0.07 | 0.01 | 0.00 | 0.00 | 0.00 | 0.00 | 0.00 | 0.00 | 0.00 | 0.01 | 0.02 | 0.01 | 0.07 | 0.04 | 0.02 | 0.00 |
| LP_4.trauma2PDSQ | 0.01 | 0.00 | 0.00 | 0.02 | 0.00 | 0.00 | 0.00 | 0.00 | 0.00 | 0.04 | 0.00 | 0.00 | 0.00 | 0.00 | 0.00 | 0.00 | 0.00 | 0.01 | 0.00 | 0.00 | 0.00 |
| Diet_fish | -0.02 | 0.00 | 0.00 | -0.02 | -0.02 | 0.00 | 0.00 | 0.00 | 0.00 | -0.00 | 0.00 | 0.01 | 0.00 | 0.00 | 0.00 | -0.02 | -0.02 | 0.00 | -0.03 | 0.00 | -0.00 |
| StressLifeEvents_total | 0.03 | 0.00 | 0.03 | 0.02 | 0.03 | 0.00 | 0.01 | 0.01 | 0.00 | 0.00 | 0.00 | 0.00 | 0.04 | 0.00 | 0.00 | 0.00 | 0.00 | 0.00 | 0.02 | 0.00 | 0.00 |
| OCI_total | 0.02 | 0.00 | 0.00 | 0.02 | 0.01 | 0.00 | 0.00 | 0.00 | 0.00 | 0.01 | 0.00 | 0.00 | 0.00 | 0.00 | 0.00 | 0.00 | 0.00 | 0.02 | 0.00 | 0.02 | 0.03 |
| BIS_total | 0.03 | 0.00 | 0.01 | 0.01 | 0.01 | 0.00 | 0.00 | 0.00 | 0.00 | 0.00 | 0.00 | 0.00 | 0.02 | 0.00 | 0.03 | 0.00 | 0.00 | 0.00 | 0.00 | 0.00 | 0.04 |
| Diagnoses_other | 0.02 | 0.00 | 0.03 | 0.01 | 0.00 | 0.00 | 0.00 | 0.01 | 0.00 | 0.00 | 0.00 | 0.00 | 0.00 | 0.00 | 0.00 | 0.06 | 0.00 | 0.00 | 0.00 | 0.02 | 0.05 |
| stable_lr | -0.01 | 0.00 | 0.00 | -0.01 | 0.00 | 0.00 | 0.00 | 0.00 | 0.00 | -0.01 | 0.00 | 0.00 | 0.00 | 0.00 | 0.00 | 0.00 | 0.00 | -0.00 | 0.00 | 0.00 | 0.00 |
| vol_a | -0.01 | 0.00 | 0.00 | -0.01 | -0.01 | 0.00 | 0.00 | 0.00 | 0.00 | 0.00 | -0.03 | 0.00 | 0.00 | 0.00 | 0.00 | -0.03 | 0.00 | -0.01 | 0.00 | 0.00 | 0.00 |
| LSAS_total | 0.03 | 0.00 | 0.00 | 0.01 | 0.01 | 0.00 | 0.00 | 0.00 | 0.00 | 0.00 | 0.00 | 0.00 | 0.00 | 0.00 | 0.01 | 0.06 | 0.00 | 0.00 | 0.00 | 0.05 | 0.00 |
| CIRS_total | 0.02 | 0.00 | 0.00 | 0.01 | 0.02 | 0.00 | 0.00 | 0.00 | 0.00 | 0.02 | 0.00 | 0.00 | 0.00 | 0.00 | 0.00 | 0.00 | 0.00 | 0.00 | 0.06 | 0.02 | 0.01 |
| mbi | 0.01 | 0.00 | 0.00 | 0.00 | 0.00 | 0.00 | 0.00 | -0.01 | 0.00 | 0.01 | 0.00 | 0.00 | 0.00 | 0.00 | 0.01 | 0.00 | 0.00 | 0.00 | 0.00 | 0.00 | 0.00 |
| CTQ_total | 0.00 | 0.00 | 0.01 | 0.00 | 0.00 | 0.00 | 0.00 | 0.03 | 0.00 | 0.00 | 0.00 | 0.00 | 0.00 | 0.00 | 0.00 | 0.00 | 0.00 | 0.00 | 0.00 | 0.00 | 0.00 |
| Sex | 0.00 | 0.00 | 0.00 | 0.00 | 0.00 | 0.00 | 0.00 | 0.00 | 0.00 | 0.01 | 0.00 | 0.00 | 0.00 | 0.00 | 0.00 | 0.00 | 0.00 | 0.00 | 0.00 | 0.00 | 0.00 |

| predictor | Total Scores | Appetite & Weight Factor | Sleep Disturbances Factor | Negative Affect & Thought Factor | Motivation & Cognition Factor | Insomnia (sleep onset) | Insomnia (mid-nocturnal) | Insomnia (early morning) | Hypersomnia | Sad Mood | Appetite decreased | Appetite increased | Weight decreased | Weight increased | Concentration or Decision-Making Problems | Self-critical thoughts | Suicidal ideation | Loss of interest | Loss of energy | Psychomotor slowing | Psychomotor agitation |
|---|---|---|---|---|---|---|---|---|---|---|---|---|---|---|---|---|---|---|---|---|---|
| LP_6.FRLNE | 0.04 | 0.00 | 0.00 | 0.00 | 0.03 | 0.00 | 0.00 | 0.00 | 0.00 | 0.00 | 0.00 | 0.00 | 0.00 | 0.00 | 0.00 | 0.00 | 0.00 | 0.06 | 0.00 | 0.10 | 0.18 |
| Exercise_min | -0.02 | 0.00 | -0.00 | 0.00 | -0.02 | 0.00 | 0.00 | -0.00 | 0.00 | 0.00 | 0.00 | 0.00 | 0.00 | 0.00 | 0.00 | 0.00 | 0.00 | -0.01 | -0.06 | 0.00 | 0.00 |
| Diagnoses_total | 0.01 | 0.00 | 0.00 | 0.00 | 0.02 | 0.00 | 0.00 | 0.00 | 0.00 | 0.00 | 0.00 | 0.00 | 0.00 | 0.00 | 0.00 | 0.00 | 0.00 | 0.00 | 0.03 | 0.00 | 0.00 |
| Diet_fishsupp | 0.01 | 0.00 | 0.00 | 0.00 | 0.02 | 0.00 | 0.00 | 0.00 | 0.00 | 0.02 | 0.00 | 0.00 | 0.00 | 0.00 | 0.00 | 0.00 | 0.00 | 0.01 | 0.00 | 0.00 | 0.00 |
| Education | -0.01 | 0.00 | 0.00 | 0.00 | -0.01 | 0.00 | 0.00 | 0.00 | 0.00 | 0.00 | 0.00 | 0.00 | 0.00 | 0.00 | 0.00 | 0.00 | 0.00 | -0.00 | 0.00 | 0.00 | 0.00 |
| rt_inter | -0.01 | 0.00 | 0.00 | 0.00 | -0.00 | 0.00 | 0.00 | 0.00 | -0.01 | 0.00 | -0.01 | 0.00 | 0.00 | 0.00 | 0.00 | 0.00 | 0.00 | 0.00 | 0.00 | 0.00 | -0.03 |
| SCZ_total | 0.01 | 0.00 | 0.00 | 0.00 | 0.00 | 0.00 | 0.01 | 0.00 | 0.00 | 0.00 | 0.00 | 0.00 | 0.00 | 0.00 | 0.00 | 0.00 | 0.00 | 0.01 | 0.00 | 0.00 | 0.00 |
| LP_7.FRCAR | 0.00 | 0.00 | -0.05 | 0.00 | 0.00 | 0.00 | 0.00 | 0.00 | 0.00 | -0.00 | -0.04 | 0.00 | 0.00 | 0.00 | 0.00 | 0.00 | 0.00 | 0.00 | 0.00 | 0.00 | 0.00 |
| confidence | 0.00 | 0.00 | 0.03 | 0.00 | 0.00 | 0.00 | 0.00 | 0.01 | 0.00 | 0.00 | 0.00 | 0.00 | 0.01 | 0.00 | -0.02 | 0.00 | 0.00 | 0.00 | 0.00 | 0.00 | 0.00 |
| SSStatus | 0.00 | 0.00 | -0.02 | 0.00 | 0.00 | -0.01 | 0.00 | -0.01 | 0.00 | 0.00 | 0.00 | 0.00 | 0.00 | 0.00 | 0.00 | 0.00 | -0.00 | 0.00 | 0.00 | 0.00 | 0.00 |
| BMI | 0.02 | 0.00 | 0.01 | 0.00 | 0.00 | 0.00 | 0.00 | 0.00 | 0.00 | 0.00 | 0.00 | 0.00 | 0.00 | 0.00 | 0.00 | 0.00 | 0.00 | 0.00 | 0.00 | 0.00 | 0.00 |
| Exercise_days | -0.00 | 0.00 | -0.01 | 0.00 | 0.00 | 0.00 | 0.00 | -0.06 | 0.00 | 0.00 | 0.00 | 0.00 | 0.00 | 0.00 | 0.00 | 0.00 | 0.00 | 0.00 | 0.00 | 0.00 | 0.00 |
| PAIN_total | 0.00 | 0.00 | 0.01 | 0.00 | 0.00 | 0.00 | 0.00 | 0.00 | 0.00 | 0.00 | 0.00 | 0.00 | 0.00 | 0.00 | -0.01 | 0.00 | 0.00 | 0.00 | 0.00 | 0.00 | 0.00 |
| AUDIT_total | 0.00 | 0.00 | -0.00 | 0.00 | 0.00 | 0.00 | 0.00 | 0.00 | 0.00 | 0.00 | 0.00 | 0.00 | 0.03 | 0.00 | 0.00 | 0.00 | 0.00 | 0.00 | 0.00 | 0.00 | 0.00 |
| rt_trans | 0.00 | 0.00 | -0.00 | 0.00 | 0.00 | 0.00 | 0.00 | -0.00 | 0.00 | 0.00 | 0.00 | -0.03 | 0.00 | 0.00 | 0.00 | 0.00 | 0.00 | 0.00 | 0.00 | 0.00 | 0.00 |
| MH_episode_currentonset | 0.00 | 0.00 | 0.00 | 0.00 | 0.00 | -0.01 | 0.00 | 0.00 | 0.00 | 0.00 | 0.00 | 0.00 | 0.00 | 0.00 | 0.00 | 0.00 | 0.00 | 0.00 | 0.00 | 0.01 | 0.00 |
| MH_ageonset_adulthood | 0.00 | 0.00 | 0.00 | 0.00 | 0.00 | -0.00 | 0.00 | 0.00 | 0.00 | 0.00 | 0.00 | 0.00 | 0.00 | 0.00 | -0.01 | 0.00 | -0.00 | 0.00 | 0.00 | 0.00 | 0.00 |
| Smoking_present_binary | 0.00 | 0.00 | 0.00 | 0.00 | 0.00 | 0.00 | 0.00 | -0.01 | 0.00 | 0.00 | 0.00 | 0.00 | 0.00 | 0.00 | 0.00 | 0.00 | -0.00 | 0.00 | 0.00 | 0.00 | 0.00 |
| LP_5.ANAVD | -0.00 | 0.00 | 0.00 | 0.00 | 0.00 | 0.00 | 0.00 | 0.00 | 0.06 | 0.00 | 0.00 | 0.00 | 0.00 | 0.00 | 0.00 | 0.00 | 0.00 | 0.00 | 0.00 | 0.00 | 0.00 |
| Diet_quality | 0.00 | 0.00 | 0.00 | 0.00 | 0.00 | 0.00 | 0.00 | 0.00 | -0.02 | 0.00 | 0.00 | -0.03 | 0.00 | 0.00 | 0.00 | 0.00 | 0.00 | 0.00 | 0.00 | 0.00 | 0.00 |
| TMT_ADpast | 0.00 | 0.00 | 0.00 | 0.00 | 0.00 | 0.00 | 0.00 | 0.00 | 0.00 | 0.02 | -0.05 | 0.00 | 0.00 | 0.00 | 0.00 | 0.00 | 0.00 | 0.00 | 0.00 | 0.00 | 0.00 |
| EAT_total | 0.00 | 0.00 | 0.00 | 0.00 | 0.00 | 0.00 | 0.00 | 0.00 | 0.00 | 0.00 | 0.02 | 0.00 | 0.04 | 0.00 | 0.00 | 0.00 | 0.00 | 0.02 | 0.00 | 0.00 | 0.00 |

| predictor | Total Scores | Appetite & Weight Factor | Sleep Disturbances Factor | Negative Affect & Thought Factor | Motivation & Cognition Factor | Insomnia (sleep onset) | Insomnia (mid-nocturnal) | Insomnia (early morning) | Hypersomnia | Sad Mood | Appetite decreased | Appetite increased | Weight decreased | Weight increased | Concentration or Decision-Making Problems | Self-critical thoughts | Suicidal ideation | Loss of interest | Loss of energy | Psychomotor slowing | Psychomotor agitation |
|---|---|---|---|---|---|---|---|---|---|---|---|---|---|---|---|---|---|---|---|---|---|
| Drugs_past_binary | 0.00 | 0.00 | 0.00 | 0.00 | 0.00 | 0.00 | 0.00 | 0.00 | 0.00 | 0.00 | -0.00 | 0.00 | 0.00 | 0.00 | 0.00 | 0.00 | 0.00 | 0.00 | 0.00 | 0.00 | 0.00 |
| STAI_total | 0.02 | 0.00 | 0.00 | 0.00 | 0.00 | 0.00 | 0.00 | 0.00 | 0.00 | 0.00 | 0.00 | 0.01 | 0.00 | 0.00 | 0.00 | 0.03 | 0.00 | 0.00 | 0.00 | 0.00 | 0.00 |
| LP_2.painPDSQ | 0.00 | 0.00 | 0.00 | 0.00 | 0.00 | 0.00 | 0.00 | 0.00 | 0.00 | 0.00 | 0.00 | 0.00 | 0.00 | 0.00 | -0.10 | -0.05 | 0.00 | 0.00 | 0.02 | 0.00 | 0.00 |
| PSS_total | 0.00 | 0.00 | 0.00 | 0.00 | 0.00 | 0.00 | 0.00 | 0.00 | 0.00 | 0.00 | 0.00 | 0.00 | 0.00 | 0.00 | 0.03 | 0.00 | 0.00 | 0.00 | 0.00 | 0.00 | 0.00 |
| reward | 0.01 | 0.00 | 0.00 | 0.00 | 0.00 | 0.00 | 0.00 | 0.00 | 0.00 | 0.00 | 0.00 | 0.00 | 0.00 | 0.00 | 0.03 | 0.05 | 0.00 | 0.00 | 0.00 | 0.00 | 0.01 |
| TMT_PTpast_helped | -0.00 | 0.00 | 0.00 | 0.00 | 0.00 | 0.00 | 0.00 | 0.00 | 0.00 | 0.00 | 0.00 | 0.00 | 0.00 | 0.00 | -0.02 | -0.04 | 0.00 | 0.00 | 0.00 | 0.00 | 0.00 |
| TMT_ADpast_helped | 0.00 | 0.00 | 0.00 | 0.00 | 0.00 | 0.00 | 0.00 | 0.00 | 0.00 | 0.00 | 0.00 | 0.00 | 0.00 | 0.00 | -0.01 | 0.00 | 0.00 | 0.00 | 0.00 | 0.00 | 0.00 |
| mratio | 0.01 | 0.00 | 0.00 | 0.00 | 0.00 | 0.00 | 0.00 | 0.00 | 0.00 | 0.00 | 0.00 | 0.00 | 0.00 | 0.00 | 0.00 | 0.00 | 0.00 | 0.00 | 0.00 | 0.00 | 0.00 |
| Diagnosis_GAD | 0.00 | 0.00 | 0.00 | 0.00 | 0.00 | 0.00 | 0.00 | 0.00 | 0.00 | 0.00 | 0.00 | 0.00 | 0.00 | 0.00 | 0.00 | -0.01 | 0.00 | 0.00 | 0.00 | 0.00 | 0.00 |
| Smoking_past_binary | 0.00 | 0.00 | 0.00 | 0.00 | 0.00 | 0.00 | 0.00 | 0.00 | 0.00 | 0.00 | 0.00 | 0.00 | 0.00 | 0.00 | 0.00 | 0.00 | 0.00 | 0.00 | 0.00 | 0.00 | 0.00 |
| mean_response_time | 0.00 | 0.00 | 0.00 | 0.00 | 0.00 | 0.00 | 0.00 | 0.00 | 0.00 | 0.00 | 0.00 | 0.00 | 0.00 | 0.00 | 0.00 | 0.00 | 0.00 | 0.04 | 0.00 | 0.00 | 0.00 |
| Diagnosis_Dep | 0.00 | 0.00 | 0.00 | 0.00 | 0.00 | 0.00 | 0.00 | 0.00 | 0.00 | 0.00 | 0.00 | 0.00 | 0.00 | 0.00 | 0.00 | 0.00 | 0.00 | 0.01 | 0.00 | 0.00 | 0.00 |
| Employment | 0.00 | 0.00 | 0.00 | 0.00 | 0.00 | 0.00 | 0.00 | 0.00 | 0.00 | 0.00 | 0.00 | 0.00 | 0.00 | 0.00 | 0.00 | 0.00 | 0.00 | 0.00 | -0.07 | 0.00 | 0.00 |
| TMT_PTpast | 0.00 | 0.00 | 0.00 | 0.00 | 0.00 | 0.00 | 0.00 | 0.00 | 0.00 | 0.00 | 0.00 | 0.00 | 0.00 | 0.00 | 0.00 | 0.00 | 0.00 | 0.00 | 0.03 | 0.00 | 0.00 |
